# Remote Patient Monitoring in Heart Failure: A Systematic Review, Meta-Analysis, and Trial Sequential Analysis

**DOI:** 10.64898/2026.02.25.26347143

**Authors:** Vicky Muller Ferreira, Victor Ayres Muller

## Abstract

Whether the cumulative evidence for remote patient monitoring (RPM) in heart failure (HF) is robust to sequential monitoring, and whether trials report geographic access modifiers, remains uncertain. We conducted a systematic review, meta-analysis, and trial sequential analysis (TSA) of 65 RCTs (59 poolable; ∼23,000 participants) across four databases through February 2026, encompassing structured telephone support (15 trials), non-invasive telemonitoring (33), and invasive hemodynamic monitoring (11). Random-effects meta-analysis used REML with Hartung-Knapp-Sidik-Jonkman adjustment. RPM significantly reduced all-cause mortality (RR 0.911, 95% CI 0.842–0.985; P=0.021; I^2^=0%; k=41; NNT 104/year; prediction interval 0.840–0.988). TSA suggested that accrued evidence exceeded the required information size under the 15% relative risk reduction assumption, supporting a stable mortality signal. HF hospitalization was reduced (RR 0.781, 95% CI 0.710–0.859; P<0.001; k=39; NNT 18/year), though the prediction interval crossed 1.0 (0.586–1.040), indicating that in some settings the effect may be attenuated. No statistically significant interaction by RPM type was detected (all-cause mortality P_interaction_=0.80; HF hospitalization P_interaction_=0.14). GRADE certainty was moderate for mortality and low for HF hospitalization. A descriptive geographic access analysis revealed that only 2 of 59 poolable trials reported formal rural/urban subgroups, precluding conclusions about whether RPM differentially benefits underserved populations.

**Highlights:**

- Trial sequential analysis supports a stable RPM mortality signal under prespecified assumptions
- All-cause mortality reduced 9% (NNT 104/yr, prediction interval narrowly excludes null)
- HF hospitalization reduced 22% (NNT 18/yr), though prediction interval crosses 1.0
- Large-trial sensitivity remained significant but attenuated for HF hospitalization (RR 0.837)
- Only 2 of 59 poolable trials reported formal rural/urban subgroups

## Introduction

Heart failure (HF) affects over 64 million people worldwide and remains a leading cause of hospitalization, disability, and death [1]. Despite advances in pharmacological therapy, HF hospitalization rates remain high, with 30-day readmission rates exceeding 20% in many health systems [2]. The burden is disproportionately borne by patients in rural and underserved areas, where access to specialized HF care is limited [3].

Remote patient monitoring (RPM) encompasses a spectrum of technologies designed to extend clinical oversight beyond the hospital or clinic setting. Three broad categories have been studied in HF: structured telephone support (STS), in which nurses or automated systems contact patients at regular intervals to assess symptoms and guide self-management; non-invasive telemonitoring (TM), which involves the electronic transmission of physiological data (weight, blood pressure, heart rate, oxygen saturation) from the patient’s home to a clinical team; and invasive hemodynamic monitoring, using implanted sensors (such as the CardioMEMS pulmonary artery pressure sensor) or cardiac implantable electronic device (CIED) algorithms to detect congestion before clinical decompensation.

The evidence base for RPM in HF has grown substantially over the past two decades. The 2015 Cochrane review by Inglis et al. established that both STS and non-invasive TM reduced all-cause mortality compared with usual care, with mortality risk ratios of 0.87 for STS and 0.80 for non-invasive TM [4]. Since then, several landmark trials have been published. The TIM-HF2 trial demonstrated that structured remote management reduced the percentage of days lost to unplanned cardiovascular hospitalization or death [5], while the CHAMPION and MONITOR-HF trials showed that hemodynamic-guided management with the CardioMEMS sensor reduced HF hospitalizations [6,7]. The GUIDE-HF trial, however, yielded neutral overall results, with benefit confined to a pre-COVID sensitivity analysis [8]. The recently published PROACTIVE-HF trial expanded invasive monitoring to HFpEF [9], and the MIGHTY-HEART trial tested a smartphone-based coaching intervention in a diverse urban population, though its primary composite endpoint was neutral [10].

Despite this growing evidence, important questions remain unanswered. First, no prior meta-analysis has applied trial sequential analysis (TSA) to evaluate whether the cumulative evidence for RPM is robust to repeated testing as trials accrue. Second, the most recent comprehensive meta-analysis predates several important trials (PROACTIVE-HF, MIGHTY-HEART, PRADOC, MESSAGE-HF, MONITOR-HF). Third, whether the benefits of RPM extend to geographically underserved populations — where RPM could theoretically provide the greatest incremental value — has not been systematically examined, despite evidence from TIM-HF2 suggesting that patients farther from cardiologists derive greater benefit [5]. Fourth, HF phenotype-specific data (HFrEF, HFpEF, HFmrEF) remain limited, as most trials enrolled mixed populations.

We therefore conducted a systematic review, meta-analysis, and TSA to evaluate the effects of RPM on mortality, hospitalization, and patient-reported outcomes in adults with HF, with the primary objectives of (1) assessing whether the cumulative evidence is robust to sequential monitoring and (2) describing geographic access reporting and exploratory effect modification.

## Materials and Methods

### Protocol and Registration

This systematic review was registered with PROSPERO (CRD420261323628) and conducted in accordance with the Preferred Reporting Items for Systematic Reviews and Meta-Analyses (PRISMA) 2020 guidelines [11]. Searches were completed before registration; this timing and subsequent protocol amendments are documented in the online data repository (DOI: 10.5281/zenodo.19865261). The completed PRISMA 2020 checklist is available in the same repository.

### Eligibility Criteria

We included randomized controlled trials that enrolled adults (age $$18 years) with an established diagnosis of HF (any LVEF phenotype: HFrEF $$40%, HFmrEF 41–49%, or HfpEF $$50%), randomized to any form of RPM (structured telephone support, non-invasive telemonitoring, or invasive hemodynamic monitoring) versus usual care or standard care, with a minimum follow-up of 3 months, reporting at least one outcome of interest (all-cause mortality, HF hospitalization, all-cause hospitalization, cardiovascular mortality, composite outcomes, emergency department visits, or quality of life). Exclusion criteria included observational studies, single-arm trials, studies with fewer than 20 participants, conference abstracts without a full publication, and studies where the RPM intervention could not be isolated from other components (e.g., bundled cardiac rehabilitation programs).

### Information Sources and Search Strategy

We searched PubMed/MEDLINE, Cochrane CENTRAL, ClinicalTrials.gov, and the WHO International Clinical Trials Registry Platform (ICTRP) from inception through February 15, 2026, without language restrictions. The search strategy combined Medical Subject Headings (MeSH) and free-text terms for remote monitoring technologies (including telemonitoring, telehealth, structured telephone support, CardioMEMS, home monitoring, mHealth, wearable devices) and heart failure, with an RCT filter. The full electronic search strategies for all four databases are provided in eTable 1 of the Supplementary Material. The protocol was registered with English-language restriction; however, the search was subsequently conducted without language restriction to maximize sensitivity (protocol amendment #6). Embase and CINAHL were not searched because institutional access was unavailable. To partially mitigate this limitation, we relied on Cochrane CENTRAL, which aggregates records from multiple sources including Embase: 42.9% of the 2,138 CENTRAL records retrieved in our search carried an Embase identifier. PubMed combined with CENTRAL captures approximately 97% of relevant RCTs in therapeutic systematic reviews [12,13]. A post-hoc cross-referencing analysis against three contemporary meta-analyses that searched additional databases [14–16] was conducted to assess the potential impact of this omission (eText 1).

### Study Selection

Records were imported, combined, and deduplicated using a multi-pass approach (exact DOI/PMID matching, normalized title matching, and fuzzy matching with manual adjudication). Title and abstract screening used a semi-automated approach with predefined exclusion rules validated against a reference set of known eligible trials. Records flagged by the algorithm (n=219, 8.3%) underwent manual adjudication with final consensus decisions retained in the screening dataset. A post-hoc sensitivity audit of the screening algorithm identified two false negatives due to missing search terms (telemanagement, CHF), which were rescued and included. Records passing initial screening underwent a structured audit to exclude registry-only entries, protocols, conference abstracts, and non-RCT designs. All remaining records were assessed at full-text level using structured eligibility tiers. The archived screening datasets preserve final consensus decisions rather than complete independent reviewer-level decisions; therefore, the process should be interpreted as semi-automated screening with manual adjudication and audit rather than fully independent dual screening. The PRISMA 2020 flow diagram is presented in Figure 1.

**Figure 1.**
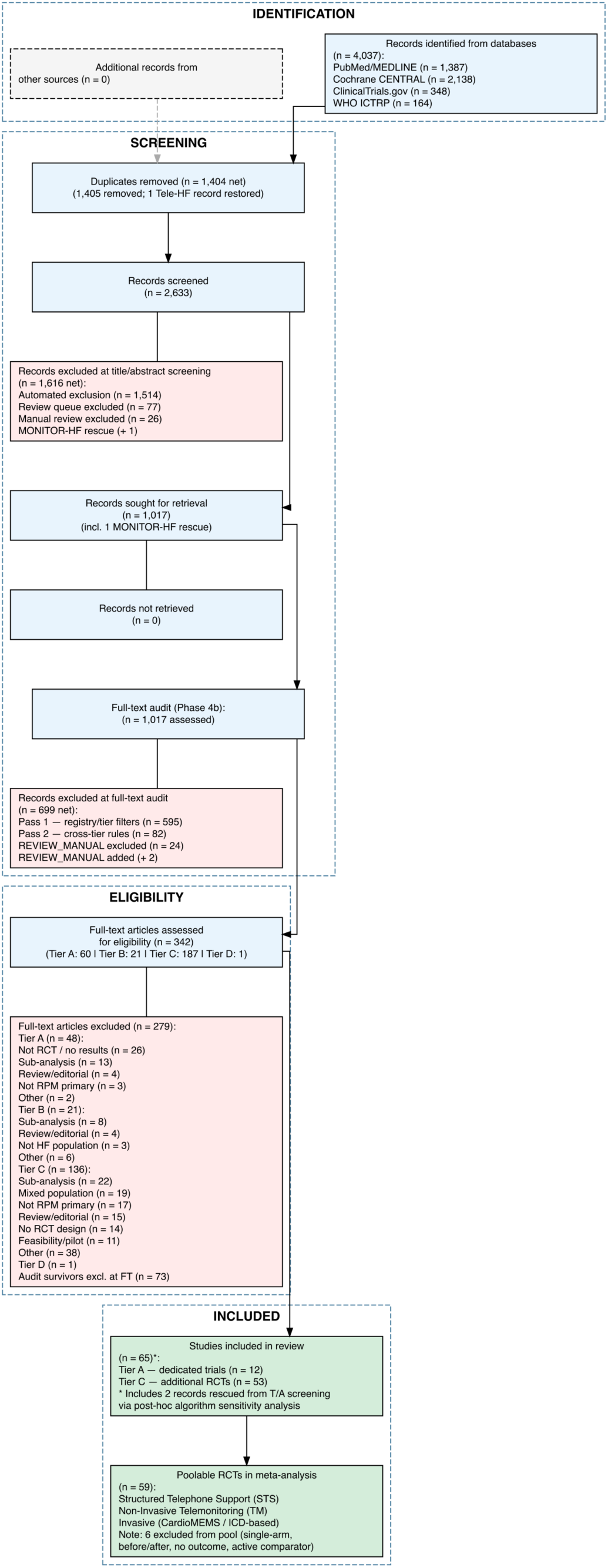
PRISMA 2020 flow diagram of study selection. A total of 4,037 records were identified across four databases. After deduplication, screening, and full-text assessment, 65 studies were included and 59 were poolable for quantitative synthesis.

### Data Extraction

Data were extracted using a two-reviewer process. One reviewer (VMF) performed the initial structured extraction with AI-assisted tabulation support, and the second reviewer (VAM) independently reviewed the extracted data against source reports; discrepancies were resolved by consensus before analysis. The archived extraction dataset reports the final consensus values used for analysis; reviewer-level intermediate disagreement fields were not retained for every study. Quality checks identified and corrected three errors during the consensus process: MONITOR-HF HF hospitalization HR (0.82 corrected to 0.56, verified against original publication Table 2), CHAMPION PMID, and the CO-0056 mortality denominator (corrected from the pooled intervention denominator to the telephone arm denominator). Extracted effect estimates were checked against original publication tables and supplementary materials where available; extraction notes and a consensus log documenting reviewer roles, correction flags, and final verification status for all 65 studies are available in the Zenodo repository. Data were extracted using a standardized form encompassing 185 variables, including study characteristics, participant demographics, intervention details, geographic context (rural/urban classification, distance to care), outcomes (event counts for binary outcomes; means and standard deviations for continuous outcomes; hazard ratios with 95% confidence intervals when reported), and risk of bias domains. RPM interventions were classified into three pre-specified categories: structured telephone support (STS), non-invasive telemonitoring, and invasive hemodynamic monitoring.

**Table 1.**
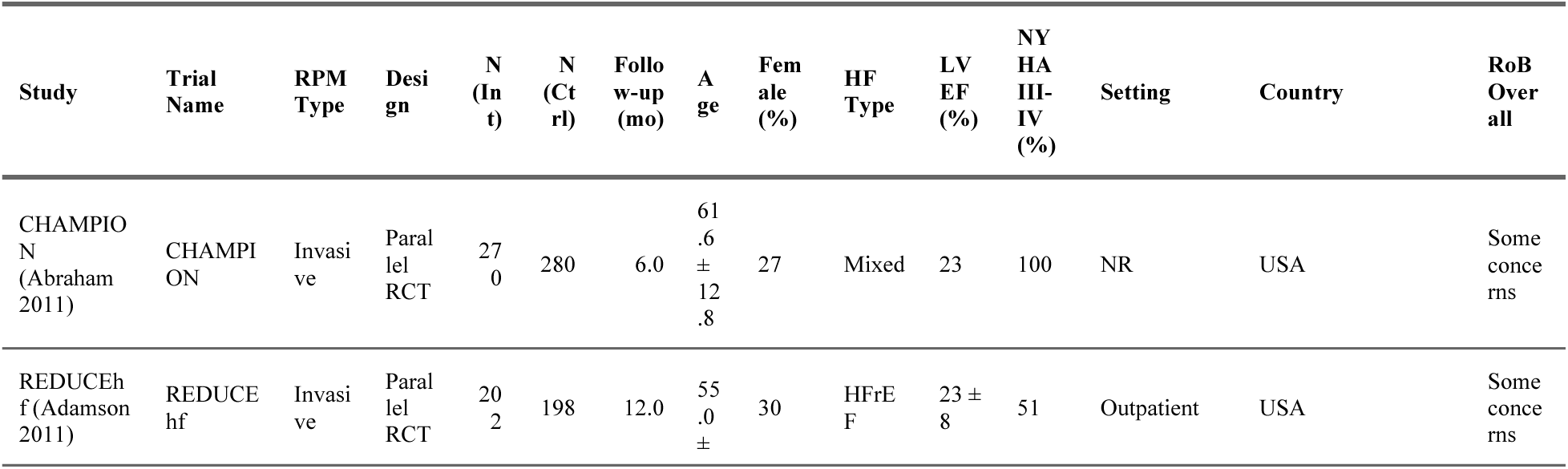
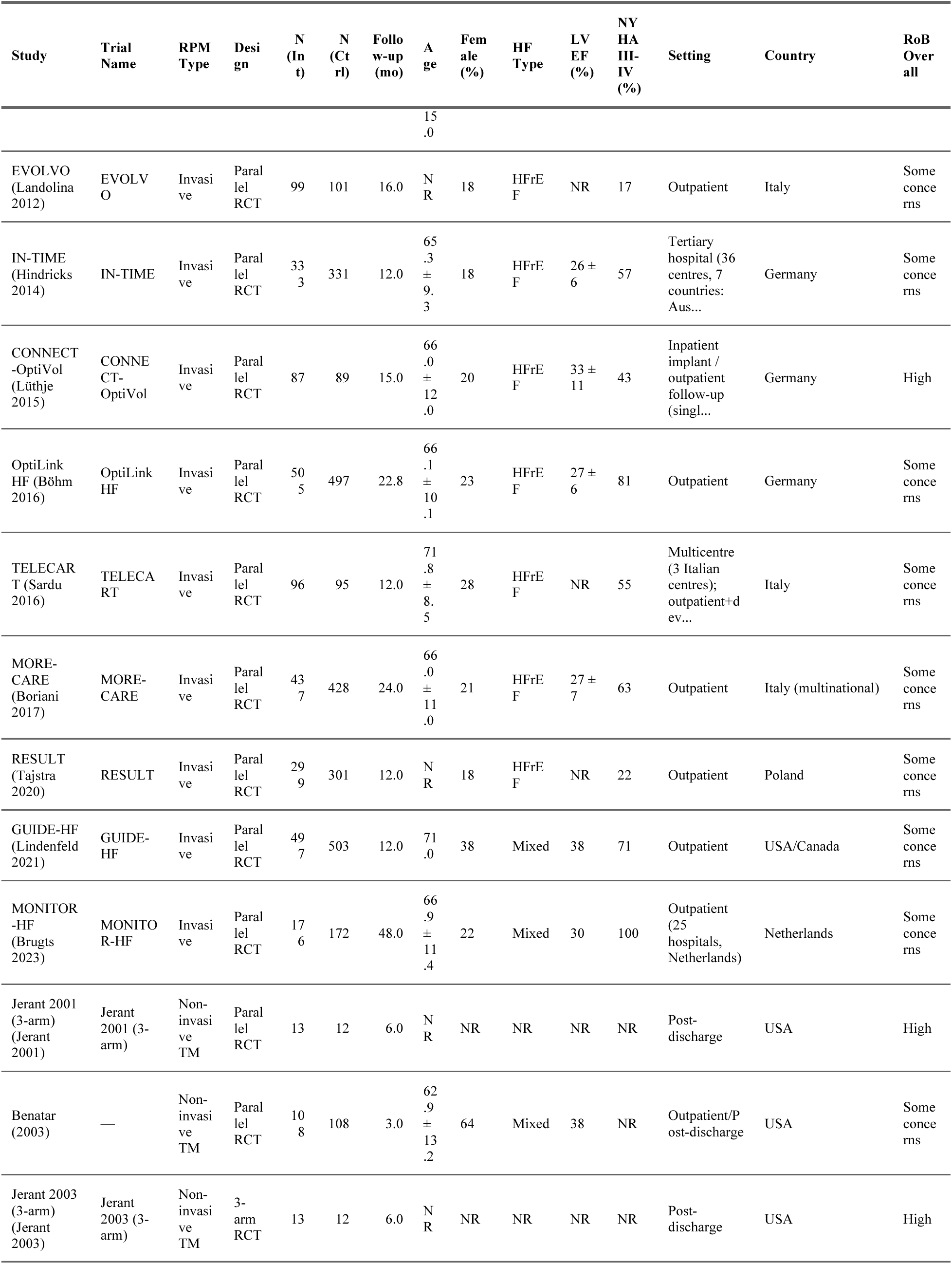
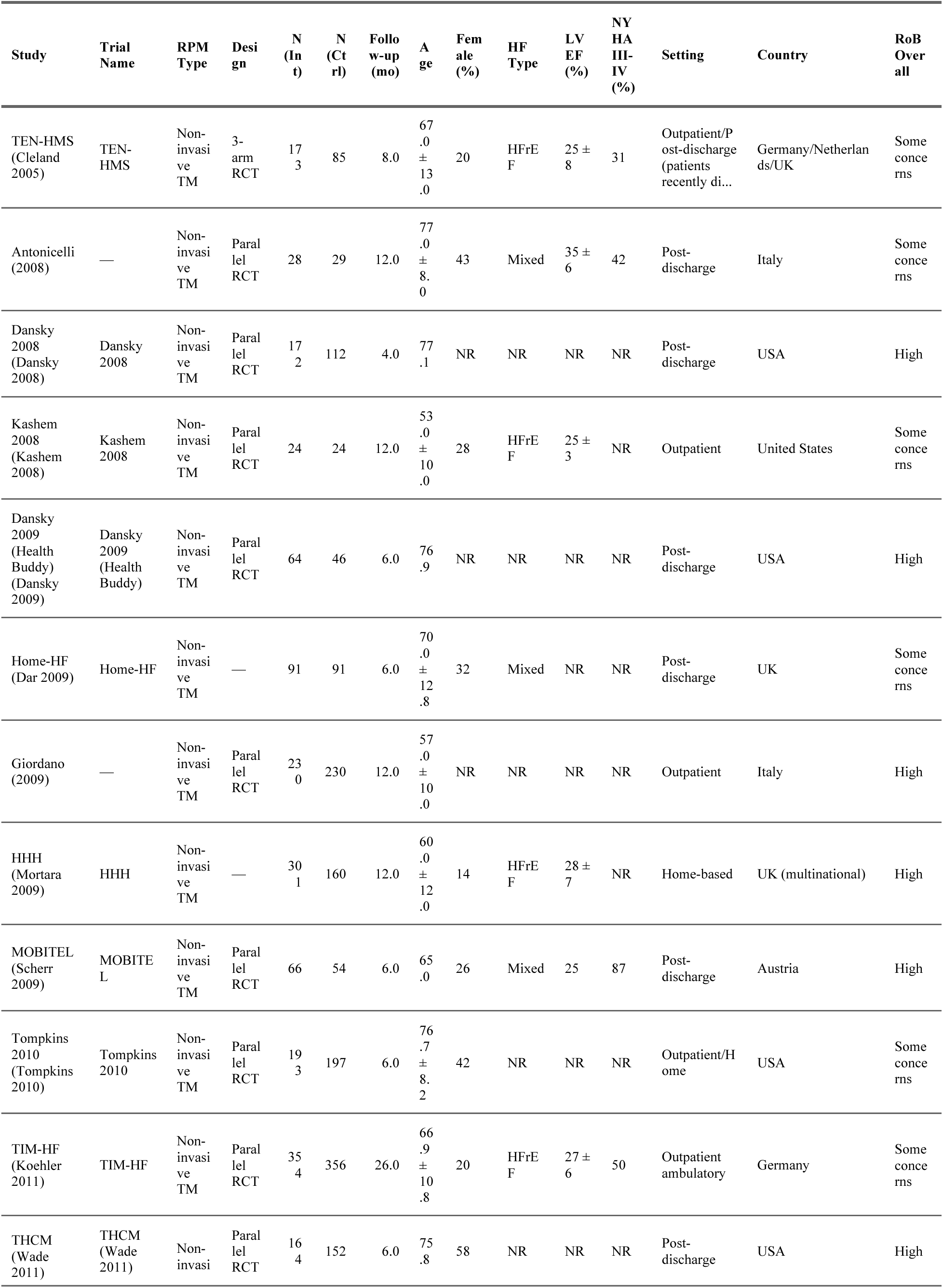
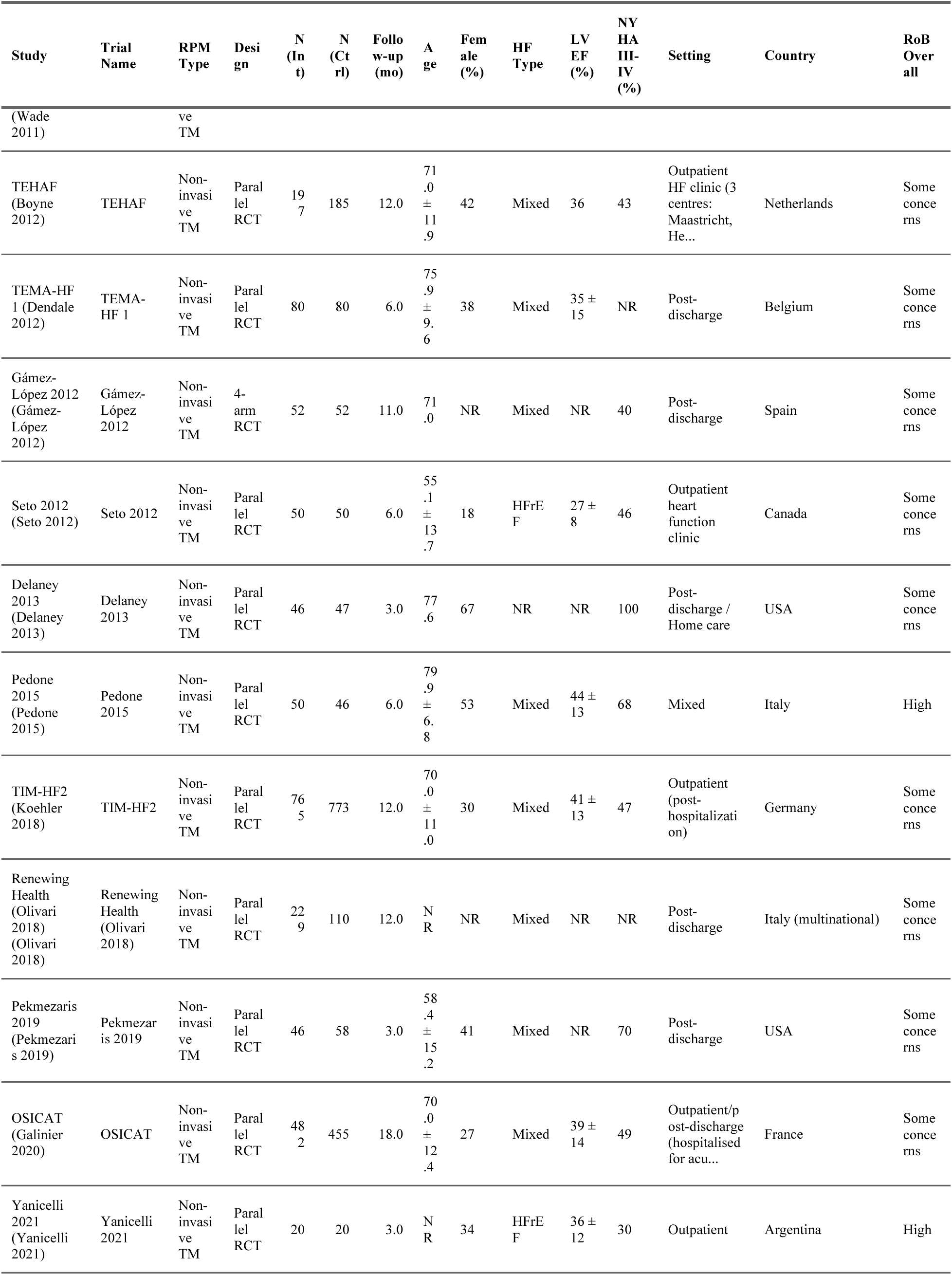
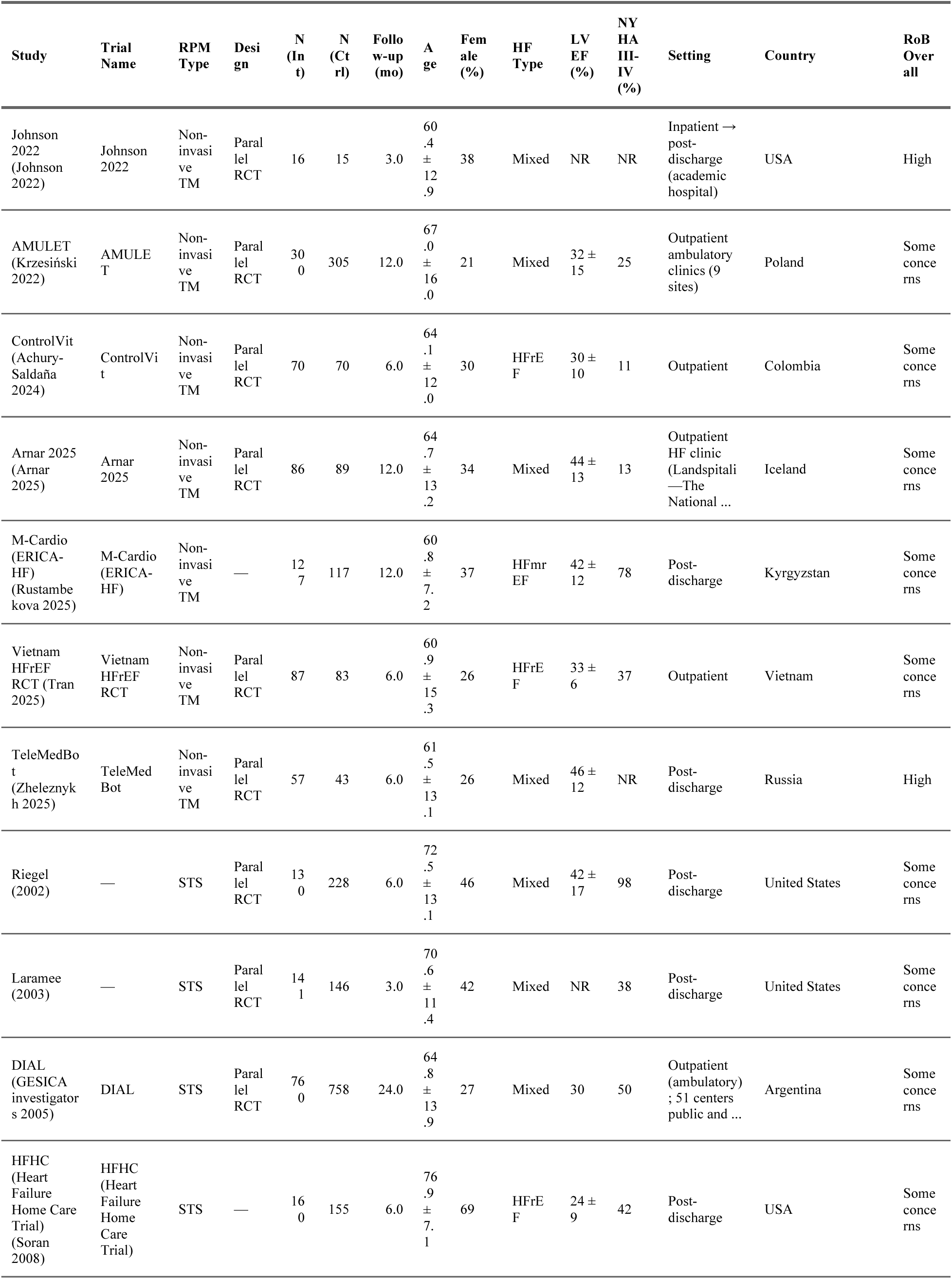
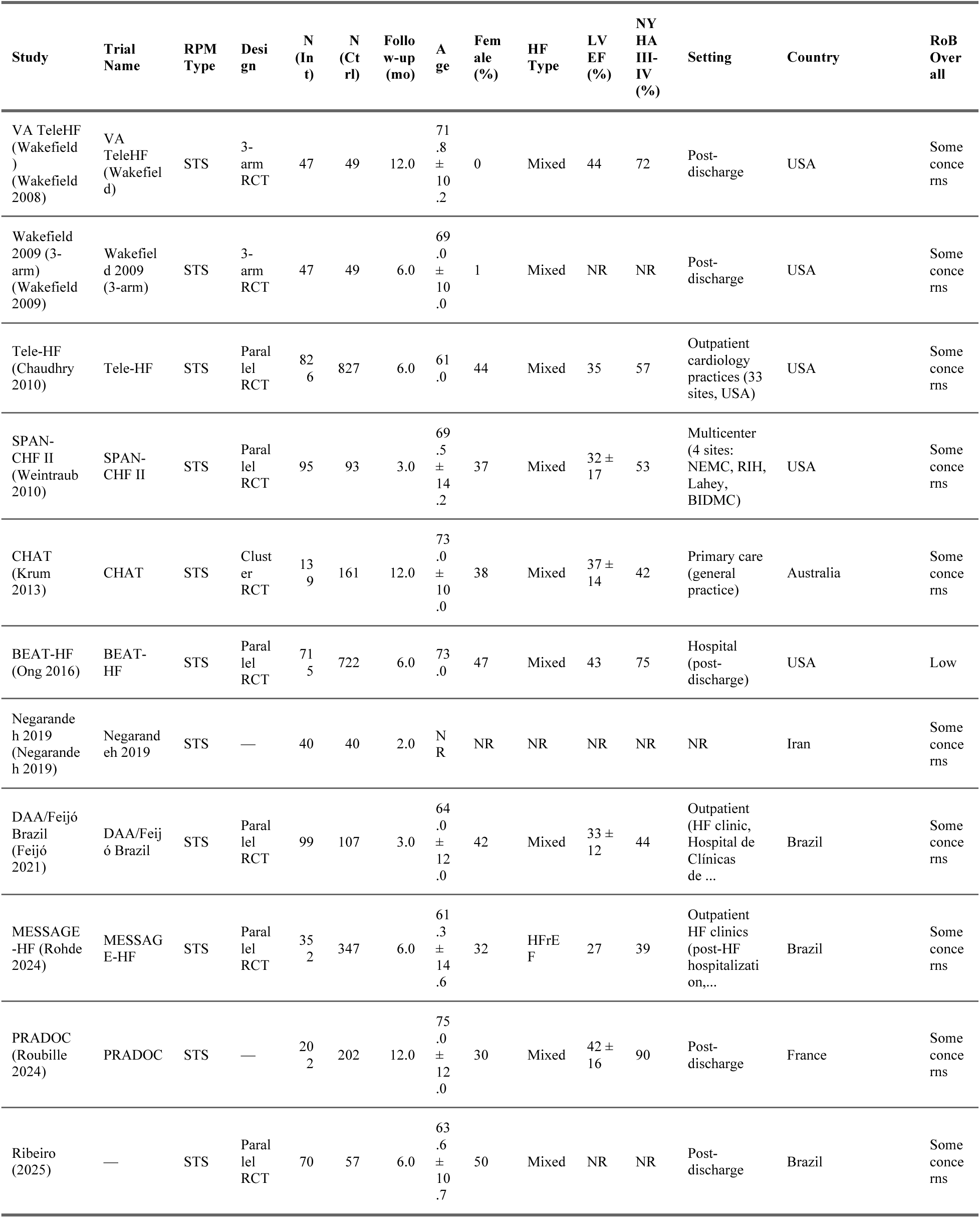
Characteristics of Included Studies.

### Risk of Bias Assessment

Risk of bias was assessed using the Cochrane Risk of Bias 2 (RoB 2) tool [17] across five domains: randomization process (D1), deviations from intended interventions (D2), missing outcome data (D3), measurement of the outcome (D4), and selection of the reported result (D5). All RPM trials are inherently open-label for participants and clinical staff; therefore, D2 ratings reflect whether knowledge of group assignment could have influenced co-interventions or adherence beyond the intended effect.

### Effect Measures and Statistical Analysis

The primary outcomes were all-cause mortality, HF hospitalization, and all-cause hospitalization. Secondary outcomes included cardiovascular mortality, composite of all-cause mortality and hospitalization, emergency department visits, and quality of life (Minnesota Living with Heart Failure Questionnaire [MLHFQ] and Kansas City Cardiomyopathy Questionnaire [KCCQ]).

For binary outcomes with event counts, risk ratios (RR) were calculated using the Mantel-Haenszel method. Rows in which event counts represented recurrent events or episodes rather than participants with events (events exceeding denominators) were excluded from binary RR pooling and from control event rate/NNT calculations. Studies reporting only hazard ratios (HR) with confidence intervals — without extractable event counts — were not included in the primary RR analysis but were pooled separately using the generic inverse-variance method with log-transformed HRs and corresponding standard errors. This yielded two parallel analyses for each primary outcome: a primary RR analysis maximizing the number of contributing studies with valid participant-level event counts (e.g., k=41 for all-cause mortality) and a sensitivity HR-only analysis restricted to time-to-event data (e.g., k=16 for all-cause mortality). Continuous outcomes were pooled as mean differences (MD). RR and HR estimates are reported separately throughout and are not combined in a single model, as mixing effect measures can introduce bias [18]. For trials reporting outcomes at multiple time points, events at the protocol-specified primary endpoint were extracted (e.g., CHAMPION at 6 months rather than extended follow-up at 15–18 months). The cluster-randomized trial (CHAT) was adjusted using the design effect method (DE = 1.35, based on ICC = 0.02 and mean cluster size 18.4), with both event counts and denominators reduced per Cochrane Handbook Section 23.1.3 [19].

Random-effects meta-analysis was performed using restricted maximum likelihood (REML) estimation for the between-study variance (𝜏^2^) with the Hartung-Knapp-Sidik-Jonkman (HKSJ) adjustment for confidence intervals [20,21]. Statistical heterogeneity was assessed using I^2^, 𝜏^2^, Cochran’s Q test, and 95% prediction intervals [22,23].

### Subgroup and Sensitivity Analyses

Pre-specified subgroup analyses examined the influence of RPM type (STS, non-invasive TM, invasive), HF phenotype (HFrEF, HFpEF, HFmrEF, mixed), geographic context (rural/remote, urban, mixed, not reported), country income level (high-income versus other), follow-up duration ($6𝑚𝑜𝑛𝑡ℎ𝑠𝑣𝑒𝑟𝑠𝑢𝑠 > 6𝑚𝑜𝑛𝑡ℎ𝑠), 𝑝𝑢𝑏𝑙𝑖𝑐𝑎𝑡𝑖𝑜𝑛𝑒𝑟𝑎($2015 versus >2015), and risk of bias (high versus low/some concerns). Interaction P values were calculated using the Q-test for between-subgroup differences.

Ten pre-specified sensitivity analyses were performed for each primary outcome: (1) leave-one-out analysis, (2) fixed-effect (common-effect) model, (3) exclusion of active comparator arms, (4) exclusion of invasive RPM studies, (5) exclusion of cluster-randomized trials, (6) exclusion of high risk of bias studies, (7) exclusion of flagged outliers, (8) restriction to HR-reporting studies only, (9) restriction to follow-up $3𝑚𝑜𝑛𝑡ℎ𝑠, 𝑎𝑛𝑑(10)𝑟𝑒𝑠𝑡𝑟𝑖𝑐𝑡𝑖𝑜𝑛𝑡𝑜𝑙𝑎𝑟𝑔𝑒𝑡𝑟𝑖𝑎𝑙𝑠(𝑁$200). Meta-regression examined the association of treatment effect with total sample size, publication year, follow-up duration, mean age, and mean LVEF.

### Publication Bias

Publication bias was assessed visually using funnel plots and statistically using Egger’s regression test [24], Begg’s rank correlation test, and Peters’ regression test. The trim-and-fill method was applied to estimate the impact of potential missing studies [25]. Contour-enhanced funnel plots were generated to distinguish asymmetry due to publication bias from asymmetry due to other causes. A large-trial sensitivity analysis (N$$200) was performed to assess whether small-study effects drove the overall signal.

### Trial Sequential Analysis

Trial sequential analysis (TSA) was performed using 𝛼=0.05, 𝛽=0.20 (80% power), and relative risk reductions (RRR) of 15% and 20%, with O’Brien-Fleming spending boundaries [26]. TSA was used as a sensitivity framework to assess whether cumulative evidence was robust to repeated testing under the specified assumptions.

### Certainty of Evidence

The certainty of evidence was assessed using the Grading of Recommendations, Assessment, Development, and Evaluations (GRADE) framework [27] across five domains: risk of bias, inconsistency, indirectness, imprecision, and publication bias.

All analyses were performed in R (version 4.5.2) using the meta [28] and metafor [29] packages. Statistical significance was defined as P < 0.05 (two-sided).

## Results

### Study Selection

The systematic search identified 4,037 records across four databases: PubMed (n=1,387), Cochrane CENTRAL (n=2,138), ClinicalTrials.gov (n=348), and WHO ICTRP (n=164). After identifying 1,405 duplicate records and rescuing one that had been incorrectly flagged, 1,404 records were removed, yielding 2,633 unique records for title and abstract screening. Of these, 1,616 were excluded, and 1,017 records proceeded to full-text assessment. A structured audit excluded 701 records (registry entries, protocols, conference abstracts, non-RCTs), while 26 records were added or reclassified into the full-text pool (Tier B candidates, manual review survivors, and pipeline rescues), yielding 342 records for full-text review across four tiers (A–D). After detailed eligibility evaluation, 65 studies met all inclusion criteria (including two records rescued from title/abstract screening via post-hoc algorithm sensitivity analysis), of which 59 were poolable for quantitative synthesis (Figure 1).

### Study Characteristics

The 65 included RCTs encompassed approximately 23,000 participants across 20 countries. RPM interventions were classified as structured telephone support (15 trials), non-invasive telemonitoring (33 trials), or invasive hemodynamic monitoring (11 trials), with 6 trials excluded from quantitative synthesis because they used active comparators, had single-arm or before/after designs, or reported only outcomes incompatible with pooling (eTable 6). The median sample size was 244 (range 25–1,653) and the median follow-up was 6 months (range 2–48 months; one study [Negarandeh 2019] had 2-month follow-up, retained by investigator decision as a borderline case). HF phenotype was predominantly HFrEF or mixed; no trials enrolled HFpEF patients exclusively among the 59 poolable studies.

### Risk of Bias

The majority of studies (44 of 59) were judged as having “some concerns” for overall risk of bias, primarily driven by the open-label design inherent to RPM interventions (D2: deviations from intended interventions). Thirteen studies were rated as high risk of bias, and two as low risk. Detailed RoB 2 assessments are presented in eFigures 1–2 and eTable 2.

**Figure 2.**
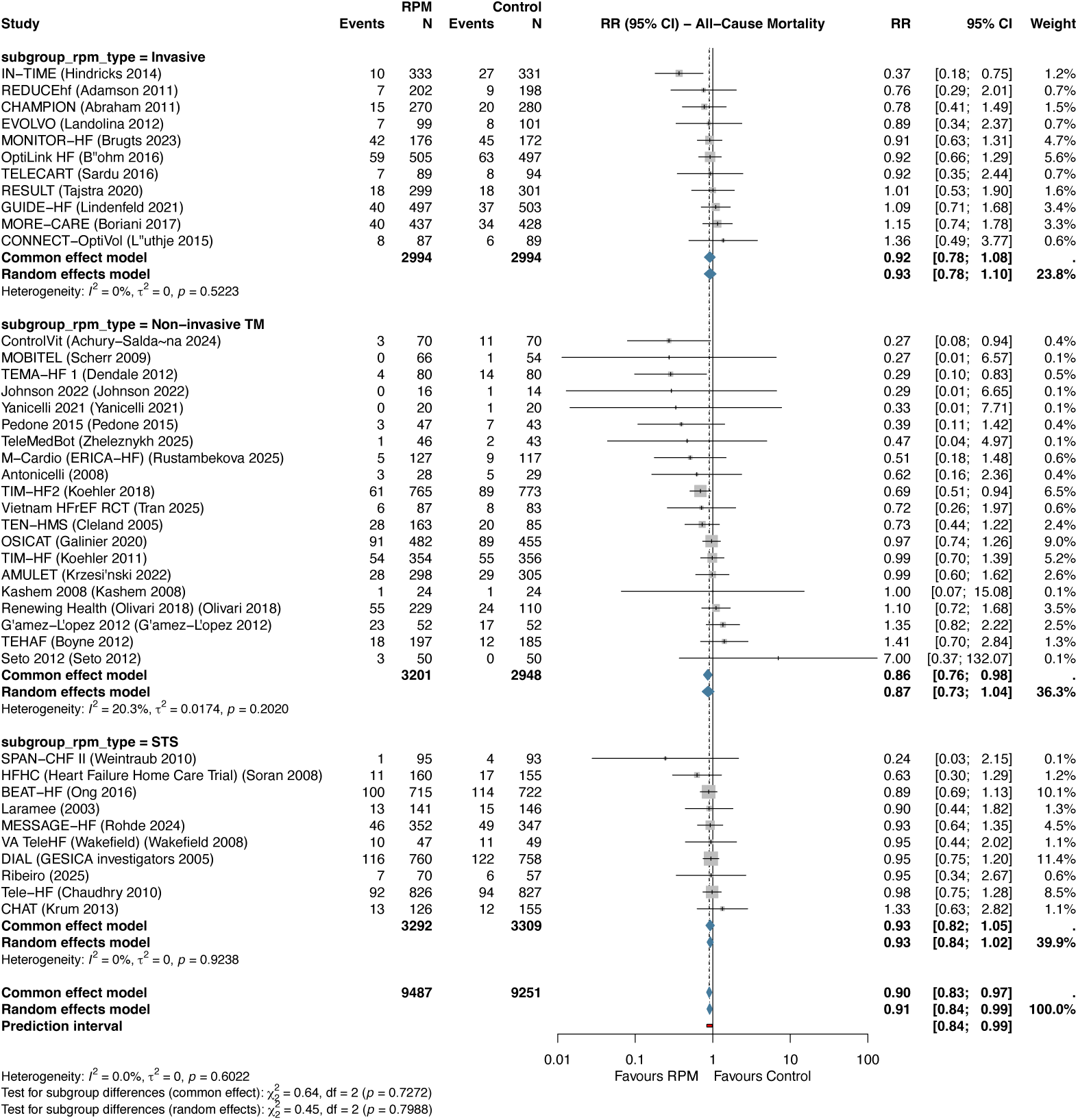
Forest plot of all-cause mortality. Risk ratios with 95% confidence intervals from random-effects meta-analysis (REML + HKSJ). The diamond represents the pooled estimate (RR 0.911, 95% CI 0.842–0.985; k=41).

### Primary Outcomes

#### All-Cause Mortality

**Table 2.** Summary of Meta-Analysis Results.

| Category | Outcome | Measure | k | Effect [95% CI] | p-value | I<sup>2</sup> (%) | Prediction Interval |
| --- | --- | --- | --- | --- | --- | --- | --- |
| Primary outcomes (RR, Mantel-Haenszel) | All-cause mortality (primary) | RR | 41 | 0.911 [0.842, 0.985] | 0.0208 | 0.0 | [0.840, 0.988] |
| Primary outcomes (RR, Mantel-Haenszel) | HF hospitalization (primary) | RR | 39 | 0.781 [0.710, 0.859] | 0.0000 | 47.2 | [0.586, 1.040] |
| Primary outcomes (RR, Mantel-Haenszel) | All-cause hospitalization (primary) | RR | 28 | 0.959 [0.892, 1.031] | 0.2462 | 54.3 | [0.809, 1.137] |
| Primary sensitivity (HR subset, metagen) | All-cause mortality | HR | 16 | 0.880 [0.784, 0.989] | 0.0335 | 0.0 | [0.783, 0.989] |
| Primary sensitivity (HR subset, metagen) | HF hospitalization | HR | 13 | 0.766 [0.698, 0.841] | 0.0000 | 1.1 | [0.698, 0.841] |
| Primary sensitivity (HR subset, metagen) | All-cause hospitalization | HR | 10 | 0.870 [0.711, 1.064] | 0.1514 | 71.0 | [0.505, 1.497] |
| Secondary outcomes (ratio) | CV Mortality | RR | 9 | 0.814 [0.668, 0.992] | 0.0433 | 0.0 | — |
| Secondary outcomes (ratio) | ED Visits | RR | 6 | 0.795 [0.550, 1.149] | 0.1699 | 41.4 | — |
| Secondary outcomes (ratio) | Composite ACM+Hospitalization | HR | 25 | 0.827 [0.765, 0.894] | 0.0000 | 37.5 | — |
| Secondary outcomes (continuous, MD) | MLHFQ | MD | 11 | -6.53 [-10.28, -2.79] | 0.0030 | 93.6 | — |
| Secondary outcomes (continuous, MD) | KCCQ | MD | 3 | 8.14 [-24.03, 40.30] | 0.3902 | 83.7 | — |

RPM significantly reduced all-cause mortality across 41 RCTs (RR 0.911, 95% CI 0.842–0.985; P=0.021; I^2^=0%; 𝜏^2^=0) (Figure 2, Table 2). The 95% prediction interval was 0.840–0.988, excluding the null under the random-effects model. The number needed to treat was 104 per year. In sensitivity analysis restricted to studies reporting hazard ratios (k=16), the result was consistent (HR 0.880, 95% CI 0.784–0.989; P=0.034).

#### HF Hospitalization

RPM significantly reduced HF hospitalization across 39 RCTs (RR 0.781, 95% CI 0.710–0.859; P<0.001; I^2^=47.2%; 𝜏^2^=0.018) (Figure 3). The 95% prediction interval was 0.586–1.040, crossing 1.0, consistent with moderate heterogeneity. The NNT was 18 per year. In HR sensitivity (k=13), the effect was more pronounced (HR 0.766, 95% CI 0.698–0.841; P<0.001; I^2^=1.1%).

**Figure 3.**
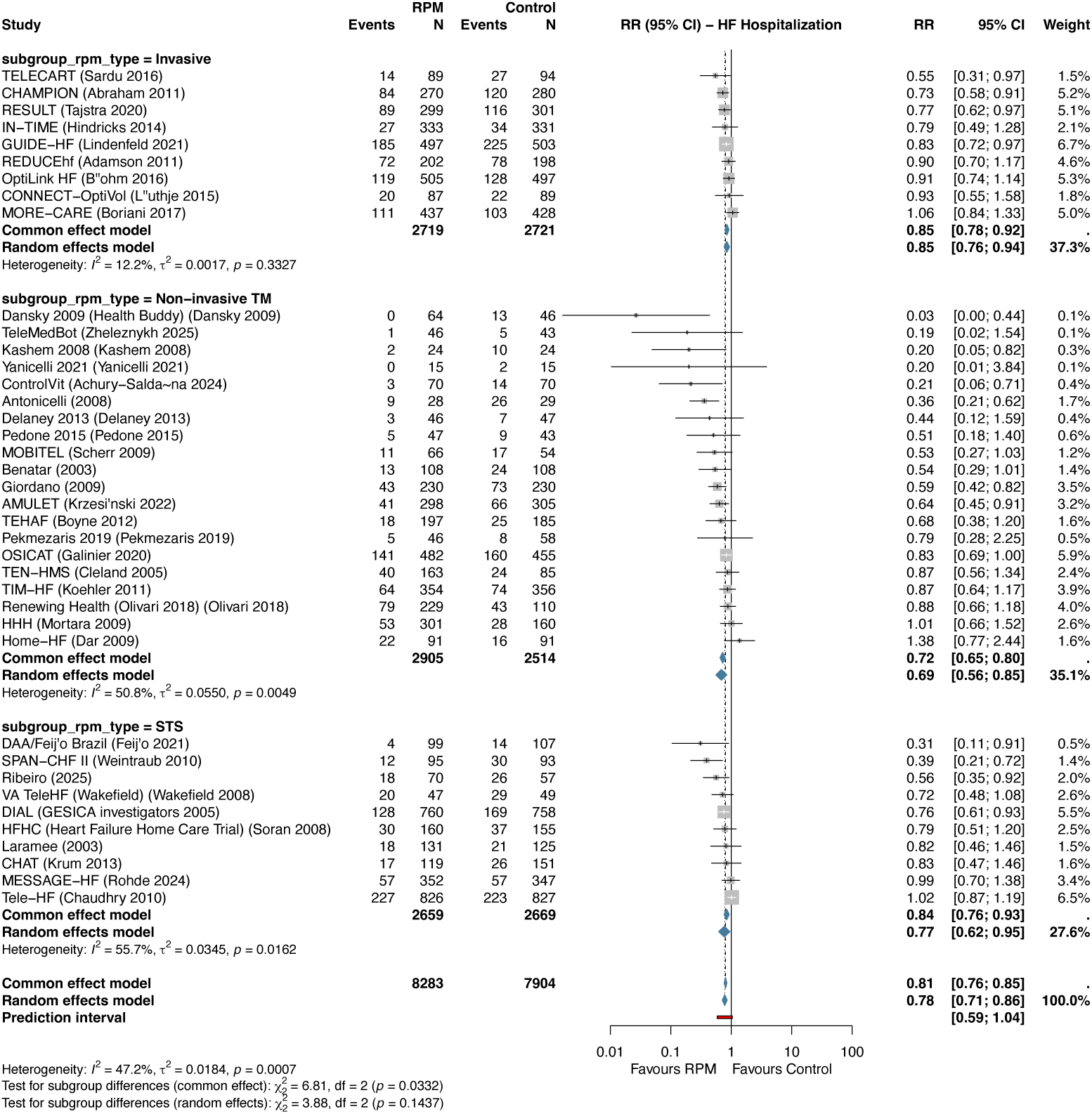
Forest plot of heart failure hospitalization. Pooled RR 0.781 (95% CI 0.710–0.859; I^2^=47.2%; k=39).

#### All-Cause Hospitalization

RPM did not significantly reduce all-cause hospitalization across 28 RCTs (RR 0.959, 95% CI 0.892–1.031; P=0.246; I^2^=54.3%) (Figure 4). The prediction interval was 0.809–1.137.

**Figure 4.**
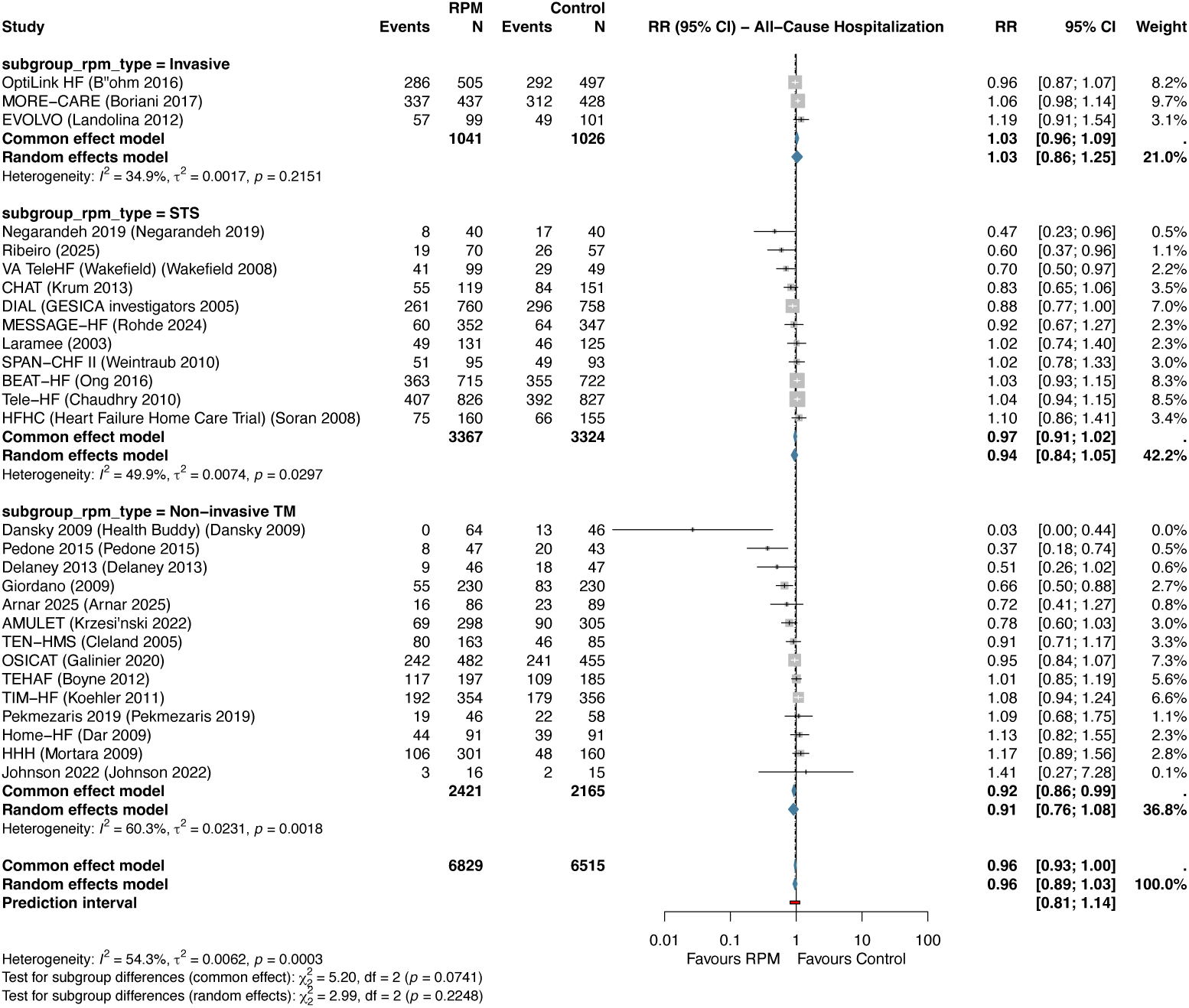
Forest plot of all-cause hospitalization. Pooled RR 0.959 (95% CI 0.892–1.031; I^2^=54.3%; k=28).

### Secondary Outcomes

Cardiovascular mortality was significantly reduced (RR 0.814, 95% CI 0.668–0.992; P=0.043; I^2^=0%; k=9; low certainty after downgrading for imprecision). The composite of all-cause mortality and hospitalization was significantly reduced (HR 0.827, 95% CI 0.765–0.894; P<0.001; I^2^=37.5%; k=25; low certainty after downgrading for indirectness due to mixed effect measures and inconsistency). Emergency department visits showed a non-significant reduction (RR 0.795, 95% CI 0.550–1.149; P=0.17; I^2^=41.4%; k=6). Quality of life measured by MLHFQ improved (MD –6.53 points, 95% CI –10.28 to –2.79; P=0.003; I^2^=93.6%; k=11), favoring RPM (lower scores indicate better quality of life), though the very high heterogeneity makes the pooled mean difference difficult to interpret. KCCQ showed a non-significant improvement (MD 8.14, 95% CI –24.03 to 40.30; P=0.39; I^2^=83.7%; k=3).

### Subgroup Analyses

No statistically significant interaction by RPM type was detected for any primary outcome, though the analysis was likely underpowered to detect moderate subgroup differences given the imbalanced subgroup sizes (STS k=10, TM k=20, Invasive k=11 for ACM). For all-cause mortality, the test for subgroup differences yielded P_interaction_=0.80, with similar point estimates across subgroups (non-invasive TM RR 0.871; invasive monitoring RR 0.925; STS RR 0.928). For HF hospitalization, P_interaction_=0.14, and for all-cause hospitalization, P_interaction_=0.22 (Table 2, Figure 7).

HF phenotype showed a significant interaction for all-cause hospitalization (P_interaction_=0.002), with mixed HF phenotype trials showing a directionally favorable estimate (RR 0.940) compared with HFrEF-only trials (RR 1.037). This was not significant for all-cause mortality (P_interaction_=0.46) and was borderline for HF hospitalization (P_interaction_=0.056). The single trial classified as HFmrEF in the subgroup analysis (M-Cardio [ERICA-HF], mean LVEF 42%) contributed a directionally favorable but imprecise estimate, though this subgroup comprises only one study and should be interpreted cautiously.

Geographic context was examined as a descriptive subgroup. Among 41 trials reporting all-cause mortality, 13 were classified as rural/remote and 28 as not reported. No significant interaction was observed (P_interaction_=0.90). The sparse reporting of geographic context — only two trials (TIM-HF2 and CHAT) provided formal rural/urban subgroup analyses — limited the interpretability of this analysis.

No significant interaction was observed for follow-up duration ($6𝑣𝑠 >

6𝑚𝑜𝑛𝑡ℎ𝑠), 𝑝𝑢𝑏𝑙𝑖𝑐𝑎𝑡𝑖𝑜𝑛𝑒𝑟𝑎($2015 vs >2015), country income level (eFigure 8), or risk of bias for any primary outcome (all P_interaction_>0.05).

In multivariable meta-regression for HF hospitalization, total sample size was borderline rather than statistically significant after excluding recurrent-count rows from the binary RR analysis (𝛽=0.00026, P=0.063). Mean LVEF was associated with treatment effect (𝛽=–0.032, P=0.0049), but this ecological finding should be interpreted cautiously because study-level covariates can be confounded by intervention type, era, and patient selection. No covariates were significant predictors for all-cause mortality.

### Sensitivity Analyses

All 10 sensitivity analyses were directionally consistent with the primary results for both all-cause mortality and HF hospitalization (Table 3). For all-cause mortality, leave-one-out analysis showed pooled RR ranging from 0.901 to 0.928, with no single study disproportionately influencing the result. The fixed-effect model was consistent (RR 0.900, P=0.009). Excluding invasive RPM studies yielded RR 0.906 (P=0.038), excluding high RoB studies yielded RR 0.914 (P=0.031), and restricting to large trials (N$$200) yielded RR 0.918 (P=0.033; k=24). For HF hospitalization, all sensitivities remained statistically significant, including the large-trial sensitivity (RR 0.837, 95% CI 0.779–0.899; k=23).

**Table 3.** Sensitivity Analyses for Primary Outcomes.

| Outcome | Analysis | k | RR | LCI | UCI | P | I<sup>2</sup><br>(%) | Note |
| --- | --- | --- | --- | --- | --- | --- | --- | --- |
| ACM | 10_large_trials_n200 | 24 | 0.918 | 0.849 | 0.993 | 0.0331 | 0.0 | N≥200 only (n excl.=17) |

| Outcome | Analysis | k | RR | LCI | UCI | P | I<sup>2</sup> (%) | Note |
| --- | --- | --- | --- | --- | --- | --- | --- | --- |
| ACM | 11_incl_active_comparators | 42 | 0.918 | 0.848 | 0.995 | 0.0372 | 0.0 | Post-hoc: including MIGHTY-HEART active comparator (CO-0570) |
| ACM | 2_fixed_effect | 41 | 0.900 | 0.832 | 0.974 | 0.0087 | 0.0 | Fixed-effect (common); no HKSJ correction |
| ACM | 3_usual_care_only | 37 | 0.910 | 0.839 | 0.986 | 0.023 | 0.0 | Comparator type = usual care only (n excl.=4) |
| ACM | 4_excl_invasive | 30 | 0.906 | 0.826 | 0.994 | 0.0382 | 0.0 | Excl. Invasive RPM (n excl.=11) |
| ACM | 5_excl_cluster_rct | 40 | 0.907 | 0.838 | 0.981 | 0.0165 | 0.0 | Excl. CHAT (CO-0944) cluster RCT |
| ACM | 6_low_rob_only | 35 | 0.914 | 0.843 | 0.992 | 0.0314 | 0.0 | Excl. High RoB (n excl.=6) |
| ACM | 7_excl_outliers | 36 | 0.930 | 0.868 | 0.998 | 0.043 | 0.0 | Excl. flagged outliers (n excl.=5) |
| ACM | 8_hr_only_subset | 16 | 0.880 | 0.784 | 0.989 | 0.0335 | 0.0 | HR-reporting studies only (metagen); k=16 |
| ACM | 9_fu_ge3mo | 41 | 0.911 | 0.842 | 0.985 | 0.0208 | 0.0 | FU >=3 months only (n excl.=0) |
| HF_hosp | 10_large_trials_n200 | 23 | 0.837 | 0.779 | 0.899 | <0.001 | 21.8 | N>=200 only (n excl.=16); excluded recurrent-count rows incompatible with binary RR (n=2) |
| HF_hosp | 11_incl_active_comparators | 40 | 0.790 | 0.721 | 0.865 | <0.001 | 46.7 | Post-hoc: including MIGHTY-HEART active comparator (CO-0570); excluded recurrent-count rows incompatible with binary RR (n=2) |
| HF_hosp | 2_fixed_effect | 39 | 0.805 | 0.763 | 0.850 | <0.001 | 47.2 | Fixed-effect (common); no HKSJ correction |
| HF_hosp | 3_usual_care_only | 33 | 0.793 | 0.723 | 0.870 | <0.001 | 43.7 | Comparator type = usual care only (n excl.=7); excluded recurrent-count rows incompatible with binary RR (n=1) |
| HF_hosp | 4_excl_invasive | 30 | 0.725 | 0.629 | 0.835 | <0.001 | 53.0 | Excl. Invasive RPM (n excl.=11) |
| HF_hosp | 5_excl_cluster_rct | 38 | 0.779 | 0.706 | 0.859 | <0.001 | 48.6 | Excl. CHAT (CO-0944) cluster RCT; excluded recurrent-count rows incompatible with binary RR (n=2) |
| HF_hosp | 6_low_rob_only | 31 | 0.797 | 0.724 | 0.877 | <0.001 | 46.3 | Excl. High RoB (n excl.=8); excluded recurrent-count rows incompatible with binary RR (n=2) |
| HF_hosp | 7_excl_outliers | 33 | 0.812 | 0.750 | 0.878 | <0.001 | 33.3 | Excl. flagged outliers (n excl.=6); excluded recurrent-count rows incompatible with binary RR (n=2) |
| HF_hosp | 8_hr_only_subset | 13 | 0.766 | 0.698 | 0.841 | <0.001 | 1.1 | HR-reporting studies only (metagen); k=13 |
| HF_hosp | 9_fu_ge3mo | 39 | 0.781 | 0.710 | 0.859 | <0.001 | 47.2 | FU >=3 months only (n excl.=0); excluded recurrent-count rows incompatible with binary RR (n=2) |

### Publication Bias

For all-cause mortality, Egger’s test was significant (P=0.040) and Begg’s test was significant (P=0.014), suggesting funnel plot asymmetry. Peters’ regression test was non-significant (P=0.246), and the large-trial sensitivity analysis (N$$200; k=24) yielded RR 0.918 (P=0.033), indicating that the mortality reduction persisted when small studies were excluded. Trim-and-fill analysis estimated 6 missing studies on the right side and yielded an adjusted RR of 0.925 (95% CI 0.851–1.005; P=0.064), attenuating the mortality estimate to borderline/non-significance (Figure 6). The contour-enhanced funnel plot suggested that asymmetry was driven primarily by small studies in regions of statistical significance, consistent with small-study effects; selective non-publication cannot be excluded.

**Figure 5.**
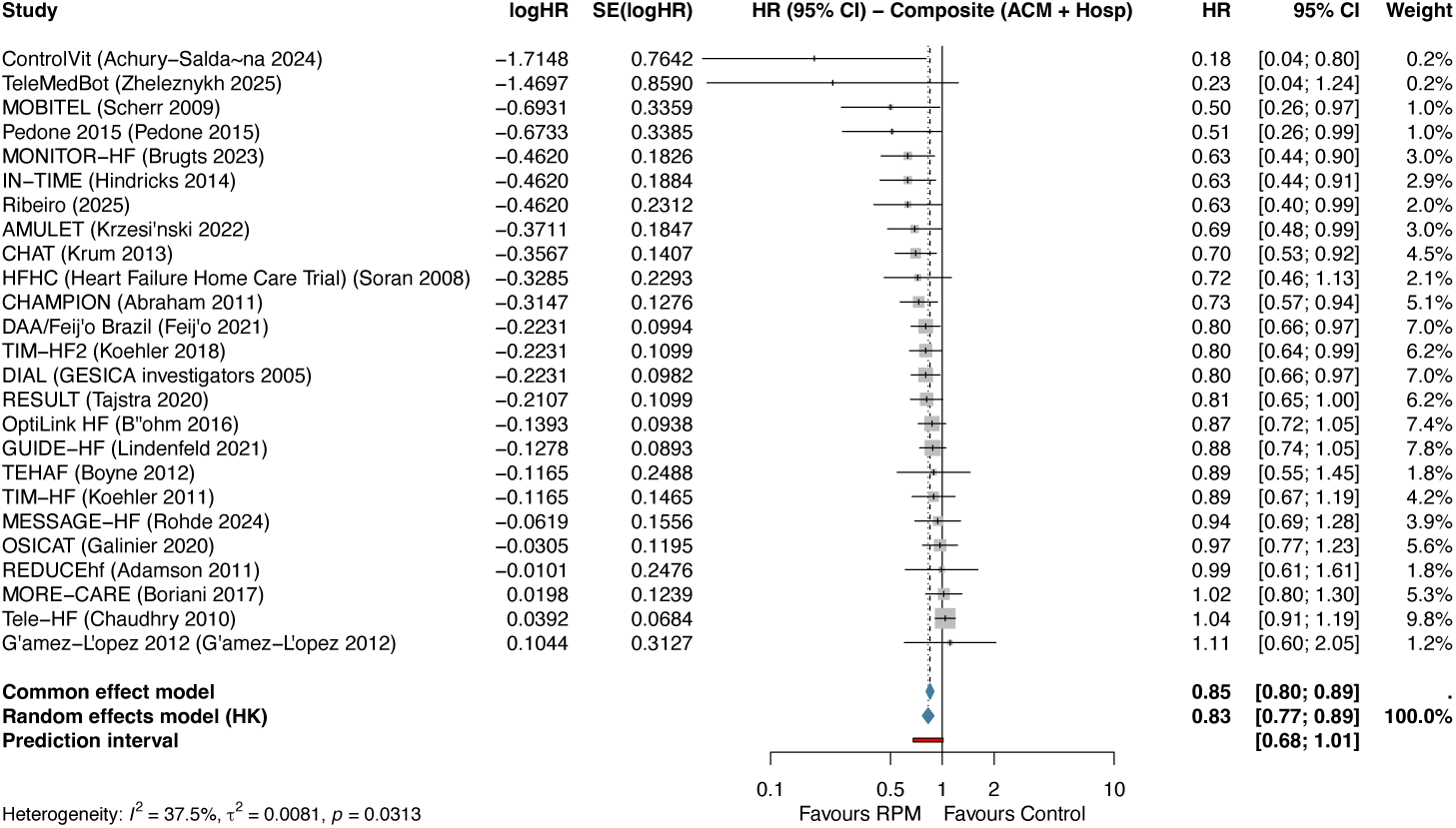
Forest plot of composite all-cause mortality and hospitalization. Pooled HR 0.827 (95% CI 0.765–0.894; I^2^=37.5%; k=25).

**Figure 6.**
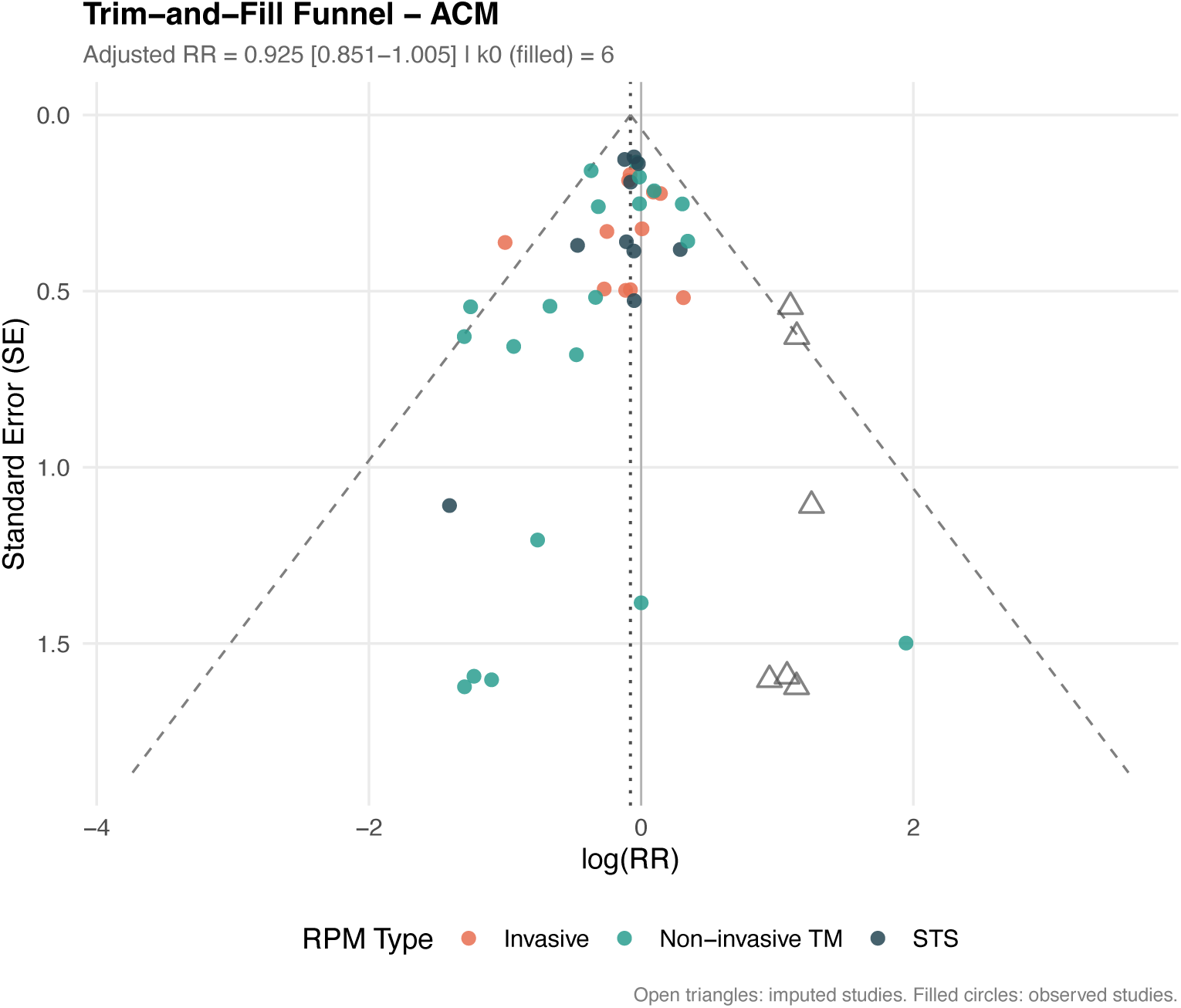
Funnel plot with trim-and-fill analysis for all-cause mortality. Open circles represent observed studies; filled circles represent imputed studies (k_0_=6). Adjusted RR 0.925 (95% CI 0.851–1.005).

For HF hospitalization, all three tests were significant (Egger P<0.001, Begg P<0.001, Peters P=0.0002), with trim-and-fill estimating 12 missing studies and an adjusted RR of 0.830 (95% CI 0.737–0.934; P=0.003). The substantial asymmetry was consistent with small-study effects and potentially greater treatment effects in smaller, more intensive interventions.

### Trial Sequential Analysis

For all-cause mortality with RRR=15%, the accrued information size (AIS=18,738 patients) exceeded the sequentially adjusted required information size (9,880 patients), and the cumulative Z-curve crossed the O’Brien-Fleming monitoring boundary, supporting a stable mortality benefit under this assumption (Figure 8). For HF hospitalization, the AIS did not reach the heterogeneity-adjusted benchmark at RRR=15%, but crossed the monitoring boundary at RRR=20%, indicating that the TSA result is assumption-dependent.

**Figure 7.**
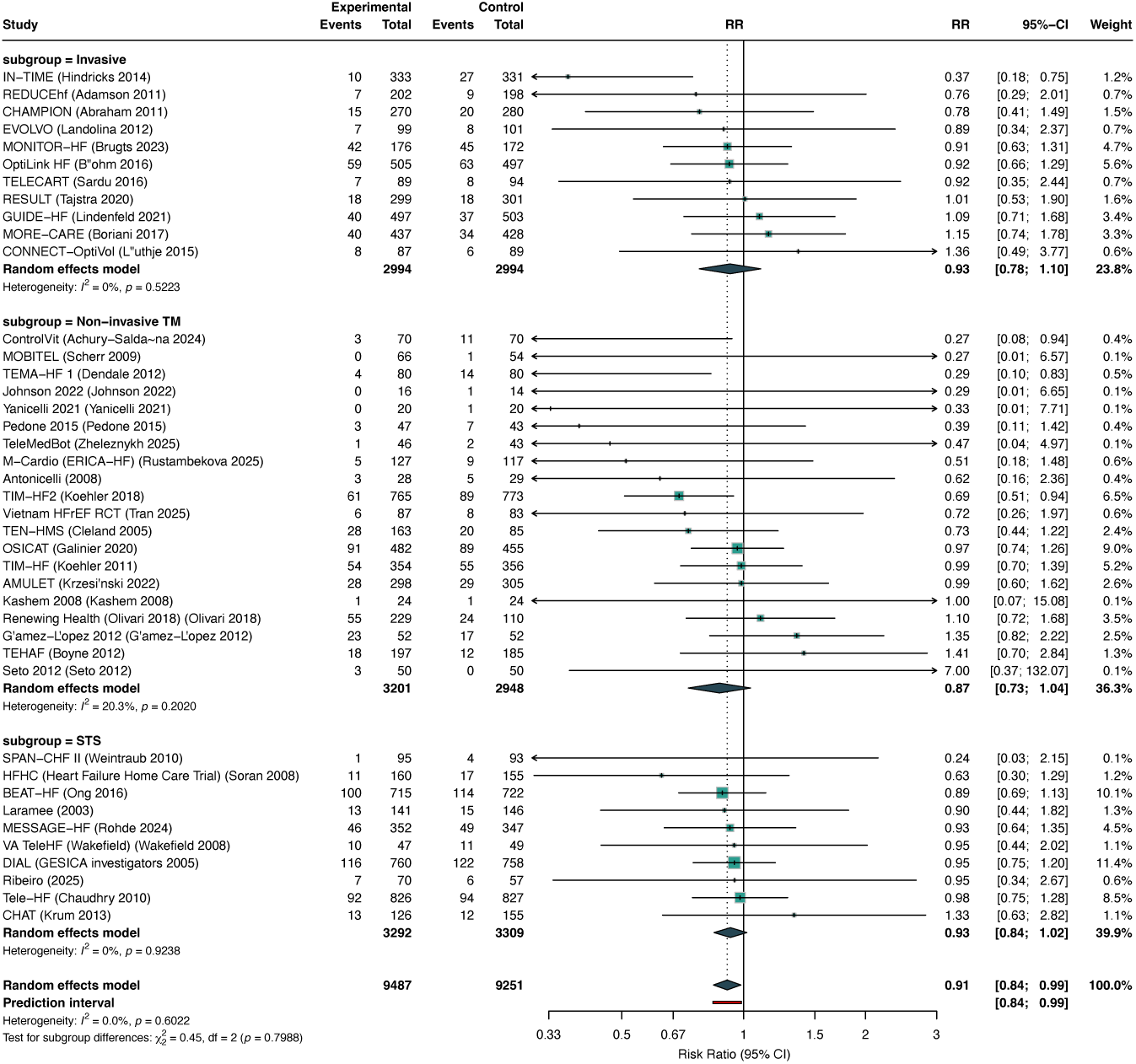
Subgroup analysis of all-cause mortality by RPM type. No significant interaction was observed (P_interaction_=0.80). STS: structured telephone support; TM: non-invasive telemonitoring.

**Figure 8.**
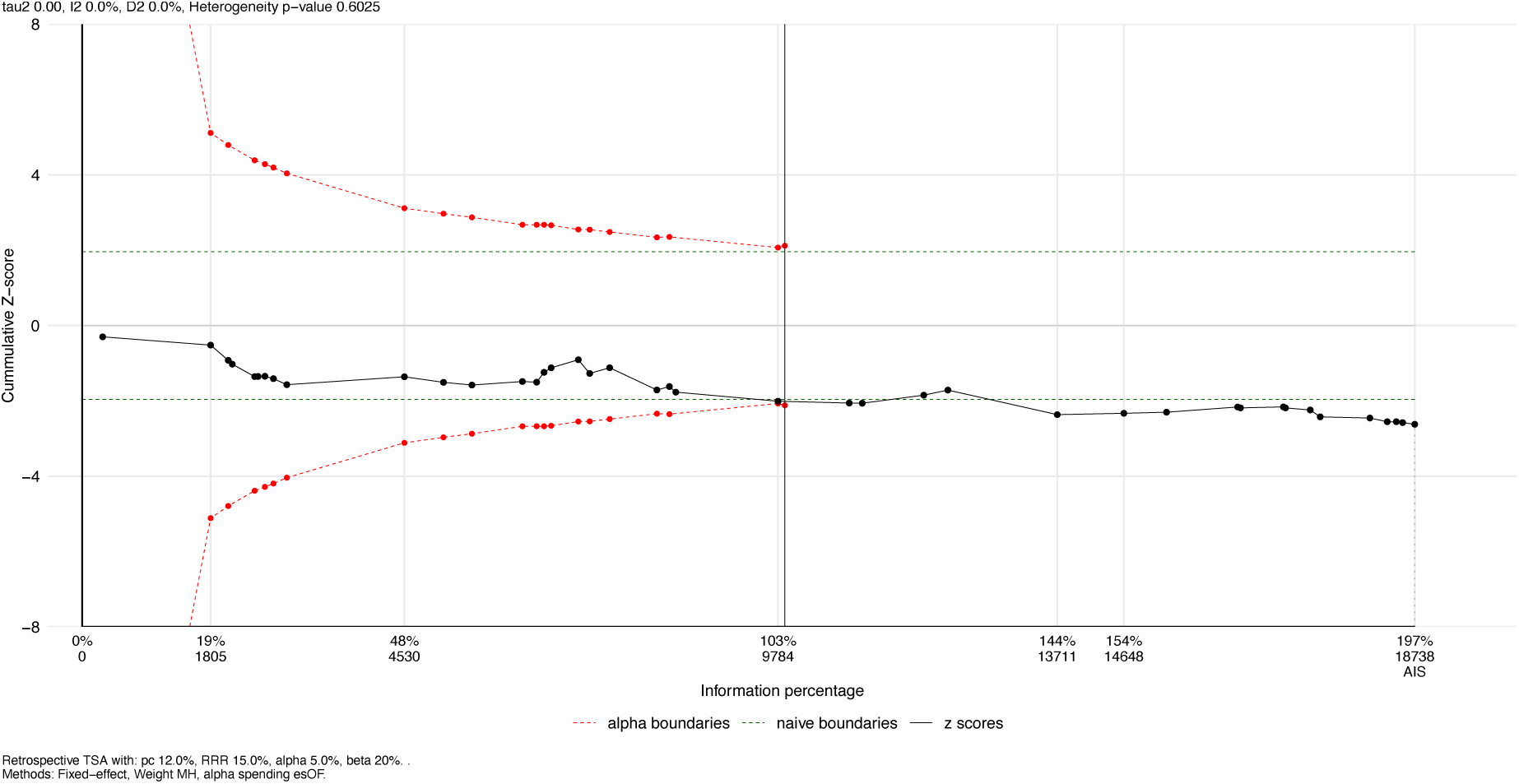
Trial sequential analysis for all-cause mortality (RRR=15%, 𝛼=0.05, 𝛽=0.20, O’Brien-Fleming boundaries). The cumulative Z-curve (blue) crosses the monitoring boundary, and the accrued information size (18,738) exceeds the sequentially adjusted required information size (9,880), supporting a stable mortality benefit under this assumption.

### NNT and GRADE

For all-cause mortality, the NNT was 93 overall and 104 per year (based on a control event rate of 12.0% over a median follow-up of 13.5 months among studies contributing valid participant-level event counts). For HF hospitalization, the NNT was 17 overall and 18 per year (control event rate 26.9%, median follow-up 13.2 months), after excluding two recurrent-count rows from CER/NNT calculation. NNT annualization assumes a constant hazard rate, which may overestimate annual benefit for outcomes with non-proportional hazards; these estimates should be interpreted with this caveat. Furthermore, the prediction interval for HF hospitalization crosses 1.0, meaning that in some clinical settings the NNT could be substantially larger or even infinite. GRADE certainty was moderate for all-cause mortality (downgraded for suspected publication bias), low for HF hospitalization (downgraded for inconsistency and suspected publication bias), very low for all-cause hospitalization, low for cardiovascular mortality (downgraded for very serious imprecision: borderline significance, small k=9, optimal information size not met), and low for the composite outcome (downgraded for indirectness due to mixed effect measures and inconsistency) (eTable 3).

## Discussion

This systematic review and meta-analysis — among the most comprehensive to date on RPM in HF, encompassing 59 poolable RCTs — demonstrates that RPM significantly reduces all-cause mortality by 9% (RR 0.911; NNT 104 per year) and HF hospitalization by 22% (RR 0.781; NNT 18 per year), with no significant effect on all-cause hospitalization. These findings were directionally robust across 10 sensitivity analyses, while publication-bias and small-study signals require cautious interpretation of the headline effects.

The mortality reduction is statistically consistent (I^2^=0%), with a narrow prediction interval (0.840–0.988). This statistical homogeneity should not be overinterpreted as clinical interchangeability across RPM modalities, eras, and healthcare systems. The HKSJ method provides more reliable confidence interval coverage than the DerSimonian-Laird approach, particularly with a moderate number of studies [20,21].

Although no statistically significant interaction by RPM technology type was detected, the analysis had limited power to detect moderate differences given the subgroup sample sizes. Ezimoha et al. reported a significant RPM-type interaction (P=0.007) in a smaller analysis (k=15) [16]. The non-significant interaction in our larger analysis should not be interpreted as proof of equivalence among technologies. Nonetheless, the directional consistency across all three RPM categories suggests that the mechanism of benefit may relate less to any single device and more to the clinical feedback loop that RPM enables — the systematic detection of deterioration, timely clinical response, and guided self-management. This interpretation aligns with the TIM-HF2 rationale, where the structured management protocol was considered as important as the monitoring technology [5].

Our results update and extend the Cochrane reviews by Inglis et al., which reported STS mortality RR 0.87 (95% CI 0.77–0.98) and TM mortality RR 0.80 (95% CI 0.68–0.94) in 2015 [4]. Our overall RR of 0.911 is broadly consistent with but less favorable than these earlier estimates, reflecting the inclusion of several large neutral trials published since 2015 (BEAT-HF, OSICAT, PRADOC, MESSAGE-HF) and the dilution of earlier positive signals by more pragmatic, larger-scale designs. With an NNT of 104 per year for mortality and 18 per year for HF hospitalization, the absolute benefit remains potentially meaningful, but the HF hospitalization estimate should be interpreted alongside the large-trial sensitivity and publication-bias analyses.

Three contemporary meta-analyses published in 2025 provide relevant context. De Lathauwer et al. pooled 41 RCTs (16,312 patients) with access to Embase and reported an OR of 0.81 (95% CI 0.69–0.95) for mortality, directionally consistent with our RR of 0.911 [14]. Their component analysis identified video communication as an effectiveness enhancer — a finding our data could not evaluate but that warrants future investigation. Dobre et al. reported 105 studies (45,072 patients) across multiple databases [15]; however, their substantially larger yield reflects inclusion of non-randomized designs and post-hoc analyses, whereas our review was restricted to RCTs with extractable outcome data. Ezimoha et al. (15 RCTs) reported a significant RPM-type interaction for mortality (P=0.007) [16], contrasting with our non-significant interaction; however, their smaller sample (k=15 vs our k=41) and different subgroup definitions limit direct comparison. The concordance of the mortality benefit direction across independent reviews strengthens confidence in the overall signal, although the exact effect size remains sensitive to analytic choices and small-study effects.

The invasive monitoring evidence deserves specific commentary. The CHAMPION trial showed a striking 28% reduction in HF hospitalization with CardioMEMS-guided management [6], subsequently replicated in MONITOR-HF (HR 0.56) [7]. However, the GUIDE-HF trial yielded neutral overall results (HR 0.88, P=0.16), with benefit confined to a pre-COVID sensitivity analysis [8] — the pandemic reduced event rates in both arms, narrowing the between-group difference. The MORE-CARE trial, which evaluated a different mechanism (remote monitoring of CRT-D device diagnostics rather than direct hemodynamic measurement), was similarly neutral. When pooled, invasive monitoring did not significantly reduce all-cause mortality (RR 0.925, P=0.35), though HF hospitalization was significantly reduced (RR 0.846, P=0.006). This pattern is consistent with the mechanism of action: hemodynamic monitoring primarily targets congestion (the driver of hospitalization) rather than arrhythmia or pump failure (drivers of mortality).

IN-TIME warrants specific discussion as a potential outlier. This trial of Biotronik Home Monitoring using ICD/CRT-D devices reported an extreme mortality hazard ratio of 0.36 (10 vs 27 deaths; n=664) [30]. Leave-one-out analysis demonstrated that removing any single trial shifted the pooled all-cause mortality RR within a narrow range (0.901–0.928), confirming that no single trial, including IN-TIME, disproportionately drives the overall mortality conclusion.

HF phenotype showed limited interaction effects. The predominance of HFrEF or mixed populations in the included trials reflects the historical focus of RPM research. None of the 59 poolable trials enrolled HFpEF patients exclusively; PROACTIVE-HF, which targeted HFpEF, was excluded from quantitative synthesis owing to its feasibility design without a usual-care control arm for the primary endpoint. The significant interaction for all-cause hospitalization by HF phenotype (P=0.002) should be interpreted cautiously given the small number of subgroup-specific trials and the potential for confounding by intervention type and era.

Geographic access was examined as a secondary descriptive analysis, motivated by the expectation that RPM could provide the greatest incremental benefit in populations with limited access to in-person HF care. The findings were informative primarily as a reporting gap. Only two of 59 poolable trials reported formal rural/urban subgroup analyses: TIM-HF2 found no significant interaction (P=0.66), and CHAT was specifically designed for rural and remote patients. Laramee et al. (2003) reported substantial differences between rural (2% HF rehospitalization) and urban (14%) subgroups, but this was a single-center study. The overwhelming majority of trials did not report geographic context at all, making patient-level geographic effect modification infeasible. If RPM is to be deployed as a strategy to address healthcare disparities, trials must be designed and powered to detect geographic access interactions.

The publication bias findings warrant cautious interpretation. Both Egger’s (P=0.040) and Begg’s (P=0.014) tests were significant for all-cause mortality, while Peters’ regression test was non-significant (P=0.246). Trim-and-fill analysis imputed 6 studies, yielding an adjusted RR of 0.925 (95% CI 0.851–1.005; P=0.064), attenuating the mortality estimate to borderline/non-significance. The large-trial sensitivity analysis (N$$200; k=24) yielded RR 0.918 (P=0.033), demonstrating that the mortality signal persists when small studies are excluded, though with a smaller effect size. The contour-enhanced funnel plots suggest that asymmetry is driven primarily by small studies in significance regions, consistent with small-study effects, but selective non-publication cannot be excluded. For HF hospitalization, the publication bias signal was more pronounced: all three tests were significant (Egger P<0.001, Begg P<0.001, Peters P=0.0002), and trim-and-fill imputed 12 studies (31% of k=39), yielding an adjusted RR of 0.830. This degree of asymmetry warrants caution in interpreting the HF hospitalization estimate.

The large-trial sensitivity complements the publication bias analysis and suggests that the HF hospitalization benefit may be closer to the attenuated large-trial estimate (RR 0.837, sensitivity #10) than the overall pooled estimate (RR 0.781) in pragmatic settings. In multivariable meta-regression, total sample size was borderline (P=0.063) rather than statistically significant after excluding recurrent-count rows from binary pooling. No covariates were significant predictors for all-cause mortality, consistent with the low statistical heterogeneity (I^2^=0%) observed for this outcome.

This review has several strengths. It is among the most comprehensive RPM meta-analyses of RCTs, including 59 poolable trials across all three RPM categories. The search encompassed four databases without language restrictions. The analytic approach used REML+HKSJ, included 10 sensitivity analyses per outcome, applied TSA as a sequential-monitoring sensitivity framework, and used GRADE to rate certainty. The geographic access analysis highlights an important reporting gap.

Several limitations should be acknowledged. First, the open-label design inherent to RPM trials introduces potential performance and detection bias, though hard endpoints (mortality, hospitalization) are less susceptible to this bias than patient-reported outcomes. Second, heterogeneity was moderate for HF hospitalization (I^2^=47.2%) and all-cause hospitalization (I^2^=54.3%), likely reflecting variation in intervention intensity, healthcare system context, and patient populations across decades of research. Third, geographic access data were sparse, limiting interpretation of this secondary descriptive analysis. Fourth, some older trials had small sample sizes, contributing to the small-study effects detected in publication bias analysis. Fifth, the mix of effect measures (RR from event counts, HR from time-to-event analyses) required parallel sensitivity analyses rather than a single unified model; additionally, recurrent-event counts could not be pooled as binary participant-level risks and were excluded from RR/NNT calculations. Sixth, the median follow-up of 6 months may not capture longer-term effects of sustained RPM programs. Seventh, the search was limited to four databases (PubMed/MEDLINE, Cochrane CENTRAL, ClinicalTrials.gov, and WHO ICTRP); Embase and CINAHL were not searched due to absence of institutional access, a recognized deviation from Cochrane Handbook recommendations [19]. However, several factors mitigate the potential impact of this omission. Cochrane CENTRAL partially aggregates Embase-indexed content: 42.9% of the 2,138 CENTRAL records retrieved in our search carried an Embase identifier, and Sampson et al. found that CENTRAL captures a growing proportion of Embase RCTs [13]. Slobogean et al. demonstrated that PubMed combined with CENTRAL identifies approximately 97% of relevant RCTs in therapeutic systematic reviews [12]. Of our 65 included studies, 63 (96.9%) had PubMed identifiers, suggesting minimal unique Embase-only yield for this topic. Furthermore, cross-referencing our study list against three contemporary meta-analyses that searched Embase (De Lathauwer et al., 41 RCTs; Dobre et al., 105 studies; Ezimoha et al., 15 RCTs) [14–16] revealed concordant mortality direction across reviews (eText 1). Nevertheless, the possibility that Embase-only European or Asian RCTs were missed cannot be excluded, and this limitation is reflected in the GRADE publication bias domain assessment. Eighth, screening used semi-automated rules with manual adjudication and audit, and archived files do not preserve complete independent reviewer-level decisions. Ninth, although extraction was completed using a two-reviewer consensus process, the archived dataset retains final consensus values rather than a complete row-level history of all initial reviewer disagreements.

### Clinical Implications

The NNT of 18 per year for HF hospitalization, while not directly comparable due to differences in patient populations, outcome definitions, and follow-up durations, is in a broadly similar range to NNTs reported for SGLT2 inhibitors in DAPA-HF and EMPEROR-Reduced (∼19–21/year for the composite of CV death and HF hospitalization) and mineralocorticoid receptor antagonists in EMPHASIS-HF (∼22/year). Combined with the mortality benefit (NNT 104/year), these results support considering RPM programs as an adjunct to guideline-directed medical therapy in HF, particularly where clinical response pathways are well defined. The absence of statistically significant differential effect by RPM type — while acknowledging the limited power to detect moderate differences — suggests that healthcare systems can consider selecting the technology most appropriate to their resources and infrastructure — an important consideration for low- and middle-income countries and for rural health systems where invasive monitoring may not be feasible.

Future research should prioritize geographic access-enriched trials to determine whether RPM differentially benefits patients in rural and underserved settings, HFpEF-specific RPM trials (currently underrepresented), cost-effectiveness analyses comparing RPM modalities, and longer-term follow-up studies to assess sustainability of benefit. Given the cumulative mortality signal, future work may be more informative as comparative effectiveness and implementation research than as simple RPM-versus-no-monitoring trials.

## Conclusions

In this systematic review, meta-analysis, and trial sequential analysis of 59 poolable RCTs encompassing approximately 23,000 patients, remote patient monitoring significantly reduced all-cause mortality (RR 0.911, 95% CI 0.842–0.985; I^2^=0%; GRADE moderate certainty), with a prediction interval (0.840–0.988) that excludes the null. Trial sequential analysis supported a stable mortality signal under the prespecified 15% relative risk reduction assumption, although publication-bias sensitivity attenuated the estimate. HF hospitalization was significantly reduced (RR 0.781, 95% CI 0.710–0.859; NNT 18/year; GRADE low certainty), though the prediction interval (0.586–1.040) indicates that in some contexts the effect may be attenuated or absent, consistent with the moderate heterogeneity observed. No statistically significant differential effect by RPM type was detected. Geographic access context remains a reporting gap: only 2 of 59 poolable trials reported formal rural/urban subgroups, and dedicated geographic access-enriched trials are needed to determine whether RPM can address disparities in HF care delivery.

## Data Availability

All data, analysis code, and materials necessary to reproduce the findings are publicly available in the Zenodo repository (DOI: 10.5281/zenodo.19865261).

