## supplement for "Remote Patient Monitoring in Heart Failure: A Systematic Review, Meta-Analysis, and Trial Sequential Analysis"

#### eTable 1. Search Strategies

##### PubMed/MEDLINE (executed February 15, 2026; 1,387 records)

```
(("telemedicine"[MeSH] OR "remote consultation"[MeSH]

  OR "monitoring, physiologic"[MeSH]

  OR "telemonitor*"[tiab] OR "tele-monitor*"[tiab]

  OR "remote monitor*"[tiab] OR "telehealth"[tiab]

  OR "telemedicine"[tiab] OR "mHealth"[tiab]

  OR "mobile health"[tiab]

  OR "structured telephone support"[tiab]

  OR "telephone follow*"[tiab] OR "phone follow*"[tiab]

  OR "home monitor*"[tiab] OR "home telecare"[tiab]

  OR "digital health"[tiab]

  OR "implantable hemodynamic"[tiab]

  OR "CardioMEMS"[tiab]

  OR "pulmonary artery pressure monitor*"[tiab]

  OR "wearable*"[tiab] OR "smartphone*"[tiab]

  OR "app-based"[tiab])

AND

 ("heart failure"[MeSH] OR "heart failure"[tiab]

  OR "cardiac failure"[tiab] OR "HFrEF"[tiab]

  OR "HFpEF"[tiab] OR "cardiomyopathy"[tiab])

AND

 ("randomized controlled trial"[pt]

  OR "controlled clinical trial"[pt]
```

```
  OR "randomized"[tiab]))
NOT (animals[mh] NOT humans[mh])
```

No date or language restrictions. MeSH Terms used for primary concepts with free-text variants in title/abstract.

#### Cochrane CENTRAL (executed February 15, 2026; 2,138 records)

```
#1  [mh "Telemedicine"]
#2  [mh "Remote Consultation"]
#3  [mh "Monitoring, Physiologic"]
#4  (telemonitor* OR tele-monitor*
    OR (remote NEXT monitor*) OR telehealth
    OR telemedicine):ti,ab,kw
#5  (mHealth OR (mobile NEXT health)
    OR (structured NEXT telephone NEXT support)
    OR (telephone NEXT follow*)):ti,ab,kw
#6  ((home NEXT monitor*) OR (home NEXT telecare)
    OR (digital NEXT health)):ti,ab,kw
#7  (CardioMEMS OR (implantable NEXT hemodynamic)
    OR (pulmonary NEXT artery NEXT pressure
    NEXT monitor*)):ti,ab,kw
#8  (wearable* OR smartphone* OR app-based):ti,ab,kw
#9  #1 OR #2 OR #3 OR #4 OR #5 OR #6 OR #7 OR #8
#10 [mh "Heart Failure"]
#11 ((heart NEXT failure) OR HFrEF OR HFpEF
    OR cardiomyopathy):ti,ab,kw
```

```
#12 #10 OR #11

#13 #9 AND #12
```

Filter: Cochrane CENTRAL (Trials) only.

##### ClinicalTrials.gov (executed February 15, 2026; 348 records)

```
AREA[ConditionSearch] "Heart Failure"

AND AREA[InterventionSearch]

  (telemonitoring OR telehealth

   OR "remote monitoring"

   OR "structured telephone support"

   OR "telephone follow-up"

   OR CardioMEMS

   OR "pulmonary artery pressure"

   OR "home monitoring"

   OR mHealth OR wearable

   OR smartphone OR "digital health")

AND AREA[StudyType] INTERVENTIONAL
```

No date or status restrictions.

##### WHO ICTRP (executed February 15, 2026; 164 records)

Multiple searches combined:

- Search 1: Title: telemonitoring AND heart failure
- Search 2: Title: telehealth AND heart failure
- Search 3: Title: remote monitoring AND heart failure

- Search 4: Title: CardioMEMS OR pulmonary artery pressure; Condition: heart failure

All searches: Recruitment status = ALL; no date restrictions.

### eTable 2. Risk of Bias 2 Detailed Domain Assessments

*eTable 2. Risk of Bias 2 Detailed Domain Assessments*

| Study | D1: Randomisation | D2: Deviations | D3: Missing data | D4: Measurement | D5: Selection | Overall |
|---|---|---|---|---|---|---|
| BEAT-HF (Ong 2016) | Low | Low | Low | Low | Low | Low |
| AMULET (Krzesiński 2022) | Low | Some concerns | Low | Low | Low | Some concerns |
| Antonicelli (2008) | Some concerns | Some concerns | Low | Some concerns | Some concerns | Some concerns |
| Arnar 2025 (Arnar 2025) | Low | Some concerns | Some concerns | Low | Low | Some concerns |
| Benatar (2003) | Low | Some concerns | Low | Low | Some concerns | Some concerns |
| CHAMPION (Abraham 2011) | Low | Some concerns | Low | Low | Low | Some concerns |
| CHAT (Krum 2013) | Some concerns | Some concerns | Low | Low | Low | Some concerns |
| ControlVit (Achury-Saldaña 2024) | Some concerns | Some concerns | Low | Low | Low | Some concerns |
| DAA/Feijó Brazil (Feijó 2021) | Low | Some concerns | Low | Low | Low | Some concerns |
| DIAL (GESICA investigators 2005) | Low | Some concerns | Low | Low | Low | Some concerns |
| Delaney 2013 (Delaney 2013) | Low | Some concerns | Low | Some concerns | Some concerns | Some concerns |
| EVOLVO (Landolina 2012) | Low | Some concerns | Low | Some concerns | Low | Some concerns |
| GUIDE-HF (Lindenfeld 2021) | Low | Some concerns | Some concerns | Low | Low | Some concerns |
| Gámez-López 2012 (Gámez-López 2012) | Some concerns | Some concerns | Some concerns | Low | Some concerns | Some concerns |
| HFHC (Heart Failure Home Care Trial) (Soran 2008) | Low | Some concerns | Low | Low | Some concerns | Some concerns |
| Home-HF (Dar 2009) | Low | Some concerns | Low | Some concerns | Some concerns | Some concerns |
| IN-TIME (Hindricks 2014) | Low | Some concerns | Low | Some concerns | Low | Some concerns |
| Kashem 2008 (Kashem 2008) | Some concerns | Some concerns | Low | Some concerns | Some concerns | Some concerns |
| Laramee (2003) | Some concerns | Some concerns | Some concerns | Low | Some concerns | Some concerns |
| M-Cardio (ERICA-HF) (Rustambekova 2025) | Low | Some concerns | Low | Some concerns | Some concerns | Some concerns |
| MESSAGE-HF (Rohde 2024) | Low | Some concerns | Low | Low | Low | Some concerns |

| Study | D1: Randomisation | D2: Deviations | D3: Missing data | D4: Measurement | D5: Selection | Overall |
|---|---|---|---|---|---|---|
| MIGHTY-HEART (Masterson Creber 2025) | Low | Some concerns | Some concerns | Low | Low | Some concerns |
| MONITOR-HF (Brugts 2023) | Low | High | Low | Low | Low | Some concerns |
| MORE-CARE (Boriani 2017) | Low | Some concerns | Some concerns | Low | Low | Some concerns |
| Negarandeh 2019 (Negarandeh 2019) | Some concerns | Some concerns | Low | Low | Some concerns | Some concerns |
| OSICAT (Galinier 2020) | Low | Some concerns | Some concerns | Low | Low | Some concerns |
| OptiLink HF (Böhm 2016) | Low | Some concerns | Low | Low | Low | Some concerns |
| PRADOC (Roubille 2024) | Low | Some concerns | Low | Some concerns | Low | Some concerns |
| Pekmezaris 2019 (Pekmezaris 2019) | Low | Some concerns | Some concerns | Low | Low | Some concerns |
| REDUCEhf (Adamson 2011) | Low | Some concerns | Low | Low | Some concerns | Some concerns |
| REMOTE-CIED (Versteeg 2019) | Low | Some concerns | Some concerns | Some concerns | Low | Some concerns |
| RESULT (Tajstra 2020) | Low | Some concerns | Low | Low | Low | Some concerns |
| Renewing Health (Olivari 2018) (Olivari 2018) | Low | Some concerns | Low | Low | Low | Some concerns |
| Ribeiro (2025) | Low | Some concerns | Low | Low | Low | Some concerns |
| Riegel (2002) | Some concerns | Some concerns | Some concerns | Low | Low | Some concerns |
| SPAN-CHF II (Weintraub 2010) | Low | Some concerns | Low | Low | Low | Some concerns |
| Seto 2012 (Seto 2012) | Low | Low | Some concerns | Some concerns | Low | Some concerns |
| TEHAF (Boyne 2012) | Low | Some concerns | Some concerns | Low | Low | Some concerns |
| TELECART (Sardu 2016) | Low | Some concerns | Low | Low | Low | Some concerns |
| TEMA-HF 1 (Dendale 2012) | Low | Some concerns | Low | Low | Low | Some concerns |
| TEN-HMS (Cleland 2005) | Low | Low | Low | Low | Some concerns | Some concerns |
| TIM-HF (Koehler 2011) | Low | Some concerns | Low | Low | Low | Some concerns |
| TIM-HF2 (Koehler 2018) | Low | Some concerns | Low | Low | Low | Some concerns |
| Tele-HF (Chaudhry 2010) | Low | Some concerns | Low | Low | Low | Some concerns |

| Study | D1: Randomisation | D2: Deviations | D3: Missing data | D4: Measurement | D5: Selection | Overall |
|---|---|---|---|---|---|---|
| Tompkins 2010 (Tompkins 2010) | Low | Some concerns | Some concerns | Low | Some concerns | Some concerns |
| VA TeleHF (Wakefield) (Wakefield 2008) | Low | Some concerns | Low | Some concerns | Low | Some concerns |
| VERDICT (Copeland VA) (Copeland 2010) | Some concerns | Some concerns | Some concerns | Low | Low | Some concerns |
| Vietnam HFrEF RCT (Tran 2025) | Low | Some concerns | Some concerns | Low | Some concerns | Some concerns |
| Wakefield 2009 (3-arm) (Wakefield 2009) | Low | Some concerns | Some concerns | Low | Some concerns | Some concerns |
| CARME (Domingo 2011) | High | High | Some concerns | Some concerns | Some concerns | High |
| CONNECT-OptiVol (Lüthje 2015) | Low | High | Low | Low | Low | High |
| Dansky 2008 (Dansky 2008) | Some concerns | Some concerns | Some concerns | Low | Some concerns | High |
| Dansky 2009 (Health Buddy) (Dansky 2009) | High | Some concerns | Some concerns | High | Some concerns | High |
| Giordano (2009) | Unclear | Unclear | Unclear | Low | Unclear | High |
| HHH (Mortara 2009) | Some concerns | High | Some concerns | Low | Some concerns | High |
| Jerant 2001 (3-arm) (Jerant 2001) | Some concerns | Some concerns | Some concerns | Low | Some concerns | High |
| Jerant 2003 (3-arm) (Jerant 2003) | Some concerns | Some concerns | Some concerns | Some concerns | Some concerns | High |
| Johnson 2022 (Johnson 2022) | Low | Some concerns | Some concerns | Low | Some concerns | High |
| MOBITEL (Scherr 2009) | Some concerns | High | Some concerns | Low | Some concerns | High |
| Ohio Telecare (Madigan) (Madigan 2013) | Low | Some concerns | Some concerns | Some concerns | Some concerns | High |
| PROACTIVE-HF (Guichard 2024) | High | Low | Low | Low | Low | High |
| Pedone 2015 (Pedone 2015) | Some concerns | Some concerns | High | Some concerns | Low | High |
| THCM (Wade 2011) (Wade 2011) | High | High | High | Some concerns | Low | High |
| TeleMedBot (Zheleznykh 2025) | Some concerns | High | High | Some concerns | Some concerns | High |
| Yanicelli 2021 (Yanicelli 2021) | Some concerns | Some concerns | High | Some concerns | Low | High |

D1: Randomisation process; D2: Deviations from intended interventions; D3: Missing outcome data; D4: Measurement of the outcome; D5: Selection of the reported result. All RPM trials are inherently open-label; D2 ratings reflect whether knowledge of assignment influenced co-interventions beyond the intended effect.

### eTable 3. GRADE Evidence Profile

*eTable 3. GRADE Evidence Profile*

| outcome | k_studies | design | starting_certainty | rob_rating | inconsistency_rating | i2_pct | indirectness_rating | imprecision_rating | pub_bias_rating | total_downgrade | certainty | effect_size | p_value | notes | total_downgrades |
|---|---|---|---|---|---|---|---|---|---|---|---|---|---|---|---|
| All-Cause Mortality | 41 | RCT | High | Not serious | Not serious | 0.0 | Not serious | Not serious | Suspected | -1 | Moderate | RR 0.911 [0.842–0.985] | 0.0208 | | |
| HF Hospitalization | 39 | RCT | High | Not serious | Serious | 47.2 | Not serious | Not serious | Suspected | -2 | Low | RR 0.781 [0.710–0.859] | 0.0000 | PI crosses null | |
| All-Cause Hospitalization | 28 | RCT | High | Not serious | Serious | 54.3 | Not serious | Serious | Suspected | -3 | Very Low | RR 0.959 [0.892–1.031] | 0.2462 | PI crosses null | |
| CV Mortality | 9 | RCT | High | Not serious | Not serious | 0.0 | Not serious | Very serious | Undetected | -1 | Low | RR 0.814 [0.668–0.992] | 0.0433 | ; Very serious imprecision: borderline significance (P=0.043), small k=9, optimal information size not met | -2 |
| Composite ACM+Hospitalization | 25 | RCT | High | Not serious | Serious | 37.5 | Serious | Not serious | Undetected | 0 | Low | RR 0.827 [0.765–0.894] | 0.0000 | PI crosses null; Mixed HR/OR/RR; log-scale pooling; Downgraded: indirectness (mixed HR/OR/RR effect | -2 |

| outcome | k_studies | design | starting_certainty | rob_rating | inconsistency_rating | i2_pct | indirectness_rating | imprecision_rating | pub_bias_rating | total_downgrade | certainty | effect_size | p_value | notes | total_downgrades |
|---|---|---|---|---|---|---|---|---|---|---|---|---|---|---|---|
| | | | | | | | | | | | | | | measures pooled on log scale) and inconsistency (I2=37.5% with heterogeneous measures) | |
| MLHFQ | 11 | RCT | High | Not serious | Very serious | 93.6 | Not serious | Not serious | Undetected | -2 | Low | MD -6.53 [-10.28–-2.79] | 0.0030 | Lower = better; high heterogeneity | |
| ED Visits | 6 | RCT | High | Not serious | Serious | 41.4 | Not serious | Serious | Undetected | -2 | Low | RR 0.795 [0.551–1.148] | 0.1699 | PI crosses null; k=6; limited data | |
| KCCQ | 3 | RCT | High | Serious | Very serious | 83.7 | Not serious | Serious | Undetected | -4 | Very Low | MD 8.14 [-24.03–40.30] | 0.3902 | Higher = better; k=3 only | |

*Note: The publication bias domain for all outcomes incorporates the potential impact of the Embase and CINAHL omission. Cross-referencing against three contemporary meta-analyses with Embase access (De Lathauwer et al. 2025, Dobre et al. 2025, Ezimoha et al. 2025) showed concordant effect estimates, supporting the robustness of these ratings (see eText 1).*

### eTable 4. Sensitivity Analysis Results

*eTable 4. Sensitivity Analyses for All Primary Outcomes*

| Outcome | Analysis | k | RR / Range | LCI | UCI | P | I2 (%) | Note |
|---|---|---|---|---|---|---|---|---|
| ACM | 10_large_trials_n200 | 24 | 0.918 | 0.849 | 0.993 | 0.0331 | 0 | N>=200 only (n excl.=17) |
| ACM | 11_incl_active_comparators | 42 | 0.918 | 0.848 | 0.995 | 0.0372 | 0 | Post-hoc: including MIGHTY-HEART active comparator (CO-0570) |
| ACM | 1_leave_one_out | 41 | [0.901–0.928] | — | — | — | — | LOO pooled-RR range [0.901–0.928] across k=41 iterations |
| ACM | 2_fixed_effect | 41 | 0.9 | 0.832 | 0.974 | 0.0087 | 0 | Fixed-effect (common); no HKSJ correction |
| ACM | 3_usual_care_only | 37 | 0.91 | 0.839 | 0.986 | 0.023 | 0 | Comparator type = usual care only (n excl.=4) |
| ACM | 4_excl_invasive | 30 | 0.906 | 0.826 | 0.994 | 0.0382 | 0 | Excl. Invasive RPM (n excl.=11) |
| ACM | 5_excl_cluster_rct | 40 | 0.907 | 0.838 | 0.981 | 0.0165 | 0 | Excl. CHAT (CO-0944) cluster RCT |
| ACM | 6_low_rob_only | 35 | 0.914 | 0.843 | 0.992 | 0.0314 | 0 | Excl. High RoB (n excl.=6) |
| ACM | 7_excl_outliers | 36 | 0.93 | 0.868 | 0.998 | 0.043 | 0 | Excl. flagged outliers (n excl.=5) |
| ACM | 8_hr_only_subset | 16 | 0.88 | 0.784 | 0.989 | 0.0335 | 0 | HR-reporting studies only (metagen); k=16 |
| ACM | 9_fu_ge3mo | 41 | 0.911 | 0.842 | 0.985 | 0.0208 | 0 | FU >=3 months only (n excl.=0) |
| Allcause_hosp | 10_large_trials_n200 | 17 | 0.988 | 0.934 | 1.046 | 0.6702 | 39.4 | N>=200 only (n excl.=12) |
| Allcause_hosp | 11_incl_active_comparators | 29 | 0.965 | 0.903 | 1.031 | 0.2765 | 52.7 | Post-hoc: including MIGHTY-HEART active comparator (CO-0570); excluded recurrent-count rows incompatible with binary RR (n=1) |
| Allcause_hosp | 1_leave_one_out | 28 | [0.948–0.978] | — | — | — | — | LOO pooled-RR range [0.948–0.978] across k=28 iterations |
| Allcause_hosp | 2_fixed_effect | 28 | 0.964 | 0.93 | 1 | 0.048 | 54.3 | Fixed-effect (common); no HKSJ correction |
| Allcause_hosp | 3_usual_care_only | 23 | 0.92 | 0.838 | 1.009 | 0.0745 | 57.1 | Comparator type = usual care only (n excl.=5); excluded recurrent-count rows incompatible with binary RR (n=1) |
| Allcause_hosp | 4_excl_invasive | 25 | 0.931 | 0.851 | 1.018 | 0.1098 | 54.6 | Excl. Invasive RPM (n excl.=3); excluded recurrent-count rows incompatible with binary RR (n=1) |
| Allcause_hosp | 5_excl_cluster_rct | 27 | 0.966 | 0.898 | 1.039 | 0.3402 | 54.5 | Excl. CHAT (CO-0944) cluster RCT; excluded recurrent-count rows incompatible with binary RR (n=1) |
| Allcause_hosp | 6_low_rob_only | 23 | 0.98 | 0.927 | 1.036 | 0.4627 | 39 | Excl. High RoB (n excl.=5); excluded recurrent-count rows incompatible with binary RR (n=1) |
| Allcause_hosp | 7_excl_outliers | 26 | 0.969 | 0.911 | 1.031 | 0.3031 | 44.7 | Excl. flagged outliers (n excl.=3) |
| Allcause_hosp | 8_hr_only_subset | 10 | 0.87 | 0.711 | 1.064 | 0.1514 | 71 | HR-reporting studies only (metagen); k=10 |
| Allcause_hosp | 9_fu_ge3mo | 27 | 0.965 | 0.9 | 1.034 | 0.2992 | 52.8 | FU >=3 months only (n excl.=1); excluded recurrent-count rows incompatible with binary RR (n=1) |
| HF_hosp | 10_large_trials_n200 | 23 | 0.837 | 0.779 | 0.899 | <0.001 | 21.8 | N>=200 only (n excl.=16); excluded recurrent-count rows incompatible with binary RR (n=2) |

| Outcome | Analysis | k | RR / Range | LCI | UCI | P | I2 (%) | Note |
|---|---|---|---|---|---|---|---|---|
| HF_hosp | 11_incl_active_comparators | 40 | 0.79 | 0.721 | 0.865 | <0.001 | 46.7 | Post-hoc: including MIGHTY-HEART active comparator (CO-0570); excluded recurrent-count rows incompatible with binary RR (n=2) |
| HF_hosp | 1_leave_one_out | 39 | [0.771–0.799] | — | — | — | — | LOO pooled-RR range [0.771–0.799] across k=39 iterations |
| HF_hosp | 2_fixed_effect | 39 | 0.805 | 0.763 | 0.85 | <0.001 | 47.2 | Fixed-effect (common); no HKSJ correction |
| HF_hosp | 3_usual_care_only | 33 | 0.793 | 0.723 | 0.87 | <0.001 | 43.7 | Comparator type = usual care only (n excl.=7); excluded recurrent-count rows incompatible with binary RR (n=1) |
| HF_hosp | 4_excl_invasive | 30 | 0.725 | 0.629 | 0.835 | <0.001 | 53 | Excl. Invasive RPM (n excl.=11) |
| HF_hosp | 5_excl_cluster_rct | 38 | 0.779 | 0.706 | 0.859 | <0.001 | 48.6 | Excl. CHAT (CO-0944) cluster RCT; excluded recurrent-count rows incompatible with binary RR (n=2) |
| HF_hosp | 6_low_rob_only | 31 | 0.797 | 0.724 | 0.877 | <0.001 | 46.3 | Excl. High RoB (n excl.=8); excluded recurrent-count rows incompatible with binary RR (n=2) |
| HF_hosp | 7_excl_outliers | 33 | 0.812 | 0.75 | 0.878 | <0.001 | 33.3 | Excl. flagged outliers (n excl.=6); excluded recurrent-count rows incompatible with binary RR (n=2) |
| HF_hosp | 8_hr_only_subset | 13 | 0.766 | 0.698 | 0.841 | <0.001 | 1.1 | HR-reporting studies only (metagen); k=13 |
| HF_hosp | 9_fu_ge3mo | 39 | 0.781 | 0.71 | 0.859 | <0.001 | 47.2 | FU >=3 months only (n excl.=0); excluded recurrent-count rows incompatible with binary RR (n=2) |

### eTable 5. Intervention Characteristics

*eTable 5. Intervention Characteristics by Study*

| Study | RPM Category | Subcategory | Technology/Platform | Monitoring Parameters | Frequency | Who Responds | Alert Threshold | Patient Education | Duration (wk) | Adherence (%) |
|---|---|---|---|---|---|---|---|---|---|---|
| CHAMPION (Abraham 2011) | Invasive | Implanted PA sensor | CardioMEMS PA pressure sensor | PA pressure (systolic, diastolic, mean) | Daily | Cardiologist | Yes | No | 26 | NR |
| REDUCEhf (Adamson 2011) | Invasive | ICD remote (IHM) | Medtronic Chronicle IHM-ICD (implantable hemodynamic monitor + ICD) | RV systolic/diastolic pressure, estimated PAD, dP/dt, heart rate, activity (continuous) | Automatic (weekly upload via telephone) | Cardiologist | Yes | Yes | 52 | NR |
| EVOLVO (Landolina 2012) | Invasive | ICD remote | Medtronic CareLink Network (wireless ICD/CRT-D; full interrogation via telephone; secure web platform; OptiVol intrathoracic impedance) | intrathoracic impedance (OptiVol), AT/AF burden, ICD shocks, ventricular rate, lead impedances, VF detection status, battery, daily activity, HR, HRV | Automatic | Cardiologist | Yes | Yes | 70 | NR |
| IN-TIME (Hindricks 2014) | Invasive | ICD/CRT-D remote monitoring | Remote monitoring (Home Monitoring, ICD/CRT-D) | Arrhythmias (VT/AT), biventricular pacing %, ventricular extrasystole frequency, patient activity trend, intracardiac electrogram, pacing/impedance safety notifications | Daily automatic transmission (typically 03:00h) + on tachyarrhythmia detection | Investigational site physician + central monitoring unit (nurses + physicians at Heart Center Leipzig); patient contact by phone within 48h of alert | Pre-defined: VT/shock, AT onset/burden, CRT <80% over 48h, VES >110/h or rising, activity decline, abnormal IEGM, pacing/impedance alerts, transmission gap >3 days | Standardised telephone interview on contact: symptoms, drug adherence, weight gain >2kg/3 d | 52 | 85% |
| CONNECT-OptiVol (Lüthje 2015) | Invasive | ICD remote | Remote ICD/CRT-D monitoring with OptiVol fluid index | Intrathoracic impedance (OptiVol fluid index); Cardiac Compass; remote transmission via CareLink network | Continuous automatic transmission on alert; otherwise routine remote check | Study investigator team (physician review of transmitted report) | OptiVol threshold set to nominal 60 Ω at implantation (unchanged during follow-up); audible alert DISABLED in both groups | Patient phones investigator on audible alert disabled; remote arm: weight monitoring daily after diuretic increase | 65 | NR |
| OptiLink HF (Böhm 2016) | Invasive | ICD remote | Medtronic OptiVol intrathoracic impedance fluid index + CareLink wireless network (CareAlert automatic text to physician) | Intrathoracic fluid index (OptiVol impedance); ICD device diagnostics | Automatic | Cardiologist | Yes | No | 97 | 76% |
| TELECART (Sardu 2016) | Invasive | CRT-D remote | Biotronik Home Monitoring (Lumax 640HF/Iforia CRT-D) — daily automatic IEGM + arrhythmia telemetry | Device diagnostics: biventricular pacing %, arrhythmia episodes (AF/VT/VF), ICD shocks, intracardiac electrograms, patient activity | Continuous automatic transmission; alerts reviewed on working days; site must confirm within 48h | Central monitoring unit (trained nurses + physicians) at Giovanni Paolo II Foundation; alerts forwarded to investigational site; investigators contact | Predefined medical events: VT/AF episodes, low biventricular pacing %, increased ventricular extrasystoles, decreased patient activity, abnormal IEGM | Patients instructed to monitor weight, dyspnoea, symptoms; telephone interview at alert: weight gain >2kg/3 d, dyspnoea, drug adherence | 52 | NR |

| Study | RPM Category | Subcategory | Technology/Platform | Monitoring Parameters | Frequency | Who Responds | Alert Threshold | Patient Education | Duration (wk) | Adherence (%) |
|---|---|---|---|---|---|---|---|---|---|---|
| | | | | | | patient by telephone | | | | |
| MORE-CARE (Boriani 2017) | Invasive | CRT-D remote | Medtronic CareLink remote monitor (wireless home unit; Medtronic CRT-D with wireless transmission) | Lung fluid accumulation (OptiVol), atrial tachyarrhythmia (AF/flutter), system integrity, ECG/device diagnostics | Automatic (remote checks every 4 months alternating with in-office; automatic alerts continuous) | Cardiologist | Yes | Yes | 96 | 83% |
| RESULT (Tajstra 2020) | Invasive | ICD remote / CRT-D remote | Multi-manufacturer (Carelink/Merlin/LATITUDE/Home Monitoring); centralized remote monitoring office | Device diagnostics (arrhythmia, system integrity, battery, charge time, VF therapy); telephone interview (dyspnea, drug compliance, weight) | Automatic daily transmission | Centralized team (2 physicians + 2 EP nurses) | Yes | Yes | 52 | 86% |
| GUIDE-HF (Lindenfeld 2021) | Invasive | Implanted PA sensor | CardioMEMS PA pressure sensor | PA systolic pressure, PA diastolic pressure, PA mean pressure, heart rate | Daily uploads | Clinician (investigator) | PA pressure elevation triggers diuretic/vasodilator titration per protocol | Yes — scripted calls every 2 weeks (first 3 mo), then monthly; masked caller | 52 | 85% |
| MONITOR-HF (Brugts 2023) | Invasive | Invasive hemodynamic (pulmonary artery pressure) | CardioMEMS PA pressure sensor (Abbott) | Pulmonary artery pressure (daily uploads) | Daily patient uploads (84.3% adherence) | HF nurse/clinician team; dedicated outpatient clinic nurses | PA pressure thresholds — diuretics titrated for hypervolemia/hypovolemia; vasodilators for increased vascular resistance | Yes — patient training on PA pressure uploads | 192 | 84% |
| Jerant 2001 (3-arm) (Jerant 2001) | Non-Invasive TM | mHealth app | 2-way video-conference device with integrated electronic stethoscope | Video clinical assessment, electronic auscultation (heart/lung sounds), symptom review | Variable | Nurse | NR | Yes | 26 | NR |
| Benatar (2003) | Non-Invasive TM | Weight + BP + HR + SpO2 (daily Internet transmission) | Transtelephonic home monitoring devices | Weight, blood pressure, heart rate, oxygen saturation (SpO2) | Daily transmission to secure Internet site | Advanced-practice nurse (APN) collaborating with cardiologist | Clinical guideline-based thresholds for medication titration | Yes — APN telephoned patients, provided education and medication adjustments per HF clinical guidelines | 13 | NR |
| Jerant 2003 (3-arm) (Jerant 2003) | Non-Invasive TM | mHealth app | Video home telecare (2-way videoconference unit) + structured telephone monitoring (telephone arm) | CHF symptoms, weight, self-care adherence, medications, health status; video clinical assessment + electronic stethoscope | Variable | Nurse | NR | Yes | 26 | NR |
| TEN-HMS (Cleland 2005) | Non-Invasive TM | Weight + BP + HR + ECG rhythm (twice-daily | Home Telemonitoring (HTM) — daily BP/weight/ECG; nurse-led telephonic review | Weight, blood pressure, heart rate, single- | Twice daily (before breakfast and before | Specialist nurse (reviews alerts, contacts | Weight change >2 kg; HR <50 or >80 bpm; new | Individualized written management plan; device | 34 | 81% |

| Study | RPM Category | Subcategory | Technology/Platform | Monitoring Parameters | Frequency | Who Responds | Alert Threshold | Patient Education | Duration (wk) | Adherence (%) |
|---|---|---|---|---|---|---|---|---|---|---|
| | | automated transmission) | | lead ECG rhythm | evening meal) | patient/primary care physician) | arrhythmia; SBP <90 or >140 mmHg | installation instruction; nurse available by telephone | | |
| Antonicelli (2008) | Non-Invasive TM | Nurse telephone | A&D UA-767 PC (BP device) + Card-Guard CG-7100 (12-lead transtelephonic ECG, Card Guard Scientific Survival Ltd Israel) | BP, HR, body weight, 24h urine output, ECG (weekly 12-lead transtelephonic), symptoms (dyspnoea, oedema), medication adherence | Weekly | Mixed | Yes | Yes | 52 | 91% |
| Dansky 2008 (Dansky 2008) | Non-Invasive TM | Non-invasive TM | HomMed Health Monitor / ViTel Net (1-way asynchronous) + Aviva (2-way video+stethoscope) | Weight, BP, HR, SpO2, blood glucose (as ordered); symptom assessment (Omaha PRSO: diet/fluid intake, physical activity, medication effectiveness) | Daily (one-way); 2-3x/week (video arm) | Nurse (HHA central station) | Yes | No | 9 | NR |
| Kashem 2008 (Kashem 2008) | Non-Invasive TM | mHealth app | InSight Telehealth system | weight, BP, HR, SpO2, symptoms | Daily | Nurse | Yes | Yes | 52 | NR |
| Dansky 2009 (Health Buddy) (Dansky 2009) | Non-Invasive TM | IVR | Health Buddy interactive device (patient-entered data to care manager) | Daily weight, ankle swelling, shortness of breath (5-item symptom questionnaire) | Daily | Care manager | Yes | Yes | 26 | NR |
| Home-HF (Dar 2009) | Non-Invasive TM | Home device | Honeywell HomMed | Weight, BP, HR, SpO2, 4 symptom questions (breathlessness, orthopnoea, dizziness, ankle swelling) | Daily (Mon-Fri nurse review) | HF nurse | Yes | Yes | 26 | 95% |
| Giordano (2009) | Non-Invasive TM | Portable ECG device + nurse teleconsultation | Portable device transmitting 1-lead ECG trace by telephone | 1-lead ECG trace | NR (abstract does not specify frequency) | Nurse (interactive teleconsultation at receiving station) | NR (abstract does not specify thresholds) | NR | 52 | NR |
| HHH (Mortara 2009) | Non-Invasive TM | Home device (IVR + cardiorespiratory recorder) | NICRAM system (IVR vital signs + portable Holter cardiorespiratory recorder + digital BP + scale) | Weight, HR, systolic BP, dyspnea score, asthenia score, edema score, therapy changes, blood results (weekly); 24h cardiorespiratory recording monthly (Strategies 2+3) | Weekly (vital signs); monthly (telephone + cardiorespiratory) | Monitoring nurse or physician (after automated alert) | Yes | Yes | 48 | 81% |
| MOBITEL (Scherr 2009) | Non-Invasive TM | mHealth app | Nokia 3510 mobile phone + Soehnle creta scale + BosoMedicus BP device + MOBITEL web platform (Zope/Interbase; AIT Graz) | blood pressure, heart rate, body weight, HF medication dose | Daily | Cardiologist | Yes | Yes | 26 | 95% |
| Tompkins 2010 | Non-Invasive TM | Wearable | Vital signs TM (daily to central nursing station) | Weight, blood pressure, heart rate, SpO2, weekly | Daily | Central nursing station | Yes | Yes | 26 | NR |

| Study | RPM Category | Subcategory | Technology/Platform | Monitoring Parameters | Frequency | Who Responds | Alert Threshold | Patient Education | Duration (wk) | Adherence (%) |
|---|---|---|---|---|---|---|---|---|---|---|
| (Tompkins 2010) | | | | symptom prompts | | | | | | |
| TIM-HF (Koehler 2011) | Non-Invasive TM | Wearable | ECG + BP + weight daily; physician-led telemedical centre | ECG (3-lead), blood pressure, body weight | Daily | Physician (24h/7d telemedical centre) | Predefined standard operating procedures; physician contacts patient when threshold triggered or patient-initiated | Device training provided within 5 working days of randomisation | ~113 | 81% |
| THCM (Wade 2011) (Wade 2011) | Non-Invasive TM | Wearable | Internet-connected telemonitoring with nurse case management alerts | weight, BP, medication adherence, symptom questions | Daily (weekdays) | Nurse | Yes | Co-intervention | 26 | 74% |
| TEHAF (Boyne 2012) | Non-Invasive TM | IVR | Health Buddy device (LCD + 4 keys, landline phone); daily pre-set tailored dialogues on symptoms/knowledge/behaviour; no automatic vital sign transfer | Symptoms (HF-related), knowledge, behaviour/adherence; BP and HR collected at face-to-face visits only | Daily (dialogues sent daily) | Heart failure nurse + nurse assistant (reviewed responses on desktop; immediate response to positive symptom answers) | Risk profiles (low/medium/high) generated from responses; positive symptom answers triggered immediate nurse response | Yes — tailored education component (4 programmes varying symptom monitoring + education intensity) | 52 | 90% |
| TEMA-HF 1 (Dendale 2012) | Non-invasive TM | Wearable | Electronic body weight scale + blood pressure monitor + cell phone (Bluetooth to central computer) | weight, blood pressure, heart rate | Daily | GP + HF nurse/specialist (collaborative) | Yes (weight ±2kg, SBP 90-140, HR 50-90 bpm) | Yes | 26 | 83% |
| Gámez-López 2012 (Gámez-López 2012) | Non-Invasive TM | Nurse telephone | Structured telephone support by nurse (monthly calls + symptom-triggered calls) | Symptoms, weight, functional status | Monthly + as needed | Nurse | Yes | Yes | 47 | NR |
| Seto 2012 (Seto 2012) | Non-Invasive TM | mHealth app | Mobile phone with automated wireless transmission + alerts to cardiologist | Daily weight, daily BP, weekly single-lead ECG, daily symptom questionnaire | Daily (weight, BP, symptoms); weekly (single-lead ECG) | Cardiologist (receives email/mobile alerts; contacts patients by phone; adjusts medications and schedules clinic visits) | Clinician-defined per-patient physiological thresholds; low-priority (retake) to high-priority (go to ED/call 911) alerts sent to cardiologist mobile phone | Yes — individual training session on system use; automated instructions sent to patient mobile phone; adherence reminder calls at 10am | 26 | 84% |
| Delaney 2013 (Delaney 2013) | Non-Invasive TM | Automated vital signs monitor | HomMed Health Monitor (Honeywell) — automated vital signs via phone lines; daily transmission to central server | weight, BP, HR, SpO2, symptoms (5 daily questions) | Daily | TM Program manager (daily review) | Yes | Co-intervention | 13 | NR |
| Pedone 2015 (Pedone 2015) | Non-Invasive TM | Non-invasive multiparameter home device | A&D Engineering sphygmomanometer + scale + Nonin Medical pulse oximeter; Android smartphone transmitter; web-based monitoring portal; + geriatrician telephone support | SpO2, heart rate, blood pressure, body weight | Daily (weight 1x/d; BP+HR 2x/d; SpO2 3x/d) | Geriatrician | Yes | Yes | 26 | 62% |
| TIM-HF2 (Koehler 2018) | Non-Invasive TM | Remote Patient Management (RPM) with physician-led telemedical centre | Daily ECG + blood pressure + weight via mobile device | Daily ECG (3-channel, 2min or streaming; PhysioMem PM1000 GETEMED), blood pressure (UA767PBT A&D), body weight (Seca | Daily transmission via mobile phone network (VPN tunnel) to telemedical | Telemedical centre physicians + HF nurses 24/7 Mon-Sun; Fontane software (CE-marked) with risk algorithms; | Risk categorization (low/high) using MR-proADM + transmitted vital signs; reassessed every 3 months; Fontane algorithms triggered alerts | Yes — HF patient education programme initiated at device setup by certified nurses; monthly | 52 | 97% |

| Study | RPM Category | Subcategory | Technology/Platform | Monitoring Parameters | Frequency | Who Responds | Alert Threshold | Patient Education | Duration (wk) | Adherence (%) |
|---|---|---|---|---|---|---|---|---|---|---|
| | | | | 861), SpO2 (Masimo SET), self-rated health status (1-5 scale) | centre at fixed time | patient GP and cardiologist also involved | | structured telephone interviews throughout study | | |
| Renewing Health (Olivari 2018) (Olivari 2018) | Non-Invasive TM | mHealth app | RENEWING HEALTH platform (European CIP ICT PSP project, telemedicine home monitoring system) | NR (standard HF parameters; RENEWING HEALTH platform) | Daily | Mixed | Yes | Co-intervention | 52 | 83% |
| Pekmezaris 2019 (Pekmezaris 2019) | Non-Invasive TM | mHealth app | American TeleCare LifeView (home-installed video telehealth + daily vital signs) | weight, BP, SpO2, HR/pulse | Daily | Nurse | Yes | Co-intervention | 13 | 50% |
| OSICAT (Galinier 2020) | Non-Invasive TM | Wearable | Electronic scale (body weight daily) + 8-symptom questionnaire device (daily); automated expert system generates alerts; secure server | Daily body weight, daily HF symptom questionnaire (8 items), personalised education (phone calls every 3 weeks) | Daily | Nurse (working days only); nurse contacts patient to validate alert; if appropriate advises patient to contact GP/cardiologist; follow-up call 48h later | Automated expert system thresholds (not specified in paper); alerts generated based on weight/symptom data | Yes — personalised info pack + telephone calls every 3 weeks with HF-specialised nurse; topics: disease knowledge, medications, sign recognition, lifestyle (diet+activity) | 78 | 60% |
| Yanicelli 2021 (Yanicelli 2021) | Non-Invasive TM | mHealth app | Custom home telemonitoring app (daily measurements + alerts) | weight, blood pressure, heart rate, symptoms (ankle/leg swelling, dyspnoea) | Daily | Nurse | Yes | Yes | 13 | NR |
| Johnson 2022 (Johnson 2022) | Non-Invasive TM | mHealth app | mHealth HF self-care app (knowledge, self-efficacy, symptom detection) | Symptom detection, self-management education, knowledge, self-efficacy; no physiologic sensor data transmitted | Daily app use (post-discharge) | Clinical escalation process available; no escalations required during study | Clinical escalation process built in (no triggers fired) | Yes — core component: HF knowledge, self-care skills, symptom recognition | 12 | 88% |
| AMULET (Krzesiński 2022) | Non-Invasive TM | Nurse telephone | Web telemedicine + nurse-led non-invasive assessments + remote cardiologist decisions | HR, SBP, DBP, TFC (thoracic fluid content), BM (body mass), TBW (total body water) — via ICG (Cardioscreen 2000) + bioimpedance (Tanita MC-418MA) | 7 scheduled outpatient visits over 12 months | Remote cardiologist (via Recommendation Support Module) | Color-coded RSM alarms: white (optimal), green/yellow/red (staged). Red TFC = urgent in-person consult within 2h | Symptom monitoring questionnaire; recommendation provided at each visit | 52 | 86% |
| ControlVit (Achury-Saldaña 2024) | Non-Invasive TM | mHealth app | ControlVit mobile app (daily weight/BP/HR + 8-item symptom questionnaire) | Weight, BP, HR + 8-item symptom questionnaire | Daily | Nurse (daily web platform review + real-time alerts) | Yes | Co-intervention | 26 | NR |
| Arnar 2025 (Arnar 2025) | Non-Invasive TM | mHealth app | Sidekick Health digital therapeutics smartphone app (RPM + self-care + education + lifestyle support) | Symptoms (breathlessness, fatigue, leg oedema, chest pain, dizziness — 5 daily MCQ); weight; vital signs (optional, own devices); step | Daily (weeks 1-12), tapering to weekly (weeks 24-52); daily optional in maintenance | Nurses — reviewed RPM data 3x/day during working hours (weekdays only); traffic-light | Three-tiered traffic light: green=stable, yellow=increased monitoring, red=immediate nurse contact | Yes — 24-week educational programme (nutrition, medication adherence, sleep, exercise, stress, | 48 | 93% |

| Study | RPM Category | Subcategory | Technology/Platform | Monitoring Parameters | Frequency | Who Responds | Alert Threshold | Patient Education | Duration (wk) | Adherence (%) |
|---|---|---|---|---|---|---|---|---|---|---|
| | | | | count (automatic via smartphone) | | algorithm (green/yellow/red) guided response | | mental health, HF knowledge); video + content cards; recap weeks 25-48 | | |
| M-Cardio (ERICA-HF) (Rustambekova 2025) | Non-Invasive TM | mHealth app | M-Cardio app (Android; custom-developed) | Dyspnea, body position, palpitations, edema, body weight, BP, HR (7 items) | Twice weekly (daily if necessary) | Supervising physician (via WhatsApp/phone when >2 values deviate) | Yes | Yes | 48 | NR |
| Vietnam HFrEF RCT (Tran 2025) | Non-Invasive TM | Nurse telephone + mHealth app | Telephone follow-ups + home telemonitoring (BP/HR/weight); STS+Non-invasive TM hybrid | weight, BP, HR, symptoms (dyspnoea, cough, oedema, palpitations) | Daily | Mixed | Yes | Co-intervention | 26 | NR |
| TeleMedBot (Zheleznykh 2025) | Non-Invasive TM | mHealth app | TeleMedBot (custom; Python REST API + PostgreSQL; patient-facing via smartphone messenger) | BP, HR, dyspnea, body position, palpitations, edema, body weight | Daily | Cardiologist | Yes | Yes | 26 | 76% |
| Riegel (2002) | STS | Nurse telephone | Pfizer "At Home With Heart Failure" — computer-supported telephonic case management software | symptoms, weight, fluid retention, SOB, medication adherence, diet adherence, functional status | Variable | Nurse | Yes | Yes | 26 | NR |
| Laramee (2003) | STS | Nurse telephone | 4-component CHF case management: discharge planning + comprehensive patient education + 12-week telephone follow-up + optimal medication promotion. Educational materials: "Heartworks" booklet, weight logs, sodium guide, home scales. | CHF symptoms, daily weight, edema, medications, self-care, fluid/sodium, lab values, appointments | Variable | Nurse | Yes | Yes | 12 | NR |
| DIAL (GESICA investigators 2005) | STS | Nurse telephone monitoring + education + counselling | Nurse telephone follow-up (structured) | Symptoms, medication adherence, weight, dietary compliance, functional class | Fortnightly first 4 calls; then individualized by nurse algorithm | Trained nurse (can adjust diuretic dose, recommend unscheduled visits) | Nurse discretion based on structured call data; algorithm-driven intervals | Yes: education booklet at randomisation; ongoing counselling each call | 96 | NR |
| HFHC (Heart Failure Home Care Trial) (Soran 2008) | STS | IVR (automated telephone) | Alere DayLink HF Monitoring System (electronic scale + IVR symptom questionnaire + nurse review) | Weight, SOB at night, extra pillow use, ankle edema, fatigue, cough (symptom IVR daily) | Daily (nurse review 7d/week, 365d/yr) | Nurse (alerts faxed to primary care physician) | Yes | Yes | 24 | 97% |
| VA TeleHF (Wakefield) (Wakefield 2008) | STS | Nurse telephone | Personal home telephone (primary arm); CyberCare EHC 200 Sentinel / TeleVyou 500SP videophone (secondary arm) | Symptoms (checklist), daily weight, blood pressure, ankle circumference | Daily contact x3 first week post-discharge, then weekly x11 weeks (14 contacts total over 3 months) | Registered nurse (study nurse) | Yes | Yes — discharge plan review, symptom checklist, behavioral compliance strategies | 12 | 80% |
| Wakefield 2009 (3-arm) (Wakefield 2009) | STS | Nurse telephone | Telephone calls (home); weekly nurse-initiated calls; max 14 contacts over 90 days. Videophone arm (secondary): EHC 200 | Symptom review checklist (weight gain, decompensation symptoms), | Weekly | Nurse | Yes | Yes | 13 | 94% |

| Study | RPM Category | Subcategory | Technology/Platform | Monitoring Parameters | Frequency | Who Responds | Alert Threshold | Patient Education | Duration (wk) | Adherence (%) |
|---|---|---|---|---|---|---|---|---|---|---|
| | | | Sentinel / TeleVyou 500SP videophone. | medication compliance, self-efficacy, patient education | | | | | | |
| Tele-HF (Chaudhry 2010) | STS | IVR | Automated telephone interactive voice-response system (daily symptoms + weight) | Daily symptoms (dyspnea, weight gain, fatigue, edema, general health) + weight via telephone keypad entry; PHQ-2 depression screen every 30 days | Daily calls (patients call toll-free number) | Site coordinators review daily on secure Internet portal (weekdays only, excluding holidays); cardiologists make clinical decisions on variances | Predetermined variance thresholds for each question; variances flagged for clinician attention; 29,163 total variances (median 21/patient, IQR 5–54) | Educational materials from Heart Failure Society of America; training on IVR system; scale and telephone provided if needed; reminder calls for non-adherence | 26 | 55% |
| SPAN-CHF II (Weintraub 2010) | STS | AHM (weight/BP/HR) + symptom questionnaire | Automated home monitoring (AHM: weight/BP/HR) added to telephonic DM | Body weight, blood pressure, heart rate, patient self-assessment | Daily (AHM transmission) | HF nurse manager (Mon-Fri daily review; 24/7 on-call) | Prespecified thresholds for weight, BP, pulse, symptoms; nurse calls patient if exceeded | Yes — enrollment visit education on HF self-monitoring, diet, medication; Health Buddy daily questionnaire | 13 | NR |
| CHAT (Krum 2013) | STS | IVR | Trained cardiac nurse telephone follow-up (TeleWatch software) | Symptoms, weight, fluid intake, medication adherence, stress, depression, exercise, smoking, alcohol (via TeleWatch IVR) | Monthly minimum (automated calls); unscheduled calls patient-initiated at any time | Trained cardiac nurse | Prespecified symptom/sign alerts via TeleWatch Patient Watch Screen → nurse follow-up | Yes — action plan for detecting deterioration; TeleWatch IVR includes education modules on HF self-management | 52 | 66% |
| BEAT-HF (Ong 2016) | STS + Non-invasive TM | Nurse telephone + Wearable | Health coaching telephone calls + telemonitoring (daily BP/HR/symptoms/weight) | Weight, blood pressure, heart rate, symptoms (3 daily questions; Bluetooth devices) | Daily TM; 9 coaching calls over 6 months (weekly x1mo, monthly x5mo) | Registered nurses at telephone call center (UCLA-based) | Yes - predetermined thresholds for weight/BP/HR; triggers nurse telephone callback | Yes - pre-discharge HF education (booklet; teach-back method; low health literacy design) | 26 | 55% |
| Negarandeh 2019 (Negarandeh 2019) | STS | — | Telephone-based monitoring | — | — | — | — | — | NR | NR |
| DAA/Feijó Brazil (Feijó 2021) | STS | Nurse telephone | Nurse telephone calls — diuretic algorithm protocol | Body weight (daily, morning pre-meal), Clinical Congestion Score (CCS 0-22), dyspnea, nocturnal paroxysmal dyspnea (PND), orthopnea, peripheral oedema, NYHA functional | Weekly (1-2x/week for 30 days) | Nurse (algorithm-guided diuretic titration) | Yes | Yes — Co-intervention | 4 | NR |

| Study | RPM Category | Subcategory | Technology/Platform | Monitoring Parameters | Frequency | Who Responds | Alert Threshold | Patient Education | Duration (wk) | Adherence (%) |
|---|---|---|---|---|---|---|---|---|---|---|
| | | | | class, furosemide dose adjustment | | | | | | |
| MESSAGE-HF (Rohde 2024) | STS | mHealth (SMS-based bidirectional telemonitoring) | Bidirectional SMS automated text messaging; 4 daily SMS; red flags → diuretic adjustment or HF team telephone call | Self-care behaviors, symptoms, adherence; red-flag → diuretic adjustment or callback | 4 SMS/day | Nurse/team (callback on red-flag alerts) | Weight gain ≥2kg/1st week or ≥3kg/1st month; nocturnal dyspnea 2 consecutive nights; non-adherence to medications 2 consecutive days | Yes — 4 SMS/day included educational messages on HF signs/symptoms, daily activities, lifestyle, medication, fluid intake | 4 | 75% |
| PRADOC (Roubille 2024) | STS | Nurse telephone | Administrative transition care coordination (PRADO-IC national program) | Post-discharge cardiology + GP follow-up scheduling | Weekly | Nurse | No | Co-intervention | 24 | NR |
| Ribeiro (2025) | STS | Nurse telephone | UFMG-developed custom software; standard SMS; BP cuff + scale provided. Decision trees with 5 pathways (emergency, same-day teleconsult, elective teleconsult ≤7d, diuretic adjustment, maintenance). | weight (daily), blood pressure (daily), heart rate (daily), decompensation symptoms, medication adherence, NYHA class | Weekly | Mixed | Yes | Co-intervention | 26 | 66% |

#### eTable 6. Studies Included in the Review but Excluded From Quantitative Synthesis

*eTable 6. Studies Included in the Review but Excluded From Quantitative Synthesis*

| Record ID | Study | Design | Comparator type | N total | Reason for exclusion from quantitative synthesis |
|---|---|---|---|---|---|
| CO-0940 | VERDICT (Copeland VA) (Copeland 2010) | Parallel RCT | Usual care | 458 | Insufficient extractable arm-level outcome data from abstract-only extraction. |
| CO-1025 | CARME (Domingo 2011) | Before/after with internal randomization | Usual care | 92 | Before/after design; both randomized internal groups received telemonitoring. |
| CO-0934 | Ohio Telecare (Madigan) (Madigan 2013) | Parallel RCT | Usual care | 99 | No extractable RR/HR for clinical event outcomes; KCCQ measured at home-health discharge rather than fixed follow-up. |
| CO-0465 | REMOTE-CIED (Versteeg 2019) | Parallel RCT | Usual care | 595 | Quality-of-life primary trial; no extractable clinical event effect estimates for pairwise pooling. |
| CO-0442 | PROACTIVE-HF (Guichard 2024) | Single-arm | Historical performance goal | 456 | Single-arm trial using a historical performance goal; no concurrent randomized usual-care control. |
| CO-0570 | MIGHTY-HEART (Masterson Creber 2025) | Parallel RCT | Active comparator | 2,003 | Active comparator and 30-day follow-up; handled only in post-hoc sensitivity including active comparators. |

### eTable 7. Publication Bias Tests

*eTable 7. Publication Bias Assessment*

| outcome | k | egger_pval | begg_pval | peters_pval | tf_k0 | tf_side | tf_rr_adjusted | tf_rr_adj_lo | tf_rr_adj_hi | tf_pval |
|---|---|---|---|---|---|---|---|---|---|---|
| ACM | 41 | 0.04 | 0.0135 | 0.2463 | 6 | R | 0.925 | 0.851 | 1.005 | 0.064 |
| HF Hospitalization | 39 | <0.001 | <0.001 | <0.001 | 12 | R | 0.830 | 0.737 | 0.934 | 0.0026 |
| All-Cause Hospitalization | 28 | 0.0016 | 0.0027 | 0.0467 | 5 | R | 0.978 | 0.897 | 1.065 | 0.5962 |

### eTable 8. Meta-Regression Results

*eTable 8. Meta-Regression Results*

| Outcome | Covariate | Term | Beta | SE | P | Residual tau2 | R2 (%) |
|---|---|---|---|---|---|---|---|
| ACM | Follow-up (months) | follow_up_months | 0.0039 | 0.0040 | 0.3209 | 0.0000 | 0.0 |
| ACM | Mean LVEF | lvef_mean_int | -0.0007 | 0.0069 | 0.92 | 0.0000 | 0.0 |
| ACM | Mean age | age_mean_int | -0.0046 | 0.0098 | 0.6346 | 0.0000 | 0.0 |
| ACM | Multivariate | age_mean_int | -0.0104 | 0.0139 | 0.4563 | 0.0000 | 0.0 |
| ACM | Multivariate | follow_up_months | 0.0064 | 0.0045 | 0.1593 | 0.0000 | 0.0 |
| ACM | Multivariate | lvef_mean_int | 0.0057 | 0.0120 | 0.6346 | 0.0000 | 0.0 |
| ACM | Multivariate | sample_size | 0.0001 | 0.0001 | 0.597 | 0.0000 | 0.0 |
| ACM | Multivariate | year | -0.0003 | 0.0086 | 0.9759 | 0.0000 | 0.0 |
| ACM | Publication year | year | -0.0010 | 0.0067 | 0.8813 | 0.0000 | 0.0 |
| ACM | Total sample size | sample_size | 0.0000 | 0.0001 | 0.9382 | 0.0000 | 0.0 |
| Allcause_hosp | Multivariate | age_mean_int | -0.0053 | 0.0067 | 0.4358 | 0.0022 | 59.9 |
| Allcause_hosp | Multivariate | follow_up_months | 0.0068 | 0.0040 | 0.0926 | 0.0022 | 59.9 |
| Allcause_hosp | Multivariate | lvef_mean_int | -0.0053 | 0.0063 | 0.4025 | 0.0022 | 59.9 |
| Allcause_hosp | Multivariate | sample_size | 0.0000 | 0.0001 | 0.9608 | 0.0022 | 59.9 |
| Allcause_hosp | Multivariate | year | -0.0009 | 0.0050 | 0.856 | 0.0022 | 59.9 |
| HF_hosp | Multivariate | age_mean_int | 0.0137 | 0.0097 | 0.1585 | 0.0273 | 36.9 |
| HF_hosp | Multivariate | follow_up_months | -0.0055 | 0.0057 | 0.3311 | 0.0273 | 36.9 |
| HF_hosp | Multivariate | lvef_mean_int | -0.0322 | 0.0114 | 0.0049 | 0.0273 | 36.9 |
| HF_hosp | Multivariate | sample_size | 0.0003 | 0.0001 | 0.0628 | 0.0273 | 36.9 |
| HF_hosp | Multivariate | year | 0.0125 | 0.0114 | 0.2714 | 0.0273 | 36.9 |

**eFigure 1. Risk of Bias 2 Traffic Light Plot**

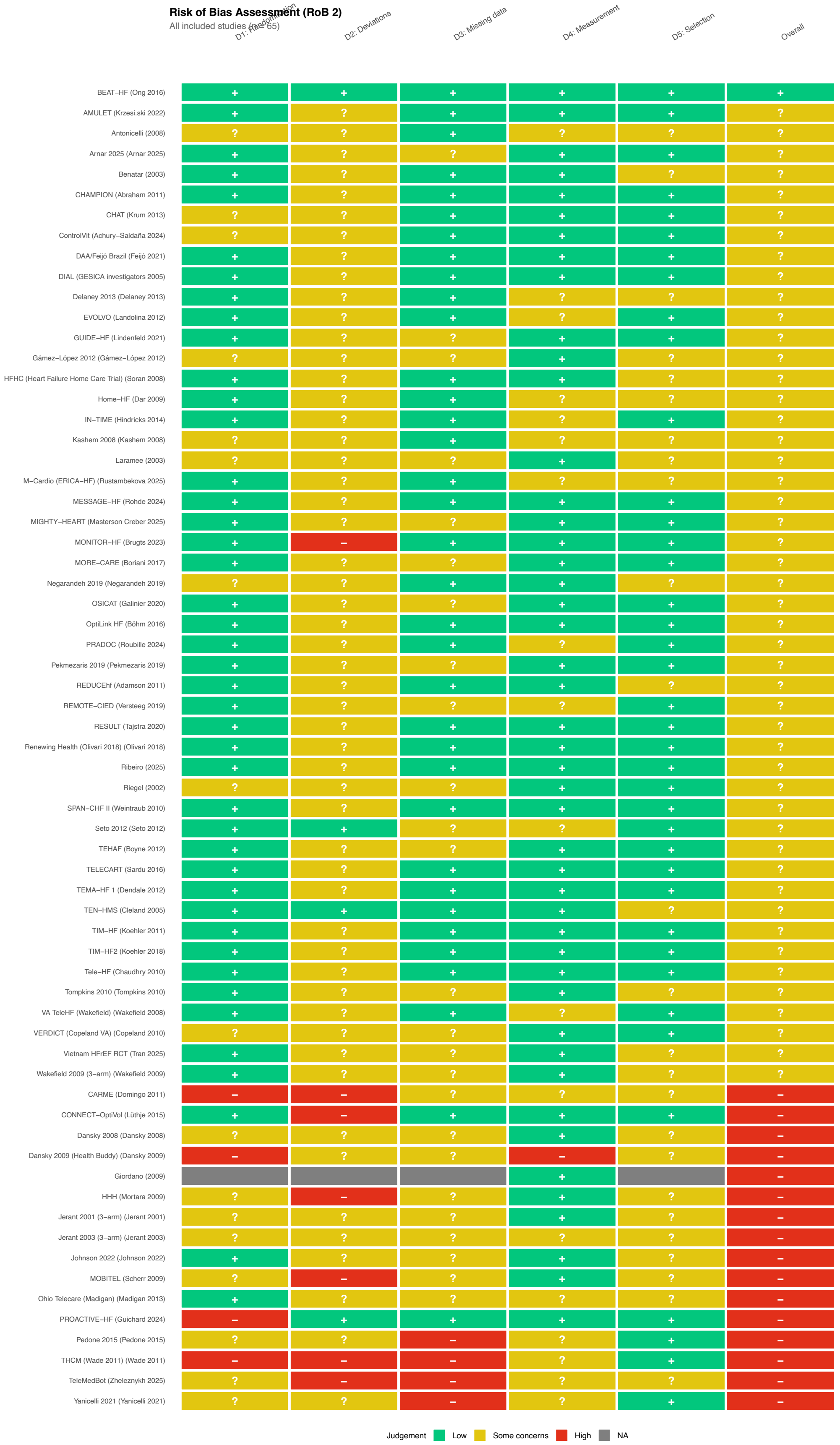

Risk of Bias Assessment (RoB 2)
All included studies (n = 65)
D1: Randomization
D2: Deviations
D3: Missing data
D4: Measurement
D5: Selection
Overall
BEAT–HF (Ong 2016)
AMULET (Krzesi.ski 2022)
Antonicelli (2008)
Arnar 2025 (Arnar 2025)
Benatar (2003)
CHAMPION (Abraham 2011)
CHAT (Krum 2013)
ControlVit (Achury–Saldaña 2024)
DAA/Feijó Brazil (Feijó 2021)
DIAL (GESICA investigators 2005)
Delaney 2013 (Delaney 2013)
EVOLVO (Landolina 2012)
GUIDE–HF (Lindenfeld 2021)
Gámez–López 2012 (Gámez–López 2012)
HFHC (Heart Failure Home Care Trial) (Soran 2008)
Home–HF (Dar 2009)
IN–TIME (Hindricks 2014)
Kashem 2008 (Kashem 2008)
Laramee (2003)
M–Cardio (ERICA–HF) (Rustambekova 2025)
MESSAGE–HF (Rohde 2024)
MIGHTY–HEART (Masterson Creber 2025)
MONITOR–HF (Brugts 2023)
MORE–CARE (Boriani 2017)
Negarandeh 2019 (Negarandeh 2019)
OSICAT (Galinier 2020)
OptiLink HF (Böhm 2016)
PRADOC (Roubille 2024)
Pekmezaris 2019 (Pekmezaris 2019)
REDUCEhf (Adamson 2011)
REMOTE–CIED (Versteeg 2019)
RESULT (Tajstra 2020)
Renewing Health (Olivari 2018) (Olivari 2018)
Ribeiro (2025)
Riegel (2002)
SPAN–CHF II (Weintraub 2010)
Seto 2012 (Seto 2012)
TEHAF (Boyne 2012)
TELECART (Sardu 2016)
TEMA–HF 1 (Dendale 2012)
TEN–HMS (Cleland 2005)
TIM–HF (Koehler 2011)
TIM–HF2 (Koehler 2018)
Tele–HF (Chaudhry 2010)
Tompkins 2010 (Tompkins 2010)
VA TeleHF (Wakefield) (Wakefield 2008)
VERDICT (Copeland VA) (Copeland 2010)
Vietnam HFrEF RCT (Tran 2025)
Wakefield 2009 (3–arm) (Wakefield 2009)
CARME (Domingo 2011)
CONNECT–OptiVol (Lüthje 2015)
Dansky 2008 (Dansky 2008)
Dansky 2009 (Health Buddy) (Dansky 2009)
Giordano (2009)
HHH (Mortara 2009)
Jerant 2001 (3–arm) (Jerant 2001)
Jerant 2003 (3–arm) (Jerant 2003)
Johnson 2022 (Johnson 2022)
MOBITEL (Scherr 2009)
Ohio Telecare (Madigan) (Madigan 2013)
PROACTIVE–HF (Guichard 2024)
Pedone 2015 (Pedone 2015)
THCM (Wade 2011) (Wade 2011)
TeleMedBot (Zheleznykh 2025)
Yanicelli 2021 (Yanicelli 2021)
Judgement
Low
Some concerns
High
NA

*eFigure 1. Risk of Bias 2 Traffic Light Plot*

#### eFigure 2. Risk of Bias 2 Summary Plot

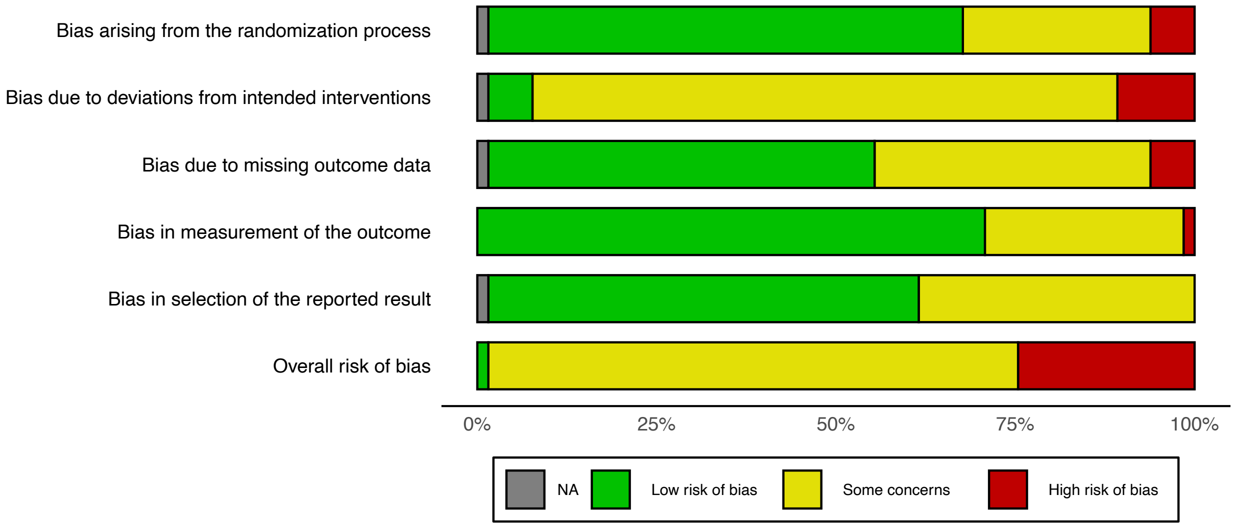


*eFigure 2. Risk of Bias 2 Summary Plot*

### eFigure 3. Forest Plot: All-Cause Mortality (HR Sensitivity)

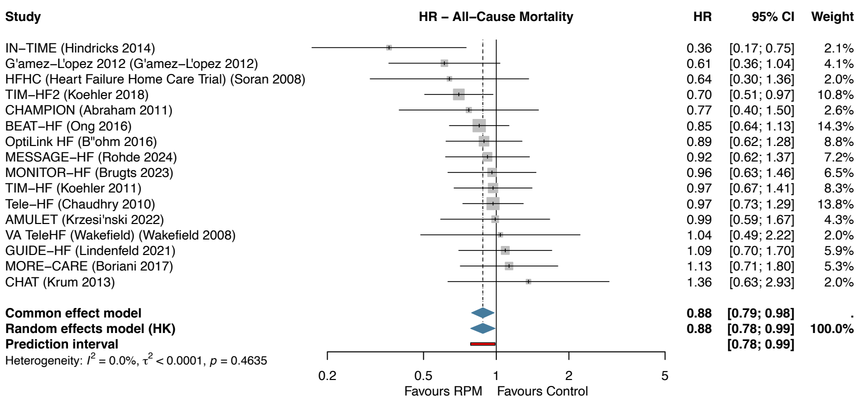


*eFigure 3. Forest plot of all-cause mortality using hazard ratios (HR sensitivity).*

### eFigure 4. Forest Plot: HF Hospitalization (HR Sensitivity)

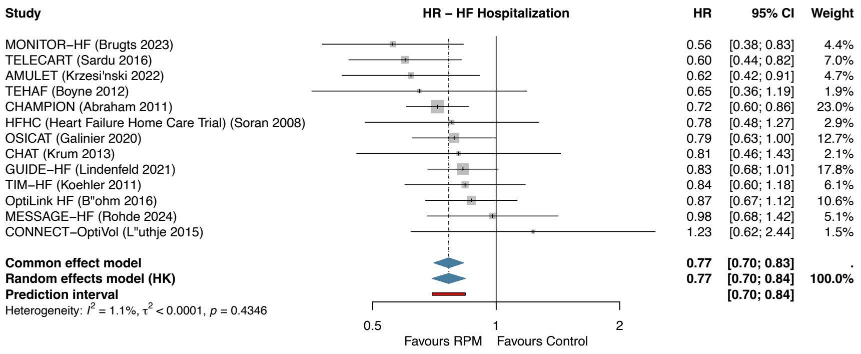


*eFigure 4. Forest plot of HF hospitalization using hazard ratios (HR sensitivity).*

### eFigure 5. Forest Plot: All-Cause Hospitalization (HR Sensitivity)

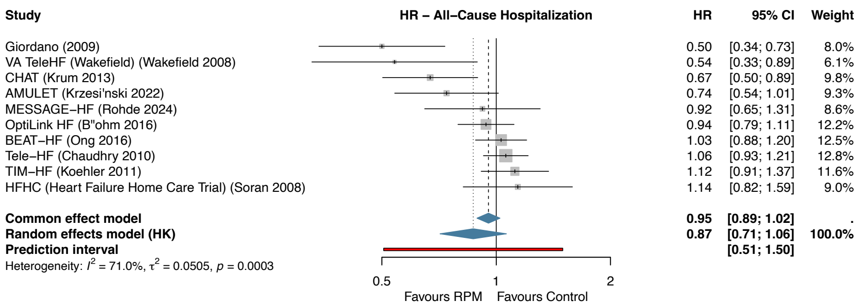


*eFigure 5. Forest plot of all-cause hospitalization using hazard ratios (HR sensitivity).*

### eFigure 6. Subgroup Forest: HF Phenotype x HF Hospitalization

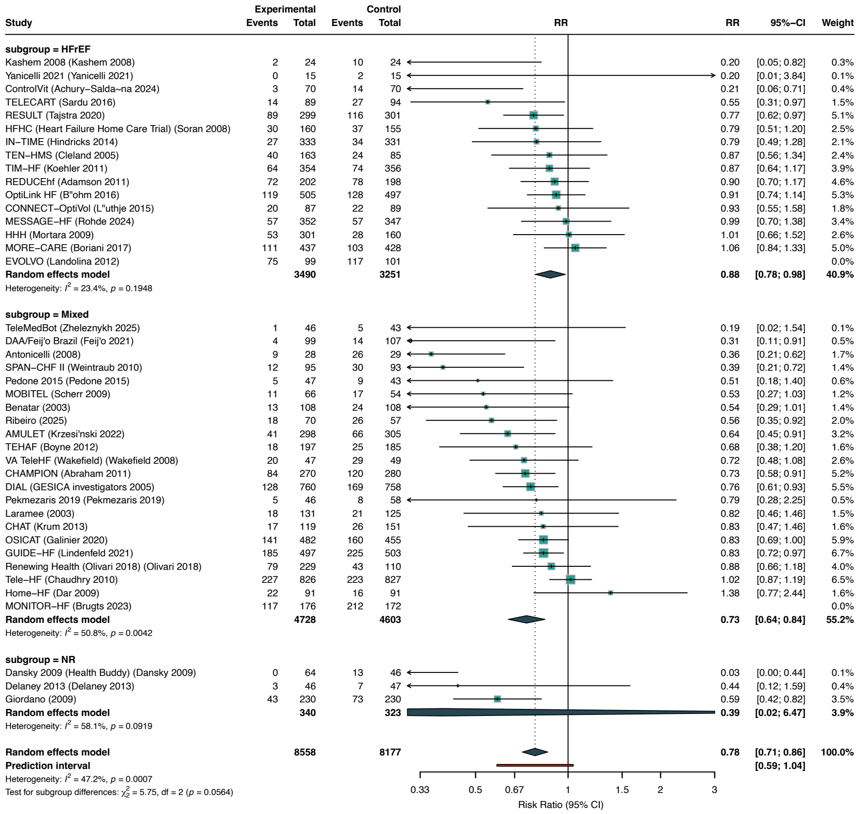


*eFigure 6. Subgroup analysis of HF hospitalization by HF phenotype.*

### eFigure 7. Subgroup Forest: Geographic Context x ACM

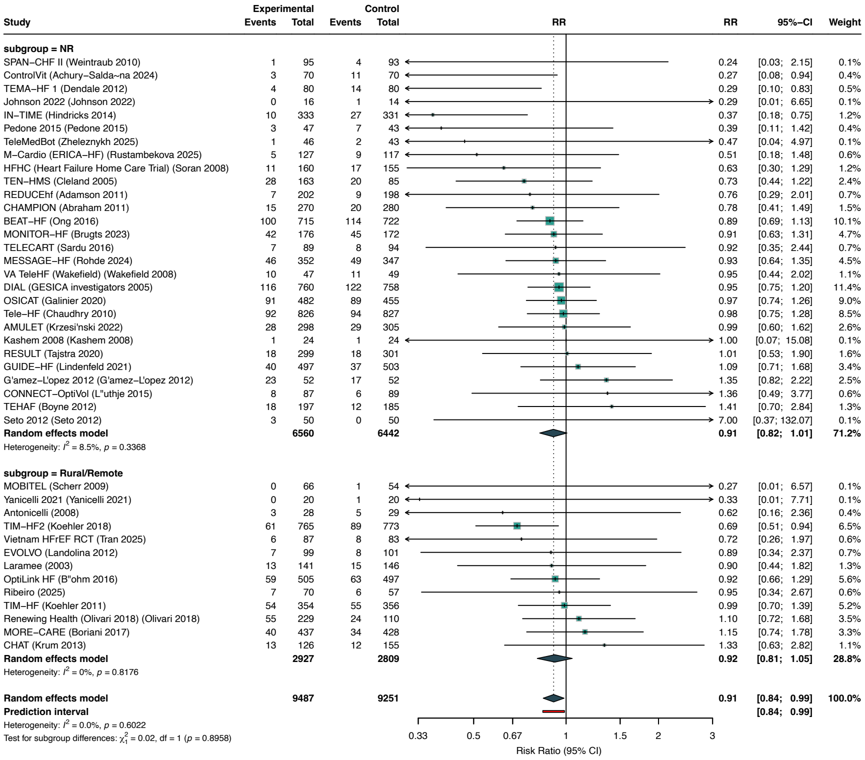


*eFigure 7. Subgroup analysis of all-cause mortality by geographic context.*

### eFigure 8. Subgroup Forest: Country Income x All-Cause Hospitalization

---

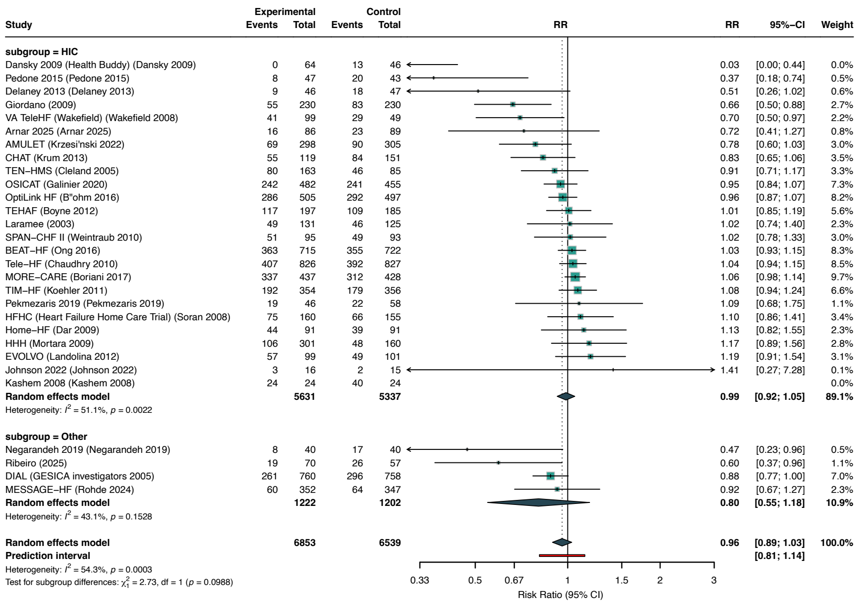


*eFigure 8. Subgroup analysis of all-cause hospitalization by country income.*

### eFigure 9. Leave-One-Out Sensitivity: ACM

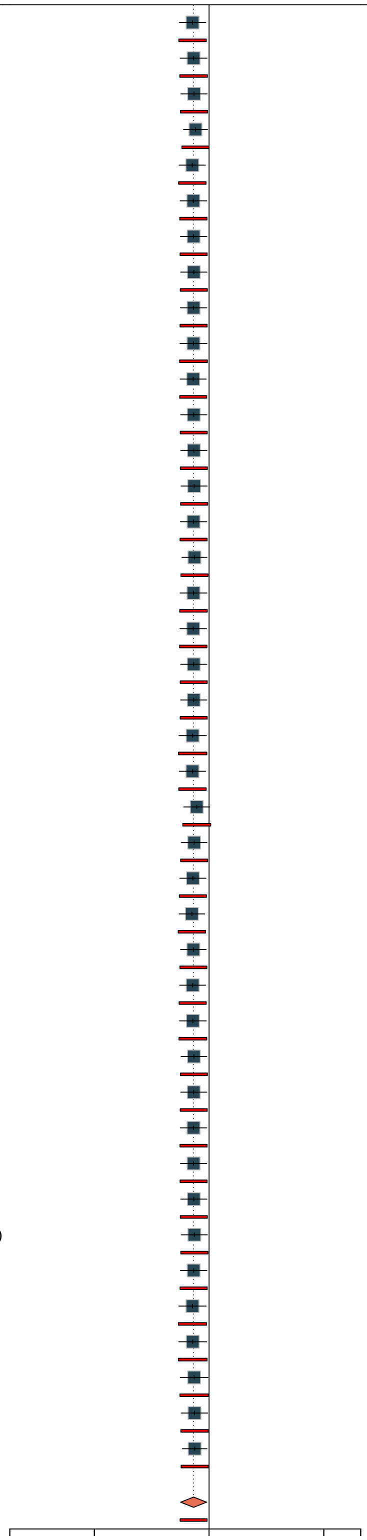

| [illegible] | [illegible] | [illegible] | [illegible] |
|---|---|---|---|
| Omitting GUIDE−HF (Lindenfeld 2021) | 0.905 | [0.835; 0.980]<br>[0.833; 0.983] | 0.0159 |
| Omitting MONITOR−HF (Brugts 2023) | 0.911 | [0.840; 0.988]<br>[0.838; 0.990] | 0.0255 |
| Omitting CHAMPION (Abraham 2011) | 0.913 | [0.843; 0.989]<br>[0.841; 0.991] | 0.0265 |
| Omitting IN−TIME (Hindricks 2014) | 0.921 | [0.857; 0.991]<br>[0.849; 1.000] | 0.0282 |
| Omitting MORE−CARE (Boriani 2017) | 0.904 | [0.835; 0.978]<br>[0.832; 0.982] | 0.0139 |
| Omitting RESULT (Tajstra 2020) | 0.909 | [0.840; 0.985]<br>[0.838; 0.987] | 0.0212 |
| Omitting EVOLVO (Landolina 2012) | 0.911 | [0.841; 0.987]<br>[0.839; 0.989] | 0.0230 |
| Omitting REDUCEhf (Adamson 2011) | 0.912 | [0.842; 0.987]<br>[0.840; 0.990] | 0.0243 |
| Omitting TELECART (Sardu 2016) | 0.911 | [0.841; 0.986]<br>[0.839; 0.988] | 0.0227 |
| Omitting OptiLink HF (Böhm 2016) | 0.910 | [0.839; 0.988]<br>[0.837; 0.990] | 0.0252 |
| Omitting CONNECT−OptiVol (Lüthje 2015) | 0.909 | [0.840; 0.983]<br>[0.837; 0.986] | 0.0188 |
| Omitting TeleMedBot (Zheleznykh 2025) | 0.912 | [0.842; 0.987]<br>[0.840; 0.989] | 0.0230 |
| Omitting Vietnam HFrEF RCT (Tran 2025) | 0.912 | [0.843; 0.988]<br>[0.841; 0.990] | 0.0245 |
| Omitting M−Cardio (ERICA−HF) (Rustambekova 2025) | 0.914 | [0.845; 0.988]<br>[0.842; 0.992] | 0.0253 |
| Omitting Ribeiro (2025) | 0.911 | [0.841; 0.986]<br>[0.839; 0.988] | 0.0225 |
| Omitting ControlVit (Achury−Saldaña 2024) | 0.915 | [0.849; 0.987]<br>[0.844; 0.993] | 0.0230 |
| Omitting MESSAGE−HF (Rohde 2024) | 0.910 | [0.839; 0.987]<br>[0.837; 0.989] | 0.0244 |
| Omitting AMULET (Krzesi.ski 2022) | 0.909 | [0.839; 0.985]<br>[0.837; 0.987] | 0.0211 |
| Omitting Johnson 2022 (Johnson 2022) | 0.912 | [0.842; 0.986]<br>[0.840; 0.989] | 0.0226 |
| Omitting Yanicelli 2021 (Yanicelli 2021) | 0.911 | [0.842; 0.986]<br>[0.840; 0.989] | 0.0227 |
| Omitting OSICAT (Galinier 2020) | 0.906 | [0.833; 0.984]<br>[0.831; 0.986] | 0.0206 |
| Omitting Renewing Health (Olivari 2018) (Olivari 2018) | 0.905 | [0.835; 0.980]<br>[0.833; 0.983] | 0.0154 |
| Omitting TIM−HF2 (Koehler 2018) | 0.928 | [0.858; 1.004]<br>[0.853; 1.010] | 0.0627 |
| Omitting Pedone 2015 (Pedone 2015) | 0.914 | [0.845; 0.988]<br>[0.842; 0.991] | 0.0242 |
| Omitting CHAT (Krum 2013) | 0.907 | [0.838; 0.981]<br>[0.836; 0.984] | 0.0165 |
| Omitting Gámez−López 2012 (Gámez−López 2012) | 0.901 | [0.834; 0.974]<br>[0.830; 0.979] | 0.0102 |
| Omitting Seto 2012 (Seto 2012) | 0.910 | [0.842; 0.983]<br>[0.838; 0.987] | 0.0177 |
| Omitting TEHAF (Boyne 2012) | 0.906 | [0.838; 0.980]<br>[0.834; 0.983] | 0.0146 |
| Omitting TIM−HF (Koehler 2011) | 0.907 | [0.836; 0.984]<br>[0.834; 0.986] | 0.0197 |
| Omitting SPAN−CHF II (Weintraub 2010) | 0.912 | [0.844; 0.986]<br>[0.841; 0.990] | 0.0224 |
| Omitting MOBITEL (Scherr 2009) | 0.912 | [0.842; 0.986]<br>[0.840; 0.989] | 0.0225 |
| Omitting VA TeleHF (Wakefield) (Wakefield 2008) | 0.910 | [0.841; 0.986]<br>[0.839; 0.988] | 0.0225 |
| Omitting Kashem 2008 (Kashem 2008) | 0.911 | [0.841; 0.986]<br>[0.839; 0.988] | 0.0224 |
| Omitting Antonicelli (2008) | 0.912 | [0.843; 0.987]<br>[0.841; 0.990] | 0.0240 |
| Omitting HFHC (Heart Failure Home Care Trial) (Soran 2008) | 0.915 | [0.846; 0.990]<br>[0.843; 0.993] | 0.0280 |
| Omitting Laramee (2003) | 0.911 | [0.841; 0.987]<br>[0.839; 0.989] | 0.0235 |
| Omitting Tele−HF (Chaudhry 2010) | 0.905 | [0.833; 0.983]<br>[0.831; 0.985] | 0.0189 |
| Omitting DIAL (GESICA investigators 2005) | 0.906 | [0.833; 0.986]<br>[0.831; 0.988] | 0.0230 |
| Omitting BEAT−HF (Ong 2016) | 0.914 | [0.840; 0.994]<br>[0.838; 0.996] | 0.0354 |
| Omitting TEN−HMS (Cleland 2005) | 0.916 | [0.846; 0.992]<br>[0.843; 0.995] | 0.0312 |
| Omitting TEMA−HF 1 (Dendale 2012) | 0.917 | [0.851; 0.988]<br>[0.845; 0.995] | 0.0234 |
| **Random effects model** | **0.911** | **[0.842; 0.985]** | **0.0208** |
| **Prediction interval** | | **[0.840; 0.988]** | |

*eFigure 9. Leave-one-out sensitivity analysis for all-cause mortality.*

### eFigure 10. Leave-One-Out Sensitivity: HF Hospitalization

| Omitted study | HR | 95% CI | p–value |
|---|---|---|---|
| Omitting GUIDE–HF (Lindenfeld 2021) | 0.772 | [0.697; 0.856]<br>[0.561; 1.062] | < 0.0001 |
| Omitting CHAMPION (Abraham 2011) | 0.781 | [0.707; 0.864]<br>[0.577; 1.058] | < 0.0001 |
| Omitting IN–TIME (Hindricks 2014) | 0.779 | [0.706; 0.860]<br>[0.578; 1.049] | < 0.0001 |
| Omitting MORE–CARE (Boriani 2017) | 0.773 | [0.703; 0.849]<br>[0.595; 1.003] | < 0.0001 |
| Omitting RESULT (Tajstra 2020) | 0.777 | [0.702; 0.860]<br>[0.569; 1.061] | < 0.0001 |
| Omitting REDUCEhf (Adamson 2011) | 0.772 | [0.699; 0.853]<br>[0.569; 1.048] | < 0.0001 |
| Omitting TELECART (Sardu 2016) | 0.787 | [0.715; 0.865]<br>[0.596; 1.038] | < 0.0001 |
| Omitting OptiLink HF (Böhm 2016) | 0.771 | [0.698; 0.852]<br>[0.570; 1.045] | < 0.0001 |
| Omitting CONNECT–OptiVol (Lüthje 2015) | 0.777 | [0.704; 0.857]<br>[0.577; 1.046] | < 0.0001 |
| Omitting TeleMedBot (Zheleznykh 2025) | 0.783 | [0.712; 0.861]<br>[0.590; 1.039] | < 0.0001 |
| Omitting Ribeiro (2025) | 0.788 | [0.717; 0.867]<br>[0.599; 1.036] | < 0.0001 |
| Omitting ControlVit (Achury–Saldaña 2024) | 0.787 | [0.718; 0.863]<br>[0.599; 1.034] | < 0.0001 |
| Omitting MESSAGE–HF (Rohde 2024) | 0.773 | [0.701; 0.853]<br>[0.575; 1.039] | < 0.0001 |
| Omitting AMULET (Krzesi.ski 2022) | 0.787 | [0.715; 0.867]<br>[0.596; 1.041] | < 0.0001 |
| Omitting Yanicelli 2021 (Yanicelli 2021) | 0.782 | [0.710; 0.860]<br>[0.588; 1.040] | < 0.0001 |
| Omitting DAA/Feijó Brazil (Feijó 2021) | 0.786 | [0.716; 0.863]<br>[0.596; 1.036] | < 0.0001 |
| Omitting OSICAT (Galinier 2020) | 0.773 | [0.698; 0.856]<br>[0.563; 1.061] | < 0.0001 |
| Omitting Pekmezaris 2019 (Pekmezaris 2019) | 0.780 | [0.708; 0.860]<br>[0.584; 1.042] | < 0.0001 |
| Omitting Renewing Health (Olivari 2018) (Olivari 2018) | 0.774 | [0.700; 0.855]<br>[0.569; 1.051] | < 0.0001 |
| Omitting Pedone 2015 (Pedone 2015) | 0.783 | [0.711; 0.862]<br>[0.590; 1.040] | < 0.0001 |
| Omitting CHAT (Krum 2013) | 0.779 | [0.706; 0.859]<br>[0.579; 1.047] | < 0.0001 |
| Omitting Delaney 2013 (Delaney 2013) | 0.783 | [0.711; 0.862]<br>[0.589; 1.040] | < 0.0001 |
| Omitting TEHAF (Boyne 2012) | 0.782 | [0.710; 0.862]<br>[0.586; 1.045] | < 0.0001 |
| Omitting TIM–HF (Koehler 2011) | 0.774 | [0.701; 0.856]<br>[0.570; 1.052] | < 0.0001 |
| Omitting SPAN–CHF II (Weintraub 2010) | 0.794 | [0.725; 0.869]<br>[0.616; 1.022] | < 0.0001 |
| Omitting Dansky 2009 (Health Buddy) (Dansky 2009) | 0.784 | [0.716; 0.858]<br>[0.592; 1.038] | < 0.0001 |
| Omitting HHH (Mortara 2009) | 0.774 | [0.702; 0.854]<br>[0.575; 1.041] | < 0.0001 |
| Omitting Home–HF (Dar 2009) | 0.774 | [0.705; 0.851]<br>[0.582; 1.029] | < 0.0001 |
| Omitting MOBITEL (Scherr 2009) | 0.786 | [0.714; 0.864]<br>[0.594; 1.039] | < 0.0001 |
| Omitting VA TeleHF (Wakefield) (Wakefield 2008) | 0.781 | [0.708; 0.862]<br>[0.581; 1.050] | < 0.0001 |
| Omitting Kashem 2008 (Kashem 2008) | 0.786 | [0.716; 0.862]<br>[0.595; 1.037] | < 0.0001 |
| Omitting Antonicelli (2008) | 0.799 | [0.733; 0.871]<br>[0.634; 1.008] | < 0.0001 |
| Omitting HFHC (Heart Failure Home Care Trial) (Soran 2008) | 0.779 | [0.705; 0.860]<br>[0.577; 1.051] | < 0.0001 |
| Omitting Laramee (2003) | 0.779 | [0.706; 0.859]<br>[0.580; 1.047] | < 0.0001 |
| Omitting Tele–HF (Chaudhry 2010) | 0.775 | [0.707; 0.849]<br>[0.619; 0.971] | < 0.0001 |
| Omitting DIAL (GESICA investigators 2005) | 0.778 | [0.703; 0.861]<br>[0.571; 1.062] | < 0.0001 |
| Omitting TEN–HMS (Cleland 2005) | 0.776 | [0.703; 0.857]<br>[0.575; 1.049] | < 0.0001 |
| Omitting Benatar (2003) | 0.786 | [0.715; 0.865]<br>[0.595; 1.039] | < 0.0001 |
| Omitting Giordano (2009) | 0.794 | [0.723; 0.872]<br>[0.615; 1.026] | < 0.0001 |
| **Random effects model** | **0.781** | **[0.710; 0.859]** | **< 0.0001** |
| **Prediction interval** | | **[0.586; 1.040]** | |

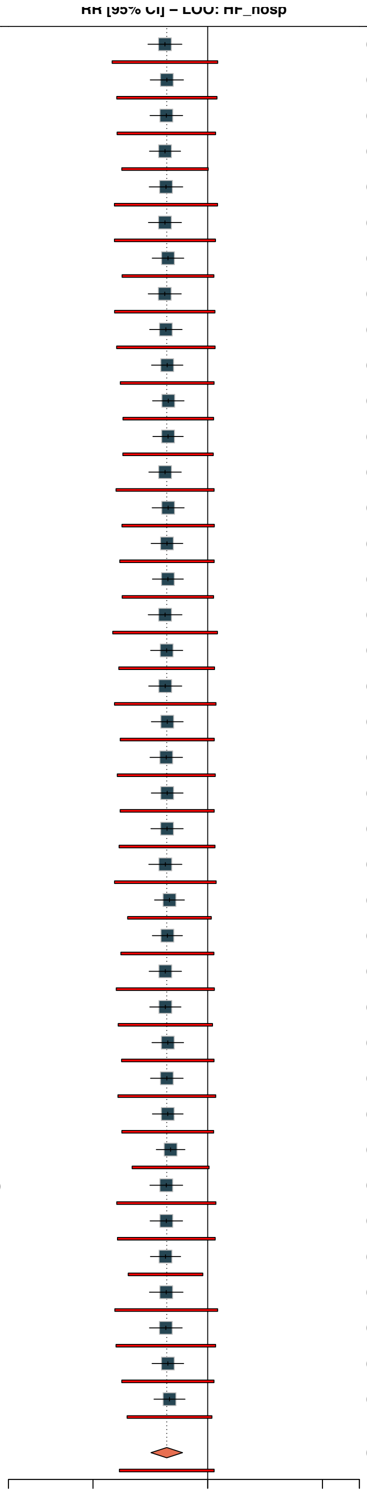


*eFigure 10. Leave-one-out sensitivity analysis for HF hospitalization.*

### eFigure 11. Leave-One-Out Sensitivity: All-Cause Hospitalization

| Omitted study | RR | 95% CI | p−value |
|---|---|---|---|
| Omitting MORE−CARE (Boriani 2017) | 0.948 | [0.878; 1.024]<br>[0.795; 1.131] | 0.1694 |
| Omitting EVOLVO (Landolina 2012) | 0.952 | [0.884; 1.025]<br>[0.799; 1.133] | 0.1816 |
| Omitting OptiLink HF (Böhm 2016) | 0.952 | [0.877; 1.033]<br>[0.775; 1.169] | 0.2255 |
| Omitting Ribeiro (2025) | 0.968 | [0.904; 1.037]<br>[0.833; 1.125] | 0.3360 |
| Omitting Arnar 2025 (Arnar 2025) | 0.962 | [0.894; 1.035]<br>[0.815; 1.136] | 0.2864 |
| Omitting MESSAGE−HF (Rohde 2024) | 0.958 | [0.888; 1.034]<br>[0.801; 1.146] | 0.2599 |
| Omitting AMULET (Krzesi.ski 2022) | 0.969 | [0.903; 1.040]<br>[0.833; 1.126] | 0.3629 |
| Omitting Johnson 2022 (Johnson 2022) | 0.958 | [0.890; 1.032]<br>[0.807; 1.138] | 0.2486 |
| Omitting OSICAT (Galinier 2020) | 0.954 | [0.880; 1.034]<br>[0.780; 1.166] | 0.2423 |
| Omitting Negarandeh 2019 (Negarandeh 2019) | 0.965 | [0.900; 1.034]<br>[0.823; 1.131] | 0.2992 |
| Omitting Pekmezaris 2019 (Pekmezaris 2019) | 0.956 | [0.887; 1.031]<br>[0.800; 1.142] | 0.2320 |
| Omitting Pedone 2015 (Pedone 2015) | 0.968 | [0.906; 1.033]<br>[0.832; 1.125] | 0.3122 |
| Omitting CHAT (Krum 2013) | 0.966 | [0.898; 1.039]<br>[0.824; 1.133] | 0.3402 |
| Omitting Delaney 2013 (Delaney 2013) | 0.964 | [0.899; 1.035]<br>[0.822; 1.131] | 0.2988 |
| Omitting TEHAF (Boyne 2012) | 0.951 | [0.878; 1.030]<br>[0.781; 1.158] | 0.2077 |
| Omitting TIM−HF (Koehler 2011) | 0.949 | [0.879; 1.025]<br>[0.793; 1.137] | 0.1755 |
| Omitting SPAN−CHF II (Weintraub 2010) | 0.954 | [0.883; 1.031]<br>[0.792; 1.150] | 0.2226 |
| Omitting Dansky 2009 (Health Buddy) (Dansky 2009) | 0.961 | [0.897; 1.029]<br>[0.813; 1.136] | 0.2404 |
| Omitting HHH (Mortara 2009) | 0.952 | [0.883; 1.026]<br>[0.797; 1.137] | 0.1883 |
| Omitting Home−HF (Dar 2009) | 0.953 | [0.884; 1.028]<br>[0.795; 1.143] | 0.2045 |
| Omitting VA TeleHF (Wakefield) (Wakefield 2008) | 0.971 | [0.907; 1.039]<br>[0.842; 1.119] | 0.3778 |
| Omitting HFHC (Heart Failure Home Care Trial) (Soran 2008) | 0.952 | [0.882; 1.027]<br>[0.792; 1.143] | 0.1950 |
| Omitting Laramee (2003) | 0.955 | [0.885; 1.031]<br>[0.795; 1.147] | 0.2303 |
| Omitting Tele−HF (Chaudhry 2010) | 0.948 | [0.875; 1.026]<br>[0.782; 1.149] | 0.1791 |
| Omitting DIAL (GESICA investigators 2005) | 0.968 | [0.898; 1.042]<br>[0.824; 1.136] | 0.3720 |
| Omitting BEAT−HF (Ong 2016) | 0.948 | [0.875; 1.027]<br>[0.779; 1.153] | 0.1827 |
| Omitting TEN−HMS (Cleland 2005) | 0.959 | [0.888; 1.035]<br>[0.801; 1.148] | 0.2724 |
| Omitting Giordano (2009) | 0.978 | [0.919; 1.041]<br>[0.871; 1.099] | 0.4763 |
| **Random effects model** | **0.959** | **[0.892; 1.031]** | **0.2462** |
| **Prediction interval** | | **[0.809; 1.137]** | |

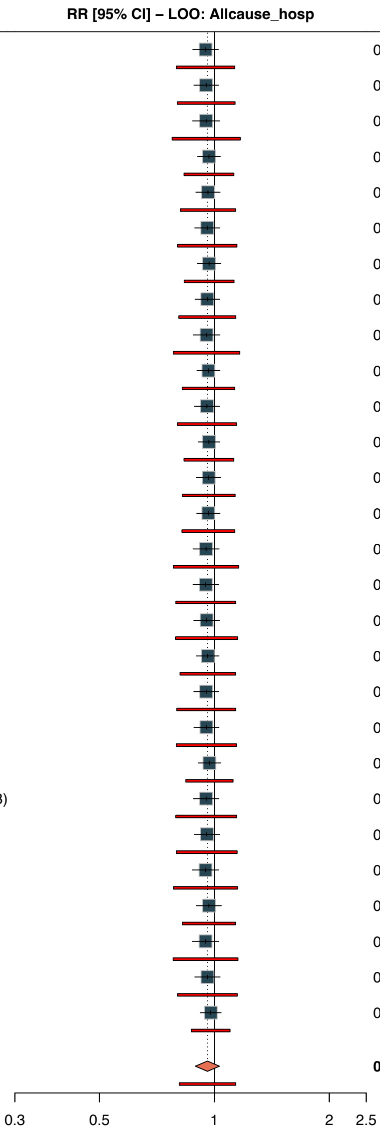


*eFigure 11. Leave-one-out sensitivity analysis for all-cause hospitalization.*

#### eFigure 12. Contour-Enhanced Funnel Plot: ACM

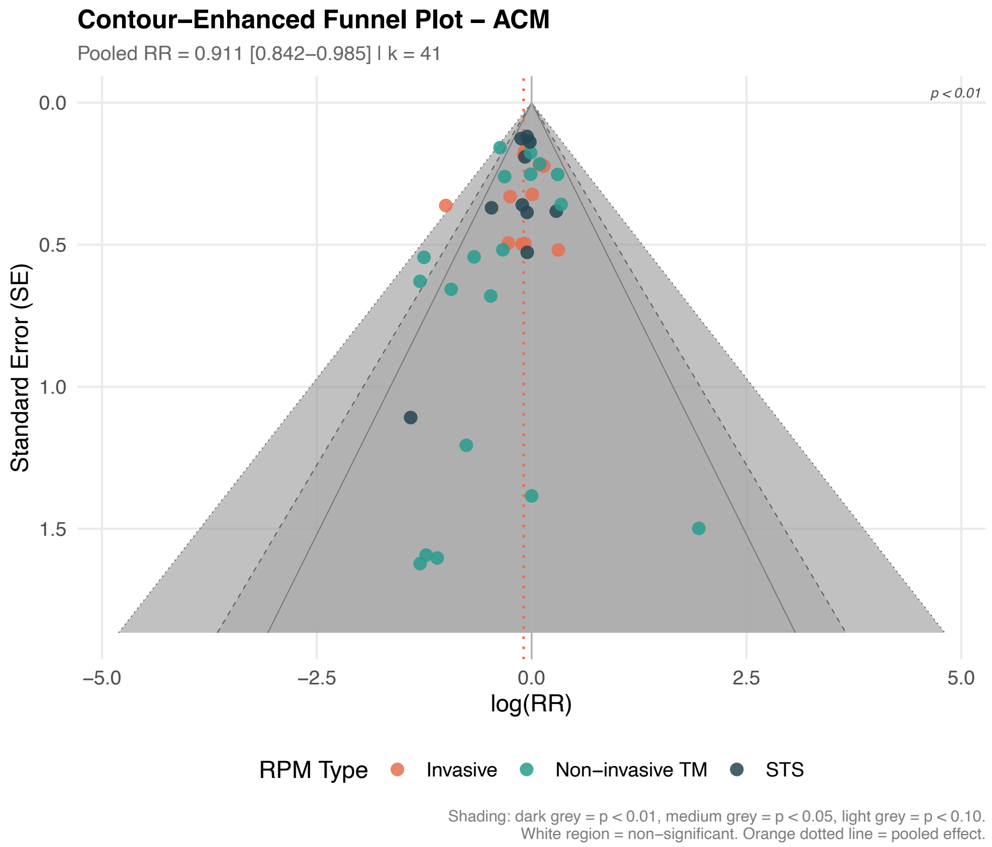


*eFigure 12. Contour-enhanced funnel plot for all-cause mortality.*

### eFigure 13. Contour-Enhanced Funnel Plot: HF Hospitalization

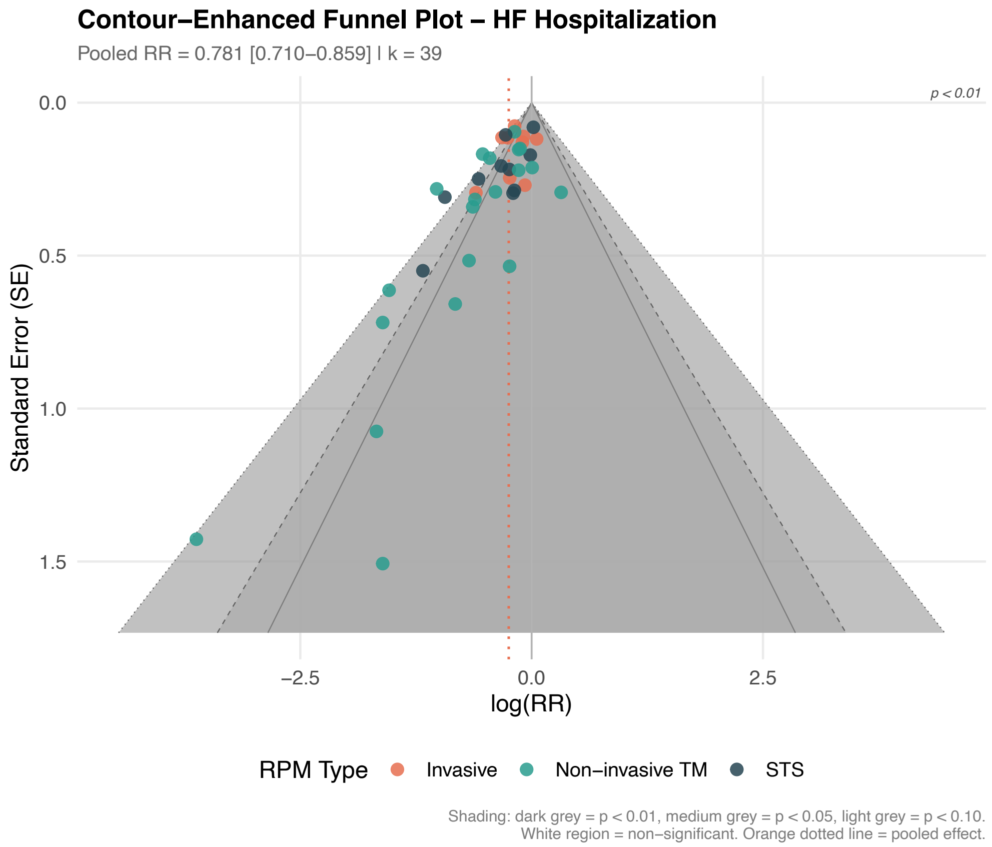


*eFigure 13. Contour-enhanced funnel plot for HF hospitalization.*

### eFigure 14. Contour-Enhanced Funnel Plot: All-Cause Hospitalization

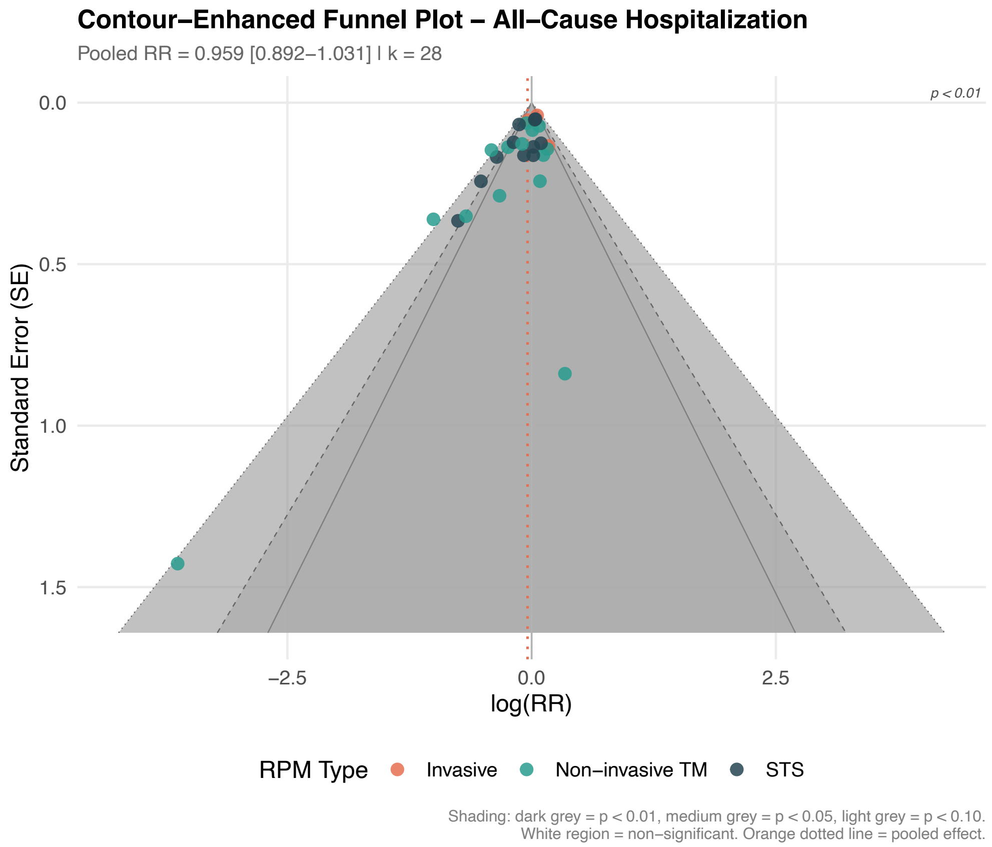


*eFigure 14. Contour-enhanced funnel plot for all-cause hospitalization.*

### eFigure 15. Trial Sequential Analysis: HF Hospitalization (RRR=15%)

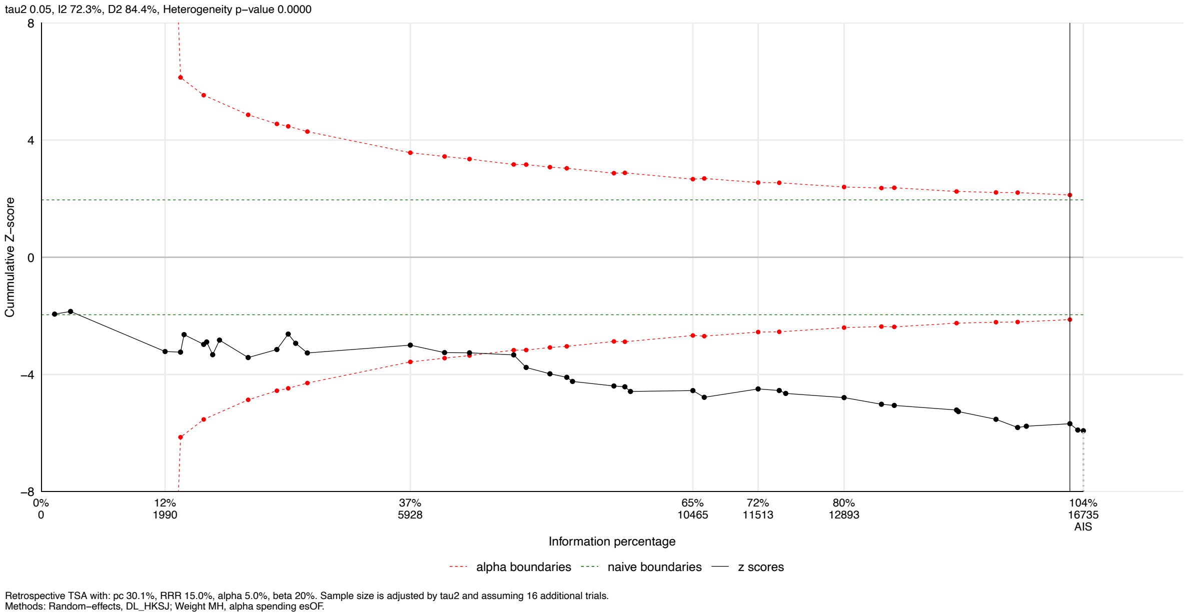


*eFigure 15. Trial sequential analysis for HF hospitalization (RRR=15%).*

### eFigure 16. Trial Sequential Analysis: HF Hospitalization (RRR=20%)

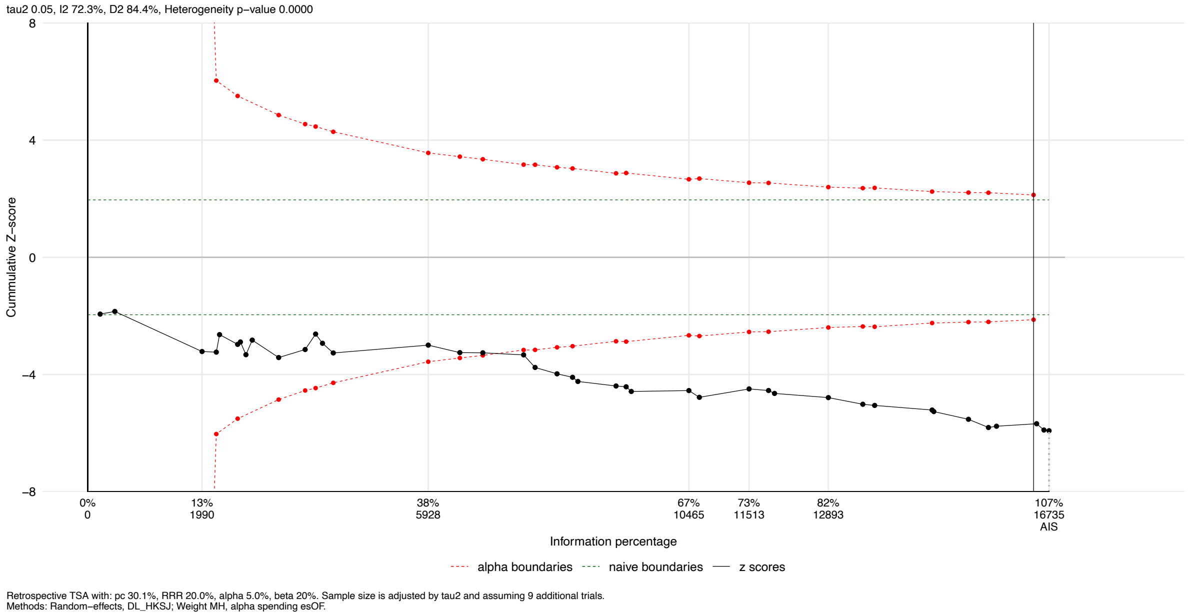


*eFigure 16. Trial sequential analysis for HF hospitalization (RRR=20%).*

### eFigure 17. Summary of Binary Outcomes

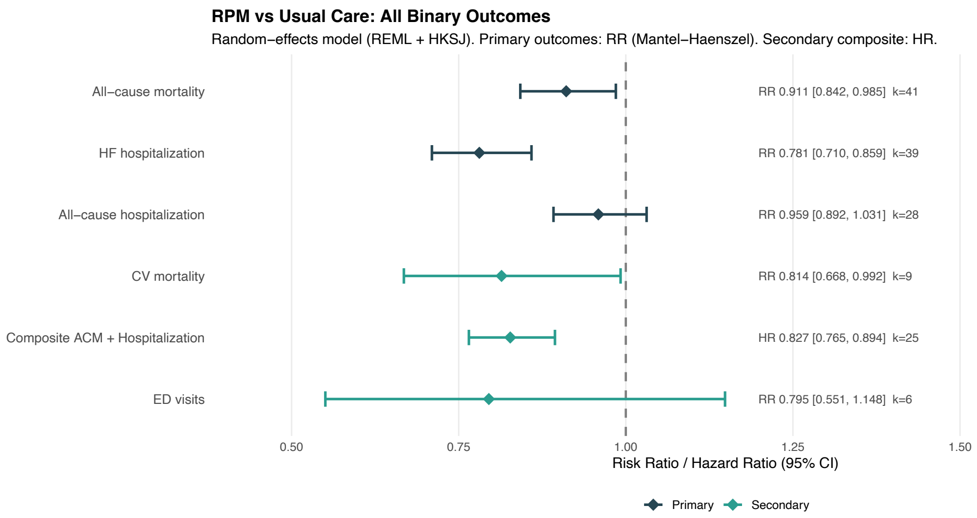


*eFigure 17. Summary forest plot of all binary outcomes.*

### eFigure 18. Summary of Continuous Outcomes (MLHFQ, KCCQ)

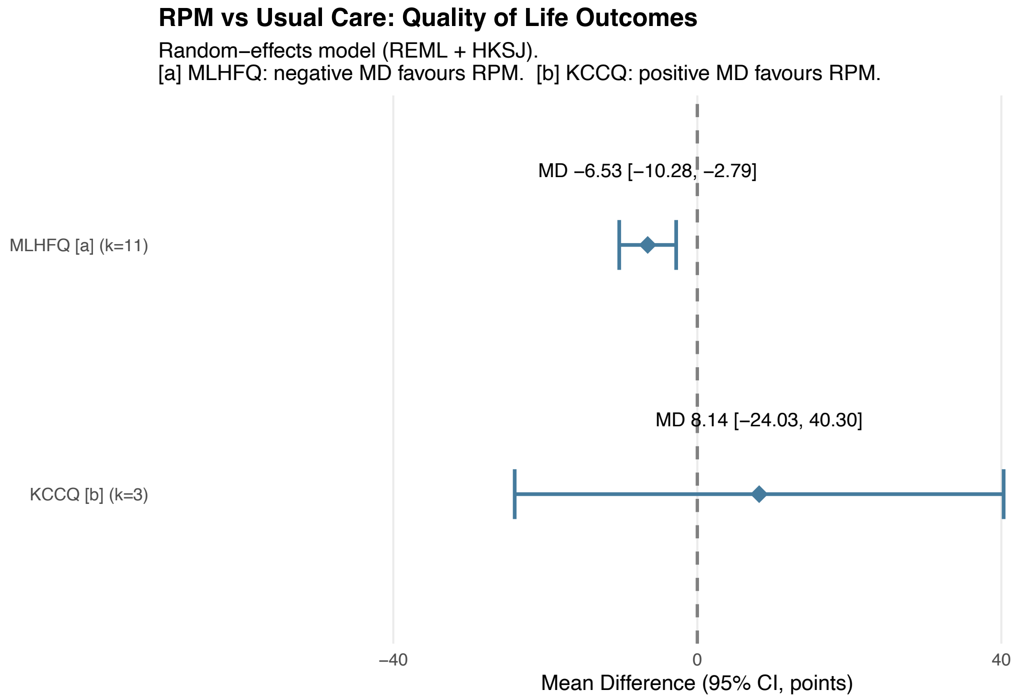


*eFigure 18. Summary forest plot of continuous outcomes (MLHFQ, KCCQ).*

#### eText 1. Database Search Sensitivity Assessment: Embase and CINAHL Omission

##### Rationale

Embase and CINAHL were not searched because institutional access was unavailable. The Cochrane Handbook for Systematic Reviews of Interventions (version 6.4) recommends searching Embase alongside MEDLINE for comprehensive systematic reviews [1]. This supplementary text documents the empirical assessment of the potential impact of this deviation on our conclusions.

##### CENTRAL's Partial Embase Coverage

Cochrane CENTRAL aggregates records from multiple sources, including a substantial proportion of Embase-indexed trials. Of the 2,138 records retrieved from CENTRAL in our search, 917 (42.9%) carried an Embase identifier, 25 (1.2%) carried a CINAHL identifier, and 1,201 (56.2%) carried a PubMed identifier. This indicates that our CENTRAL search captured a meaningful proportion of Embase-indexed content relevant to remote patient monitoring in heart failure.

##### PubMed Coverage of Included Studies

Of our 65 included studies, 63 (96.9%) had PubMed/MEDLINE identifiers (PMIDs). Only two studies lacked PMIDs: Zheleznykh 2025 (a Russian-language trial identified via ClinicalTrials.gov) and Delaney 2013 (an Australian study identified via CENTRAL). Both were captured through our existing four-database strategy despite the absence of Embase. This high

PubMed indexing rate suggests that the incremental yield of Embase for this topic is likely modest.

#### Methodological Evidence

Sampson et al. (2003) found that CENTRAL's coverage of Embase-indexed RCTs has grown steadily, and that for therapeutic interventions the incremental yield of Embase beyond PubMed and CENTRAL is modest [2]. Slobogean et al. (2009) demonstrated that PubMed combined with CENTRAL identified approximately 97% of primary studies included in meta-analyses [3]. Suarez-Almazor et al. (2000) reported that MEDLINE alone may miss some relevant trials, but noted that Embase's unique contribution is primarily observational studies and European conference abstracts rather than RCTs [4].

#### Cross-Reference Against Contemporary Meta-Analyses

Three meta-analyses published in 2025 searched additional databases beyond our four and provide a benchmark for assessing whether the Embase omission altered our conclusions:

| Review | Databases searched | Studies | Patients | ACM estimate | Direction consistent? |
|---|---|---|---|---|---|
| This review | PubMed, CENTRAL, CTgov, WHO ICTRP | 65 RCTs (59 poolable) | ~23,000 | RR 0.911 [0.842–0.985] | — |

| Review | Databases searched | Studies | Patients | ACM estimate | Direction consistent? |
| --- | --- | --- | --- | --- | --- |
| De Lathauwer et al. 2025 [5] | Including Embase | 41 RCTs | 16,312 | OR 0.81 [0.69–0.95] | Yes |
| Dobre et al. 2025 [6] | PubMed + Cochrane + additional | 105 studies | 45,072 | Mortality benefit reported | Yes |
| Ezimoha et al. 2025 [7] | Multiple databases | 15 RCTs | NR | Mortality benefit reported | Yes |

The substantially larger yield in Dobre et al. (105 studies vs our 65) likely reflects inclusion of non-randomized designs and post-hoc analyses; our review was restricted to RCTs with extractable primary outcome data. De Lathauwer et al., who searched Embase, identified 41 RCTs — fewer than our 59 poolable trials — suggesting that their more restrictive inclusion criteria, rather than database access, was the primary determinant of yield.

#### Conclusion

The concordance of effect estimates across reviews with and without Embase access, combined with the high PubMed indexing rate of included studies (96.9%) and CENTRAL's partial Embase coverage (42.9% of records), suggests that the Embase omission is unlikely to have altered the direction or clinical significance of our conclusions. Nevertheless, the possibility that some Embase-only European or Asian RCTs were missed cannot be excluded. This limitation is transparently documented (protocol amendment #13) and is reflected in the GRADE publication bias domain assessment for all outcomes.

#### eText 2. Semi-Automated Screening Algorithm

##### Overview

Title and abstract screening was performed using a semi-automated approach implemented in Python (`scripts/python/03_screening_ta.py`). The algorithm applies exclusion rules in four hierarchical tiers:

- **Tier 1 (High-confidence exclusion):** Animal studies, records lacking any remote patient monitoring intervention term, records lacking any heart failure term.
- **Tier 2 (Study type filter):** Reviews, observational studies, protocols, case reports, qualitative studies, conference abstracts.
- **Tier 3 (PICO filter):** Pediatric populations, LVAD/transplant, studies with follow-up < 4 weeks, studies reporting no clinical outcomes.
- **Tier 4 (Flags for human review):** Mixed interventions, post-surgical populations, cluster RCTs, conference abstracts with ambiguous content.

##### Intervention Terms

The RPM intervention filter matched a comprehensive set of terms including: telemonitor, remote monitor, telehealth, telemedicine, telemanagement, mHealth, structured telephone support, home monitor, CardioMEMS, pulmonary artery pressure monitor, wearable, smartphone, ICD-based monitor, interactive voice response, videoconference, virtual visit, and eHealth.

##### Heart Failure Terms

The HF filter matched: heart failure, cardiac failure, HFrEF, HFpEF, HFmrEF, CHF, preserved/reduced ejection fraction, systolic/diastolic dysfunction, ventricular dysfunction, cardiomyopathy, NYHA, left ventricular, and ejection fraction.

#### Validation

The algorithm was validated against 8 landmark trials known to be eligible (GUIDE-HF, MONITOR-HF, CHAMPION, IN-TIME, TIM-HF2, BEAT-HF, TEN-HMS, Laramee 2003), all of which survived the automated filters. A post-hoc sensitivity audit identified 3 regex gaps (missing: telemanagement, CHF as standalone term, and an overly broad exclusion on "guideline"), resulting in 2 false negatives (CO-0203 Benatar 2003, CO-1045 Giordano 2009) that were rescued and included.

#### Performance

- **Total records screened:** 2,633
- **Automatically excluded:** 2,414 (91.7%)
- **Flagged for manual adjudication:** 219 (8.3%) — final consensus decisions retained in the archived screening dataset
- **Sensitivity against 65 ultimately included studies:** 63/65 = 96.9% (2 rescued via post-hoc audit)
- **False negative rate:** 2/65 = 3.1% (both due to specific regex gaps that were subsequently corrected)

The archived screening files retain final decisions and audit reasons but do not consistently retain independent reviewer-level decisions for all records. The decision rules, regex patterns, and validation results are fully documented in the analysis code available in the Zenodo repository.

#### eText 3. Data Extraction Consensus Process

Data extraction used a two-reviewer consensus process. One reviewer performed the initial structured extraction using the standardized 185-variable form with AI-assisted tabulation support. The second reviewer independently checked extracted data against source reports, focusing on study characteristics, denominators, event counts, effect measures, confidence intervals, time points, intervention/comparator classification, and risk-of-bias judgments. Discrepancies were resolved by consensus before analysis. The repository includes `extraction_consensus_log.csv`, which records reviewer roles, consensus status, and correction flags for each included study.

The archived extraction dataset reports the final consensus values used in the analysis. Reviewer-level intermediate disagreement fields were not retained for every study, so the dataset should be interpreted as a consensus extraction file rather than a raw disagreement log. Three corrections were documented during the consensus process: MONITOR-HF HF hospitalization HR, CHAMPION PMID, and the CO-0056 all-cause mortality denominator. The CO-0056 correction changed the non-invasive telemonitoring mortality subgroup estimate; this is recorded as an audit correction rather than a new analytic choice.
